# The effectiveness, implementation, and experiences of peer support approaches for mental health: a systematic umbrella review

**DOI:** 10.1101/2023.10.23.23297394

**Authors:** Ruth Cooper, Katherine R.K. Saunders, Anna Greenburgh, Prisha Shah, Rebecca Appleton, Karen Machin, Tamar Jeynes, Phoebe Barnett, Sophie M. Allan, Jessica Griffiths, Ruth Stuart, Lizzie Mitchell, Beverley Chipp, Stephen Jeffreys, Brynmor Lloyd-Evans, Alan Simpson, Sonia Johnson

**Affiliations:** NIHR Mental Health Policy Research Unit, Institute of Psychiatry, Psychology and Neuroscience, King’s College London, London, UK; NIHR Mental Health Policy Research Unit, Division of Psychiatry, University College London, UK; Centre for Outcomes Research and Effectiveness, Research Department of Clinical, Educational and Health Psychology, University College London, UK; National Collaborating Centre for Mental Health, Royal College of Psychiatrists, London, UK; University of East Anglia, Norwich, UK; MHPRU Lived Experience Working Group; Florence Nightingale Faculty of Nursing, Midwifery and Palliative Care; Camden and Islington NHS Foundation Trust

## Abstract

**Background:** Peer support for mental health is recommended across international policy guidance and provision and has recently been expanded in countries including the UK. We conducted a systematic umbrella review, summarising the evidence from published reviews of the: 1) effectiveness, 2) implementation, and 3) experiences of paid peer support approaches for mental health.

**Methods:** We searched MEDLINE, EMBASE, PsycINFO, The Campbell Collaboration, and The Cochrane Database of Systematic Reviews (from January 2012-November 2022) for published reviews of paid peer support interventions for mental health. Review quality was assessed using the AMSTAR2. Results were synthesised narratively, with implementation reported in accordance with the Consolidated Framework for Implementation Research (CFIR).

**Results:** We included 35 reviews: systematic reviews with (n=13) or without (n=13) meta-analysis, systematic reviews with qualitative synthesis (n=3), and scoping reviews (n=6). The reviews included 426 primary studies and between 95-40,927 participants. Most reviews were low or critically low (97%) quality, one review was high quality. Effectiveness was investigated in 23 reviews. While results were mixed, there was some evidence from meta-analyses that peer support may improve depression symptoms (particularly in perinatal depression), self-efficacy, and recovery. Factors promoting successful implementation, investigated in 9 reviews, included adequate training and supervision, a recovery-oriented workplace, strong leadership, and a supportive and trusting workplace culture with effective collaboration. Barriers to implementation included lack of time, resources and funding, and a lack of recognised PSW certification. Experiences of peer support were explored in 11 reviews, with 3 overarching themes: i) what the peer support role could contribute, including recovery and improved wellbeing for both service users and peer support workers (PSWs); ii) confusion over the PSW role, including role ambiguity and unclear boundaries; and iii) organisational challenges, including low pay for PSWs, negative non-peer staff attitudes, and lack of support and training.

**Discussion:** Peer support may be effective at improving some clinical outcomes, self-efficacy, and recovery outcomes for some people. Evidence suggests that certain populations, such as perinatal populations, may especially benefit from peer support. Potential strategies to successfully implement PSWs in healthcare settings include coproduction, with clearly defined PSW roles, a receptive hierarchical structure and staff, strong leadership, and appropriate PSW and staff training with clinical and/or peer supervision alongside safeguarding. Services could also benefit from clear, coproduced, setting specific implementation guidelines for PSW. PSW roles tend to be poorly defined and associations between content of PSW interventions and their impacts needs further investigation. Future research in this area should reflect the priorities of those directly involved in peer support, either as providers or service users.

## Introduction

Peer support in mental health care is a recovery-orientated approach delivered by individuals who have lived experience of mental health difficulties (as service users, carers, parents, or supporters). Peer support workers (PSWs) are employed to draw on these experiences to support mental health service users or carers of people with mental health conditions [2,3]. As such, PSWs are uniquely positioned to facilitate recovery through empathic engagement with service users and their support networks. The success of peer support is thought to be based in the sharing of lived experiences and mental health knowledge and through interpersonal connection [4,5]. Across diagnoses, peer support may promote recovery through the modelling of coping strategies, and by providing hope and an example of recovery to those dealing with mental health difficulties [6].

Peer support has been utilised across various populations and types of service, for example in services for early intervention in psychosis [7], for people with co-occurring substance abuse and mental health difficulties [8], and in community interventions to reduce mental health inpatient admissions [9]. The format of peer support varies across services, for example it may involve one-to-one or group sessions, online or face-to-face delivery, unstructured open-ended conversations or more structured manualised support, or activities such as walking groups [10,11]. Peer support may be delivered by trained peer support staff or on a more ad-hoc basis among peers [12]. Peer support for mental health takes place both within mental health services and the voluntary sector [12]. Although PSWs may be paid or unpaid [7,13], paid roles have become increasingly available in mental health care settings [14]. Professionalising PSW roles as paid demonstrates the value of the role and appropriately rewards work done, should ensure formal training, supervision and management, and may help to clarify the boundaries of the role [15].

Service user networks and researchers in relevant fields have strongly advocated for provision of peer support [15,16], and peer support is now recognised and recommended across international mental health policy guidance, reflecting an increased understanding of the value of embedding lived experience support in formal mental health services [17–20]. In the UK, peer support is currently being expanded in the NHS [17].

There have been many reviews of the peer support literature separately evaluating the efficacy, implementation and experiences of peer support from a variety of different perspectives (e.g. [21–24]). Given the numerous and sometimes inconclusive results from existing reviews on this topic, our research group, the NIHR Mental Health Policy Research Unit, agreed with policy makers in England to conduct an umbrella review of peer support to provide clinicians, policy makers and researchers with an overall assessment on the evidence available, comparing results between reviews, while taking the quality of these reviews into account [25,26]. The aim of this systematic umbrella review is to collate, synthesise and summarise the available evidence from published reviews to address the following research questions:

1. What is the effectiveness (e.g., clinical, social, functional) and cost-effectiveness of paid peer support approaches for mental health?
2. What influences the implementation of peer support approaches for mental health?
3. What are the experiences of peer support approaches for mental health (e.g., of acceptability) from the perspective of peer support workers, healthcare practitioners, service users, carers?

## Methods

### This umbrella review was conducted by the NIHR Mental Health Policy Research Unit (MHPRU), based at King’s College London and University College London, which delivers evidence to inform government and NHS policy in England, agreeing a programme of rapid research with policymakers

### Study design and protocol

We conducted a systematic umbrella review following guidance from Fusar-Poli et al. [27] and Cochrane [28] and in accordance with the PRISMA statement (see appendix 1 for the PRISMA checklist). The protocol was registered with PROSPERO (registration number: CRD42022362099) [1].

### Lived experience researcher involvement

Members of the MHPRU Lived Experience Working Group (LEWG), who collectively have substantial experience of delivering or receiving peer support, contributed extensively to this review, including protocol development, study selection, data extraction, quality appraisal, data synthesis, drafting the manuscript and lived experience commentary, and attending working group meetings.

### Eligibility criteria

The eligibility criteria are detailed in full in the protocol [1]. In summary, we included:

#### Study designs

Published, peer reviewed systematic, scoping or realist reviews which synthesised quantitative or qualitative data (narratively or formally using e.g., a meta-analysis or meta-synthesis) that examined outcomes or experiences relevant to our research questions.

#### Intervention

We defined peer support as ‘involving a person who has lived experience of mental health condition(s), or caring for those with mental health conditions, being employed to use and draw on their experiences and empathy to support service users who have mental health conditions or carers or parents of people with mental health conditions.’ Eligible peer support approaches were paid, meaning that the peer support worker was paid for their work, and delivered face-to-face or remotely, for people with mental health conditions or for carers of people with mental health conditions, across any mental healthcare settings. Peer support approaches were ineligible if the peer support workers were not in a dedicated peer support role, if they were primarily for physical health, or automated (i.e. peer support ‘bots’ or avatars). We excluded reviews where over 50% of primary studies in the review did not meet eligibility criteria, e.g., if the majority of people delivering the interventions were unpaid.

#### Population

Children, young people, and adults with a mental health condition (including substance use disorders), carers, paid peer support workers, and mental healthcare practitioners working alongside peer support workers. We excluded service users with a primary diagnosis of an organic mental disorder (e.g., dementia), neurodevelopmental disorders, acquired cognitive impairment and adjustment disorders.

#### Outcome measure

Included reviews reported outcomes or data on at least one of the following peer support related outcomes that addressed our research questions: i) clinical outcomes, ii) economic or cost-effectiveness, iii) personal recovery outcomes e.g., hope, empowerment, goal-attainment, quality of life, iv) social outcomes, v) implementation outcomes and barriers and facilitators to implementation, vi) experiences of delivering, receiving or working alongside peer support and vii) theories of what works for whom in peer support.

### Information sources and search strategy

We combined terms for peer support, reviews, and mental health conditions using Boolean operators (AND, OR). We searched the following databases: MEDLINE, EMBASE, PsycINFO, The Campbell Collaboration, and The Cochrane Database of Systematic Reviews (see Appendix 2 for full search strategy). Searches were run from January 2012-November 2022 as these reviews will include primary research published before 2012 [29]. We had no language restrictions.

### Selection process

Reviewers (KS, RC, JG, RS, RA, KM, PS, SA) screened titles and abstracts, and subsequently full texts. To ensure consistent application of eligibility criteria all reviewers initially independently screened the same ten titles and abstracts and discussed inclusion/exclusion. The remaining titles and abstracts were then screened. Records were double screened blind by two reviewers at both the title and abstract (94% agreement) and full text (86% agreement) stages. All disagreements were resolved through discussion with the study team.

### Data extraction

Data extraction was completed in Microsoft Excel by the review team (RC, KS, KM, PS, JG, RS, PB, RA). The data used in the paper were checked by another member of the review team. The extracted data included: basic information about reviews (e.g. number of included studies, number of participants, review type, aim/objectives), basic information about primary studies (e.g. references, designs), search strategy (e.g. databases searched, eligibility criteria), population (e.g. gender, age), peer support approach (e.g. peer support type and description), type of comparator, additional information (e.g. quality appraisal methods, review author conclusions), primary and secondary outcomes of systematic review, or qualitative results.

### Quality appraisal of included reviews

The quality of included reviews was independently assessed by reviewers (RC, KS, KM, PS, JG, RS, PB, RA) using the AMSTAR 2 (A MeaSurement Tool to Assess systematic Reviews), a 16-point tool for assessment of the methodological quality of systematic reviews [30]. We adapted the AMSTAR 2 to apply for scoping reviews and systematic reviews of qualitative data, as described in Appendix 5. Two reviewers (KS, AG) 100% double-scored reviews blind with any outstanding disagreements resolved through discussion between AG, KS, and RC. Overall ratings for each study were calculated according to guidance [17], based on 7 critical domains and 6 non-critical domains within the AMSTAR 2 tool. Studies with no or one non-critical weakness and no critical flaws were rated as high quality. Studies with more than one non-critical weakness and no critical weaknesses were rated as moderate quality. Studies with one critical flaw irrespective of non-critical weaknesses were rated as low quality, and those with more than one critical flaw irrespective of non-critical weaknesses were rated as critically low quality. The AMSTAR 2 guidance [17] states that reviews of critically low quality should not be relied on for comprehensive and accurate summaries of the literature.

### Synthesis methods

**RQ1: What is the effectiveness (e.g., clinical, social, functional) and cost-effectiveness of paid peer support approaches for mental health?**

Data were tabulated and summarised narratively by two researchers (KS, AG); effectiveness meta-analysis data calculated from two or more studies were tabulated separately from non-meta-analysis effectiveness outcomes. Review outcomes were similar, but not similar enough to combine meaningfully in a meta-analysis. Effect sizes (with 95% CIs and p-values) were reported along with I^2^ statistic (with 95% CIs, p-values, Chi2, and degrees of freedom) where available. We did not tabulate data for subgroup analyses.

**RQ 2: What influences the implementation of peer support approaches for mental health?**

Outcomes were tabulated according to the main domains in the Consolidated Framework for Implementation Research (CFIR) [31]. The CFIR provides a comprehensive framework, composed of 5 domains, associated with the effective implementation of interventions [31]. Synthesis was conducted using a collaborative process involving one member of the study team (RA) and one lived experience researcher (PS).

**RQ 3: What are the experiences of peer support approaches for mental health (e.g., of acceptability) from the perspective of peer support workers, healthcare practitioners, service users, carers?**

Experiences were synthesised narratively, by three researchers, including two lived experience researchers (TJ, KM, RC) [32]. Themes from reviews which were identified as addressing research question 3 were extracted and similar themes across the reviews were grouped together. Each group was accounted for using an existing theme from one or more of the reviews or if this was not possible a new theme was developed. Three overarching themes were identified through iterative scrutiny of the data and discussion between TJ, KM and RC. A summary of the common themes across the reviews, grouped under the three overarching themes, was then developed, including highlighting contrasting findings.

## Results

### Study selection

The search strategy identified 777 references to be screened (a further 2 papers were identified through other methods); 93 full text articles were assessed for eligibility with 57 excluded (see Appendix 3 for reasons for exclusion). 35 reviews (reported in 36 papers) were included (see Figure 1).

**Figure 1.**
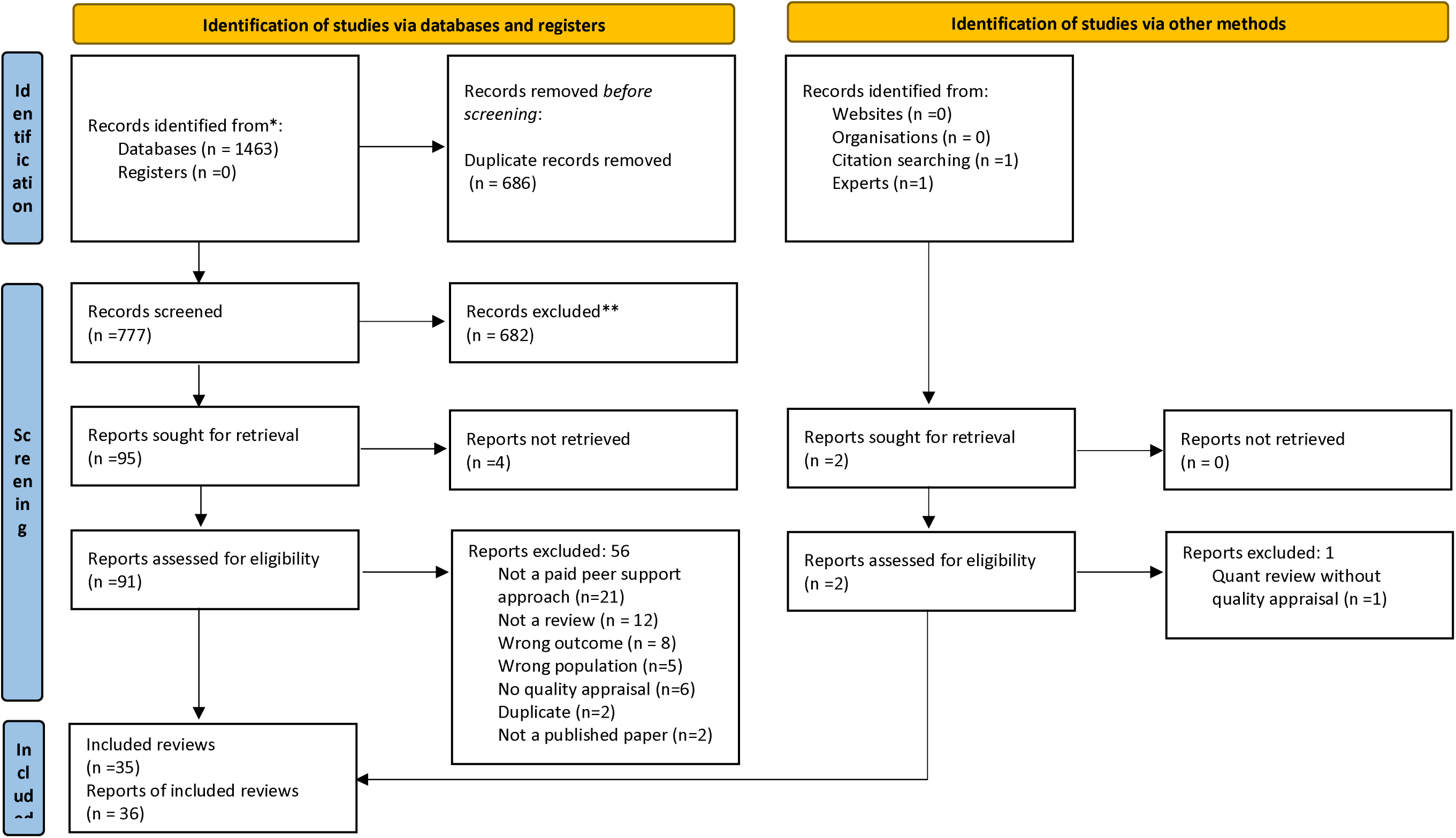
PRISMA flow diagram [64].

### Characteristics of included reviews

Review characteristics are detailed in Table 1. Of the 35 included reviews, 13 were systematic reviews with meta-analyses, 13 were systematic reviews without meta-analyses, 3 were systematic reviews with a qualitative synthesis, and 6 were scoping reviews. The individual reviews included between 95-40,927 participants; 6 reviews did not report the number of participants. For reviews where the population were service users, almost all were categorised as adults with mental health problems. Thirteen reviews specified that participants had severe mental illness (SMI) diagnoses [2,21,22,33–43], five reviews explicitly included studies with participants accessing mental health services [22,34,36,39,40] [44], three reviews were conducted in perinatal populations [45–47], three reviews included participants with any/common mental health conditions [48–50], four reviews included participants with substance use disorders [2,40,51,52], two reviews included participants with eating disorders [53,54], one included people experiencing suicidality [55], one included articles on peer support for crisis management [56]. The samples in the remaining reviews were PSWs and various stakeholders (e.g. non-peer staff, service users) [23,24,32,57–62]. Most reviews included interventions involving any form of peer support, individual, group or combined, although three reviews looked at group peer support alone [34,37,47], and three reviews looked at individual peer support alone [2,36,42]. Reviews looked at peer support delivered in-person, online, or over the phone; and surveyed a range of approaches including both structured and unstructured peer support (see Table 1). The reviews included 426 primary studies. We assessed study overlap; most primary studies (n=300) were only included in one review, however many primary studies were included twice (n=72), three times, (n=18) to a maximum of nine times (n=1) (see Appendix 4 for overlapping studies). Only 1 review reported that people with lived experience were involved in the review [55]. Only 2 reviews assessed certainty of evidence (using GRADE) [21,22].

**Table 1:**
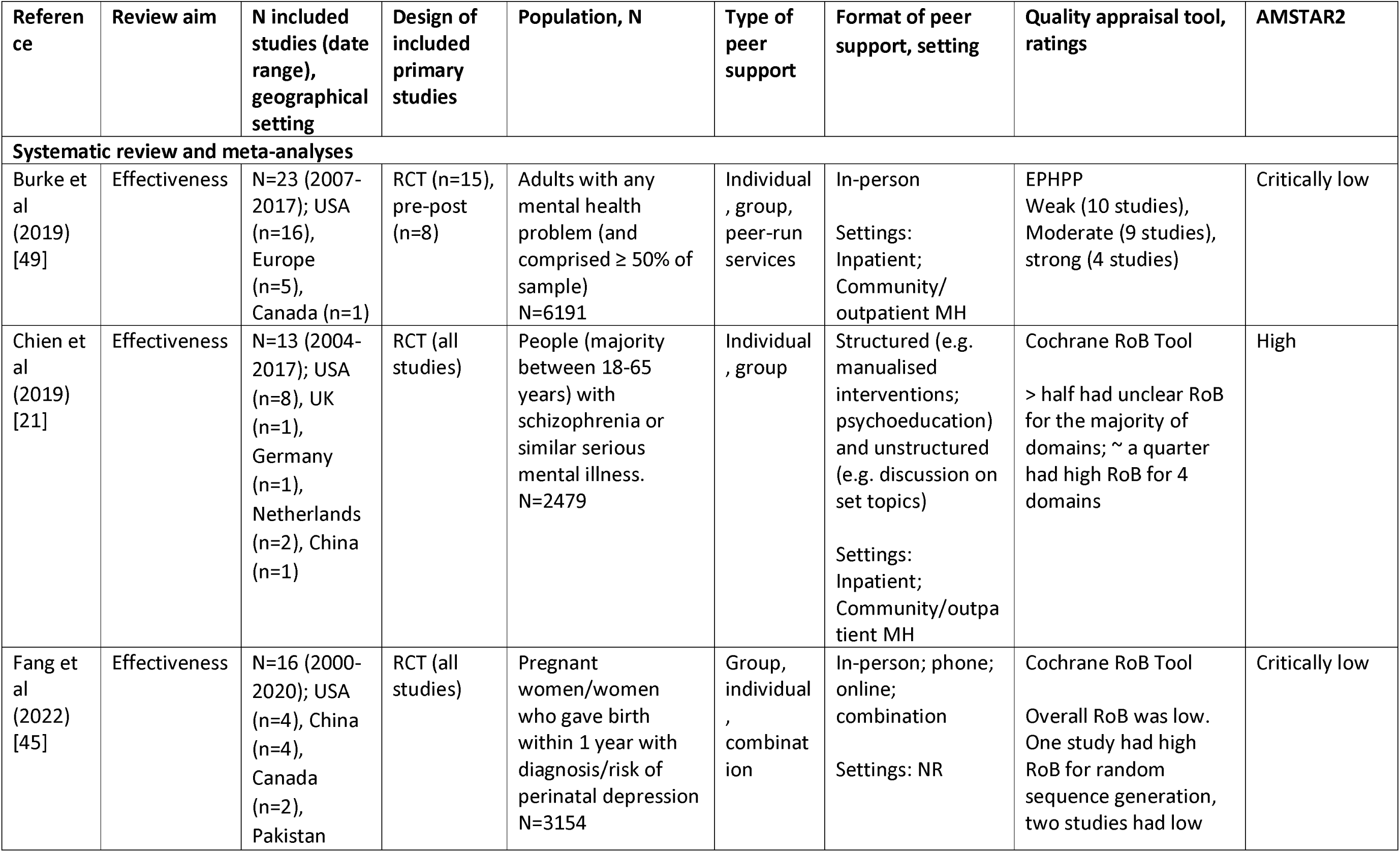
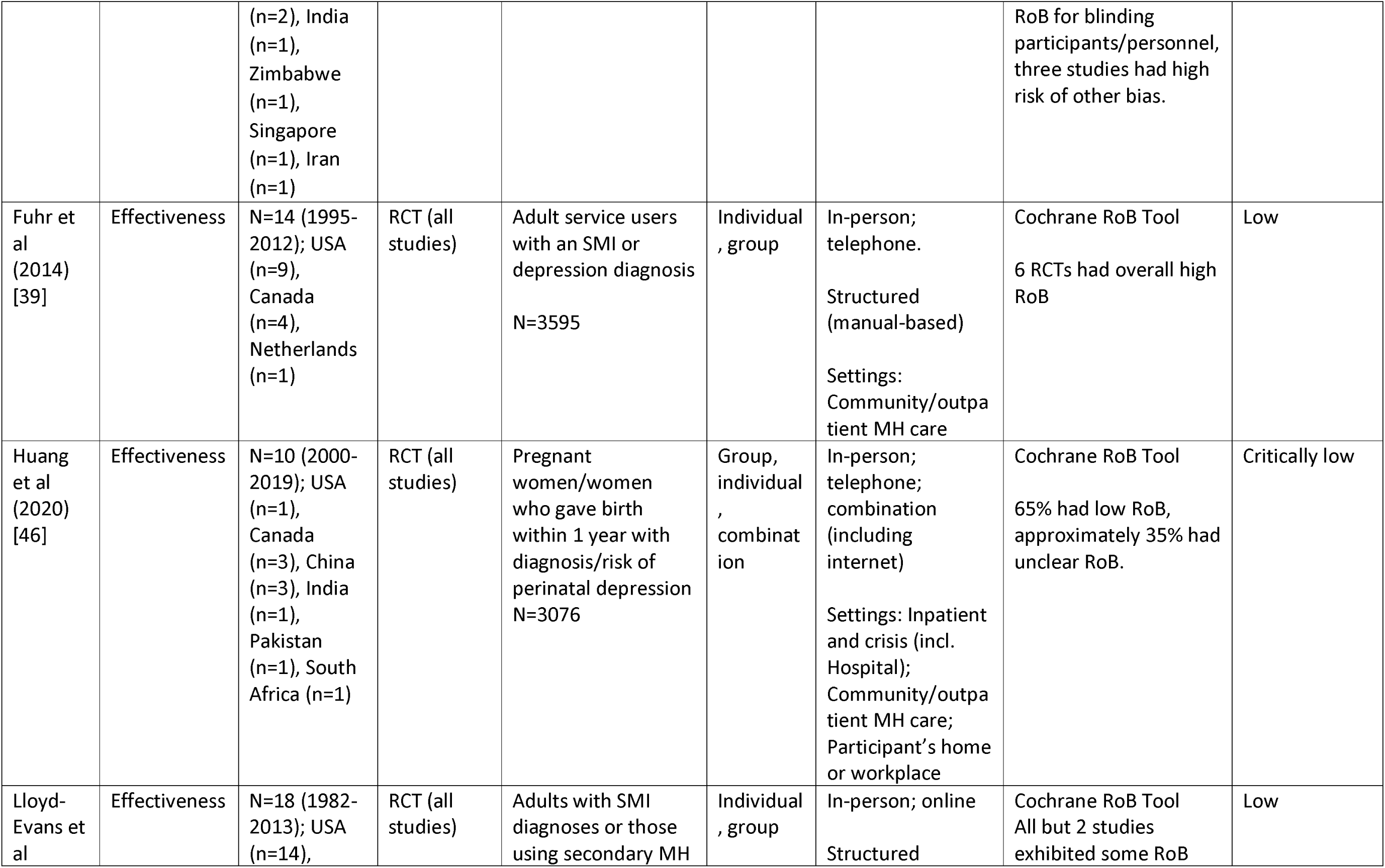
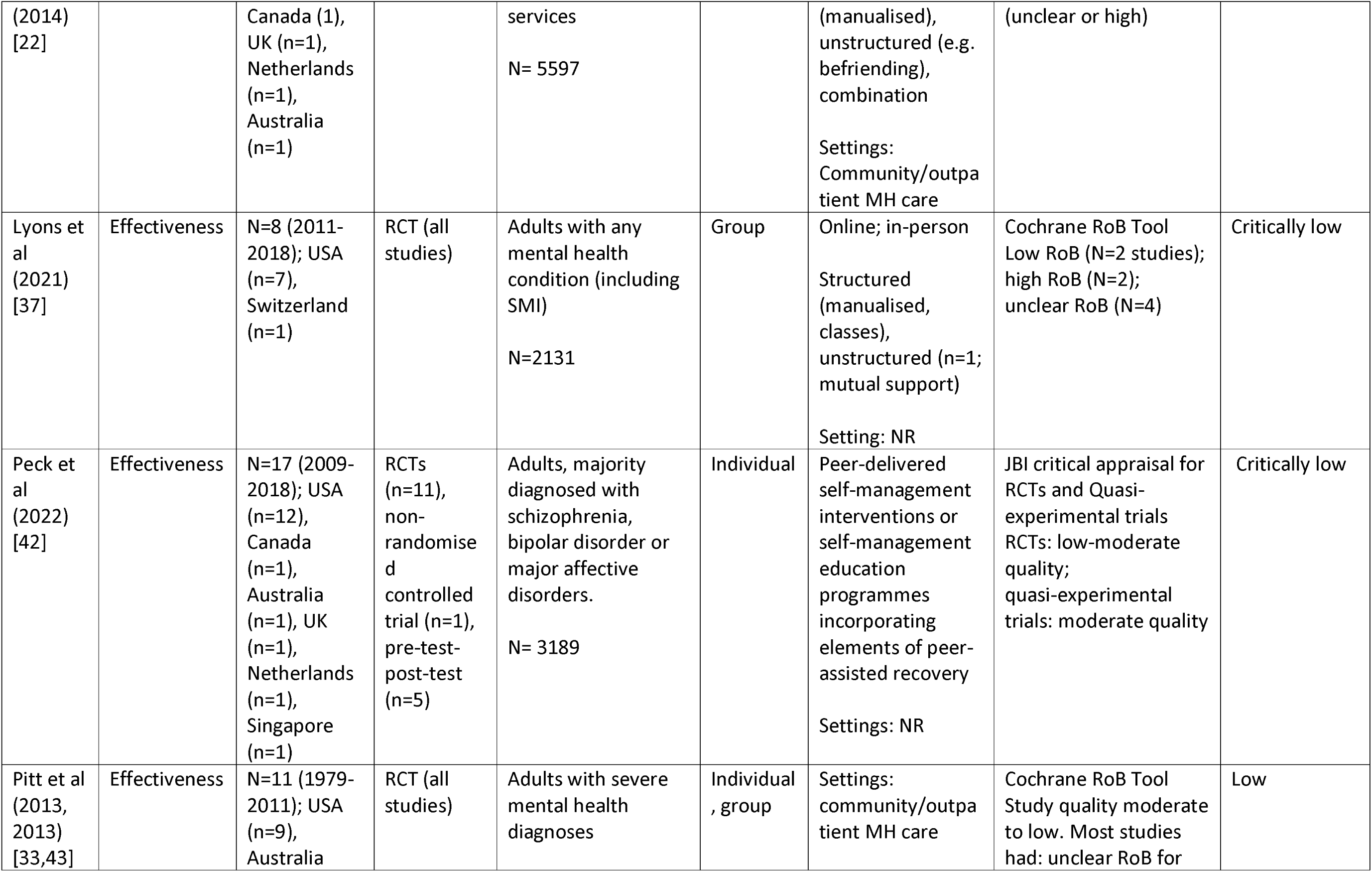
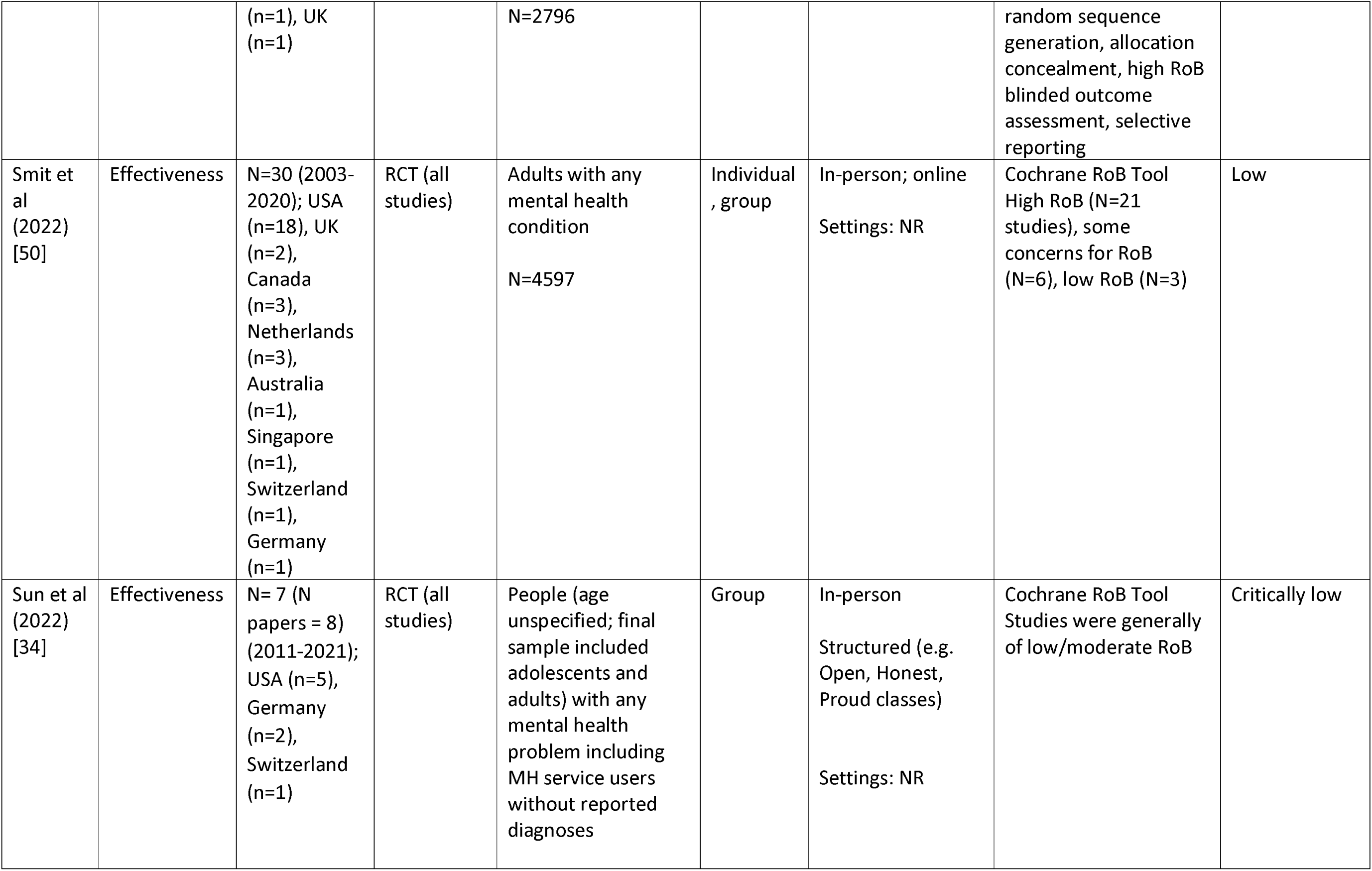
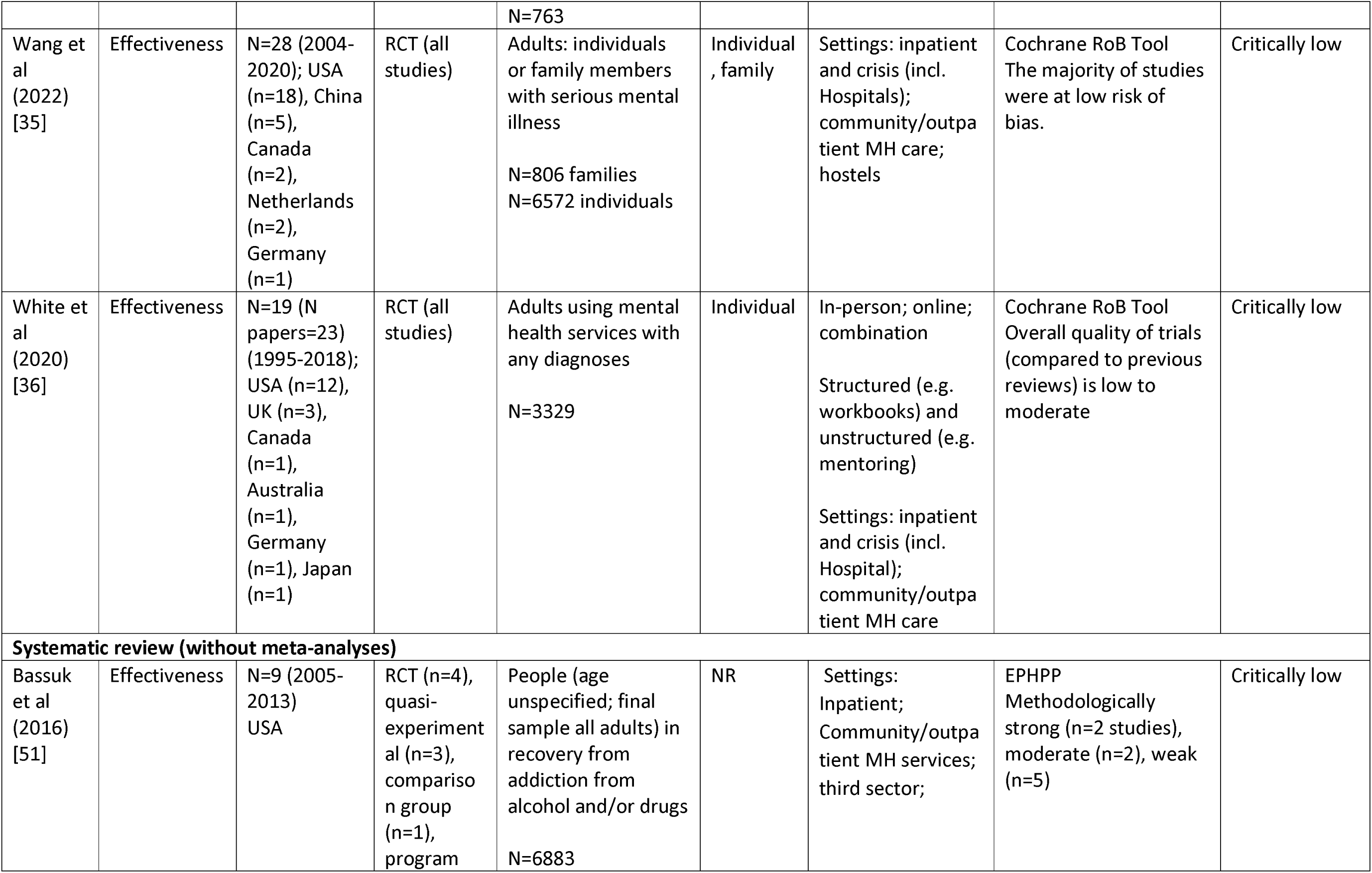
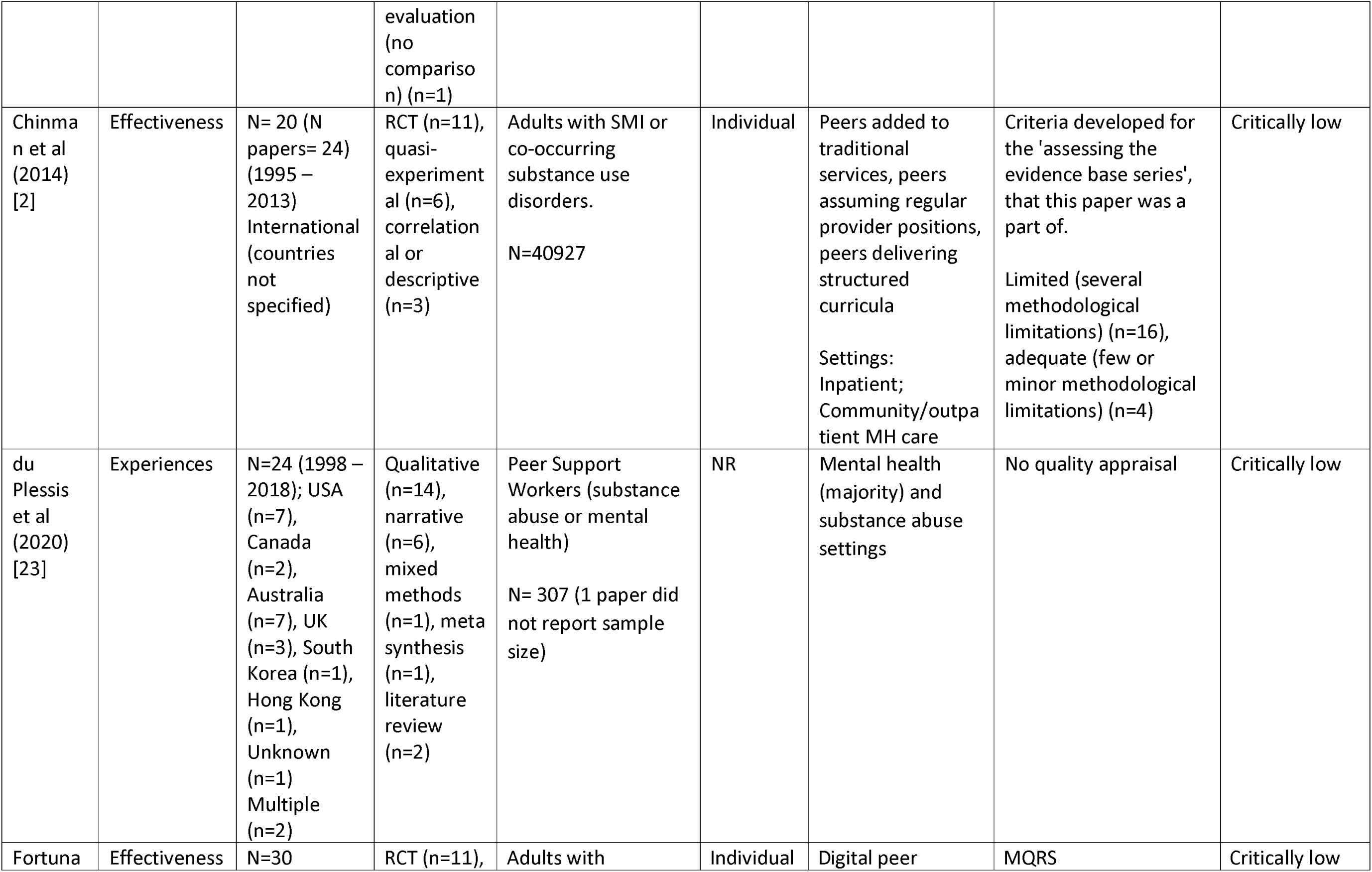
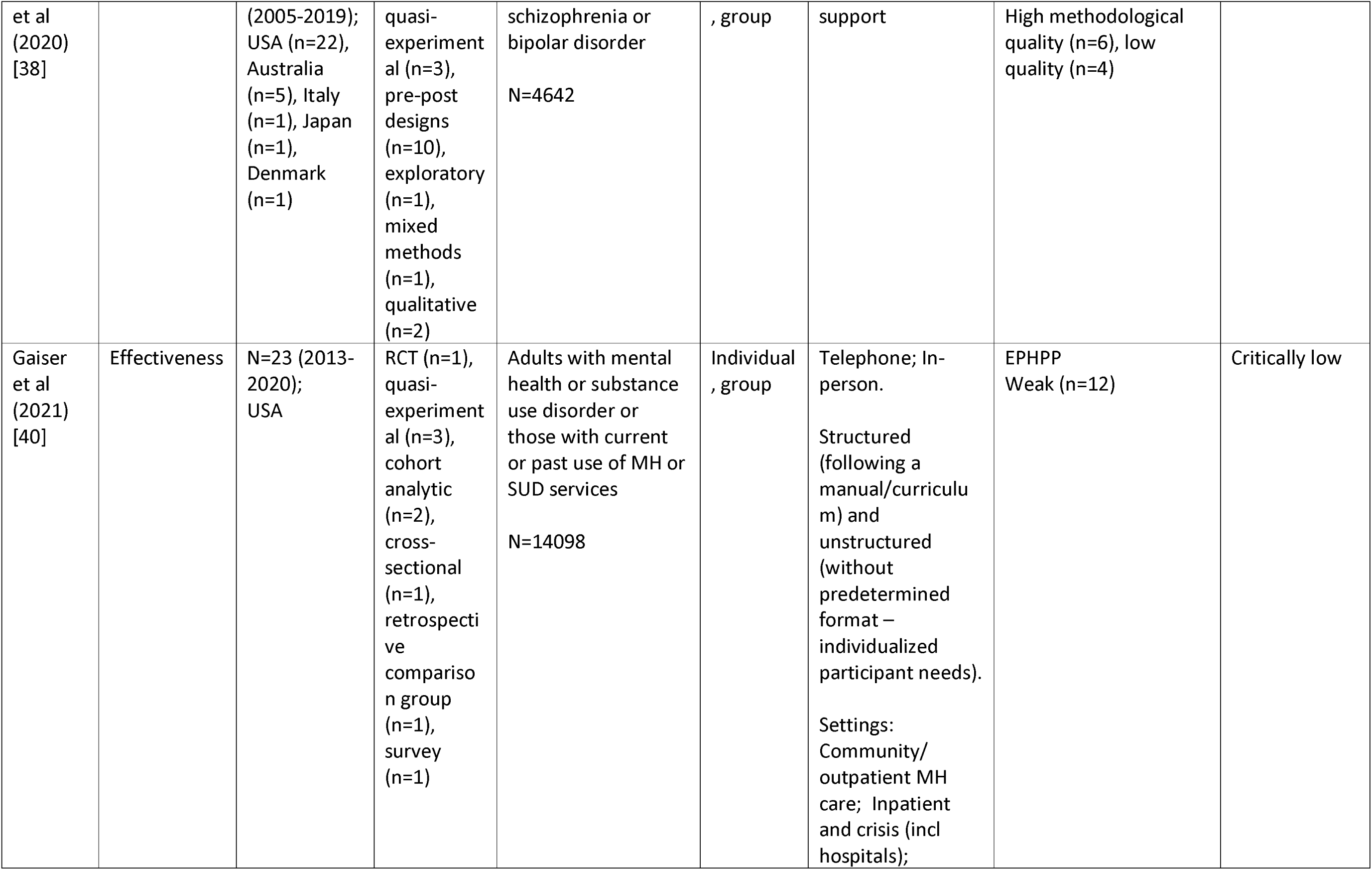
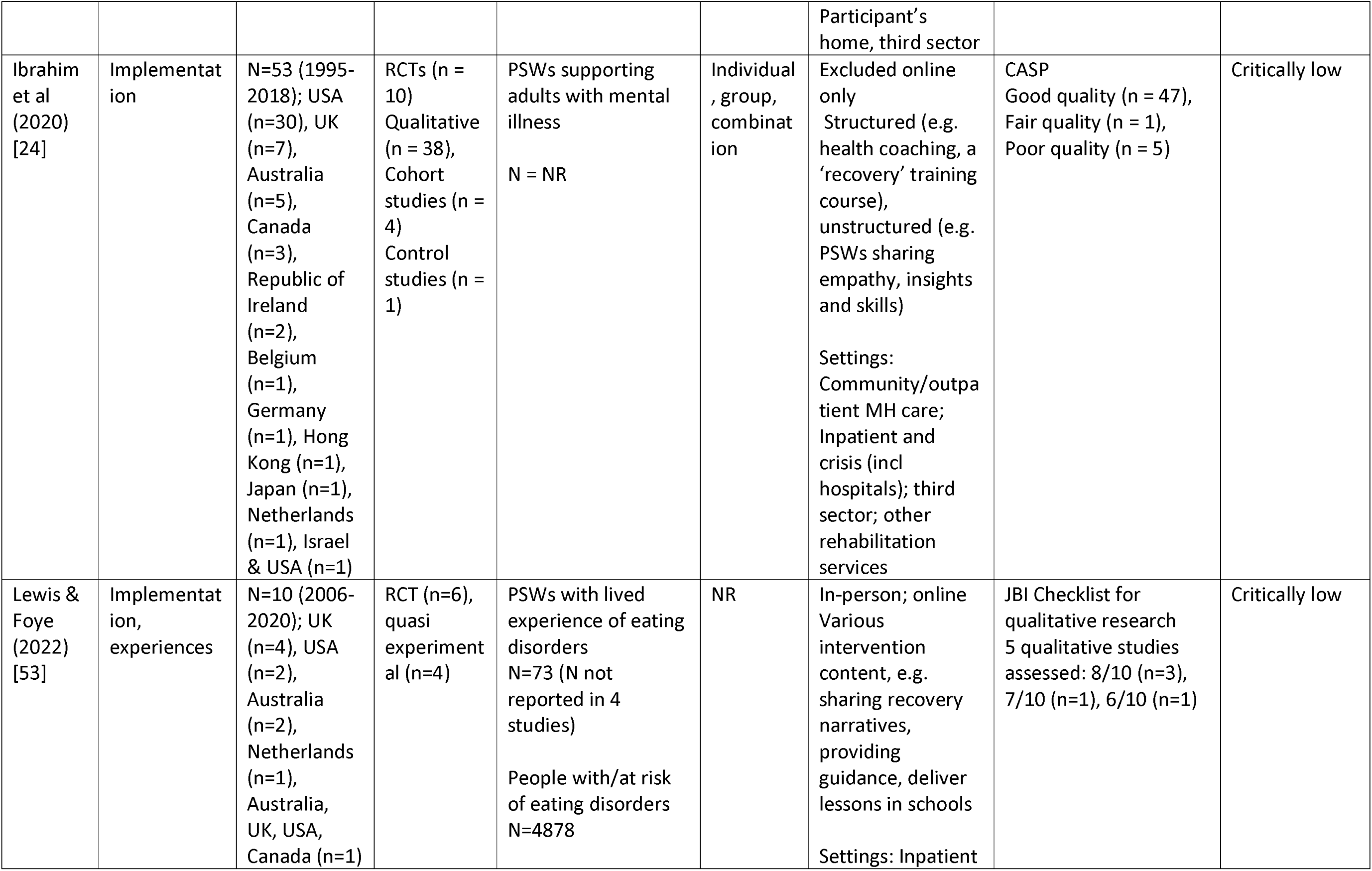
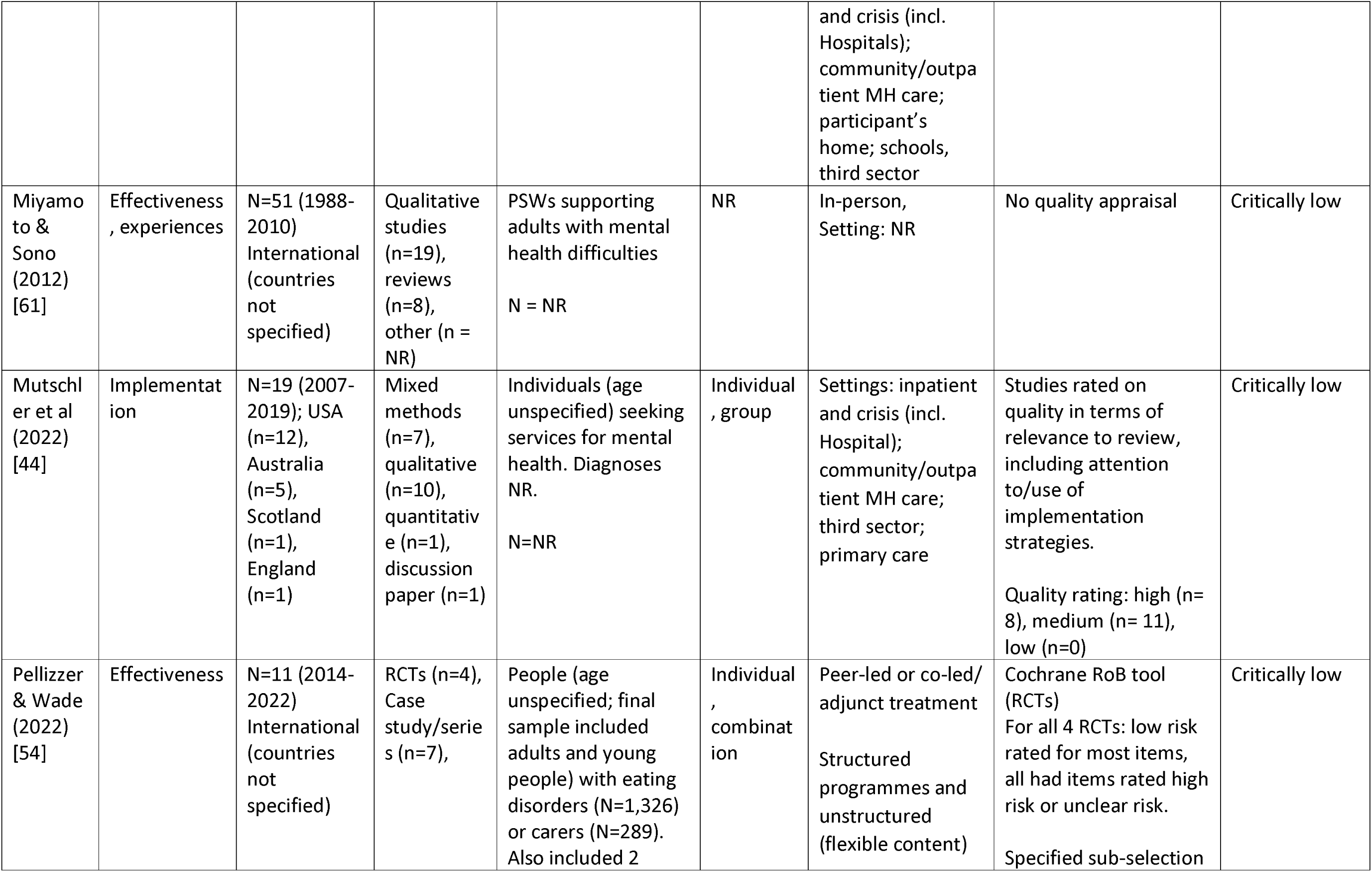
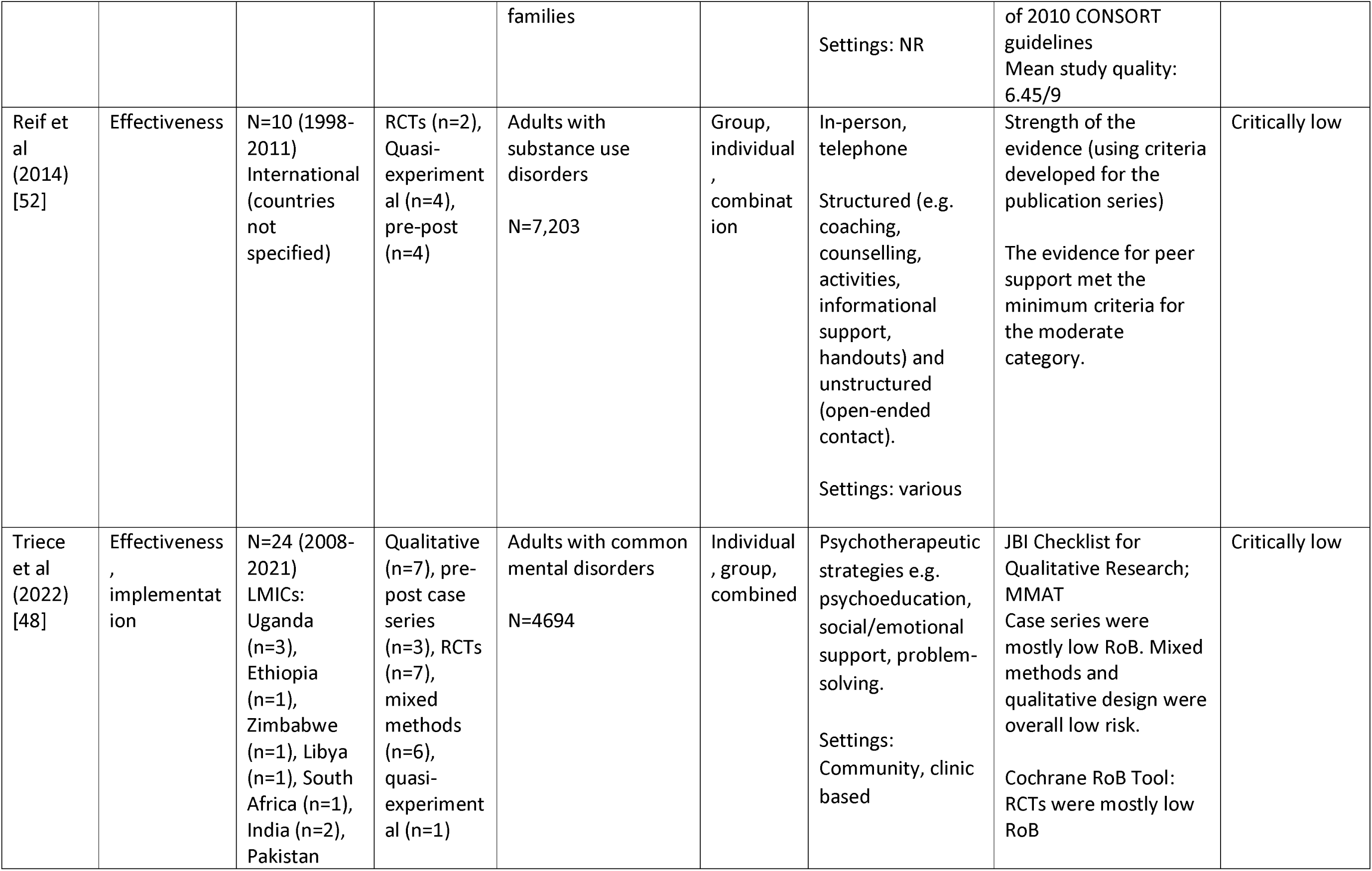
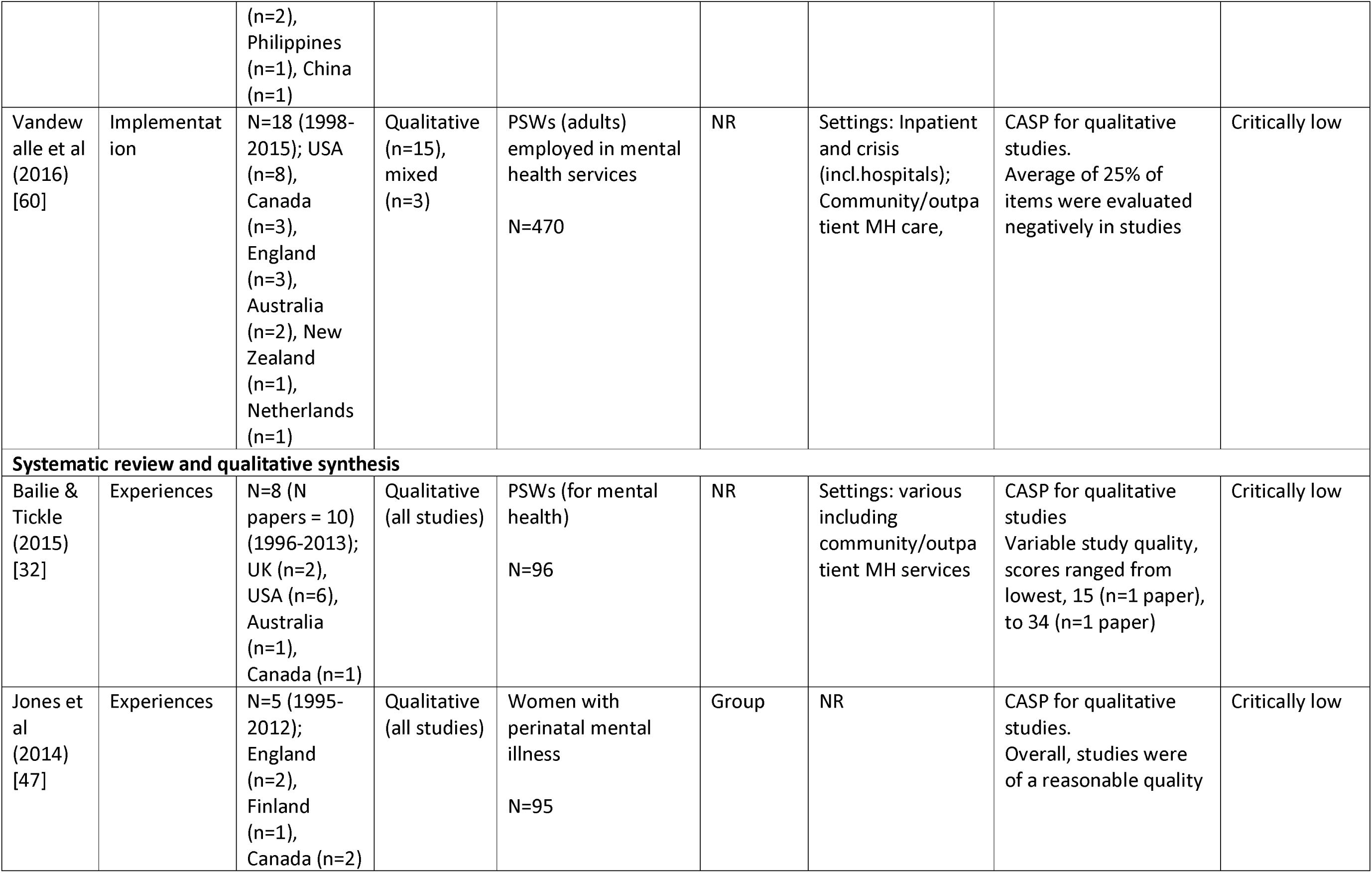
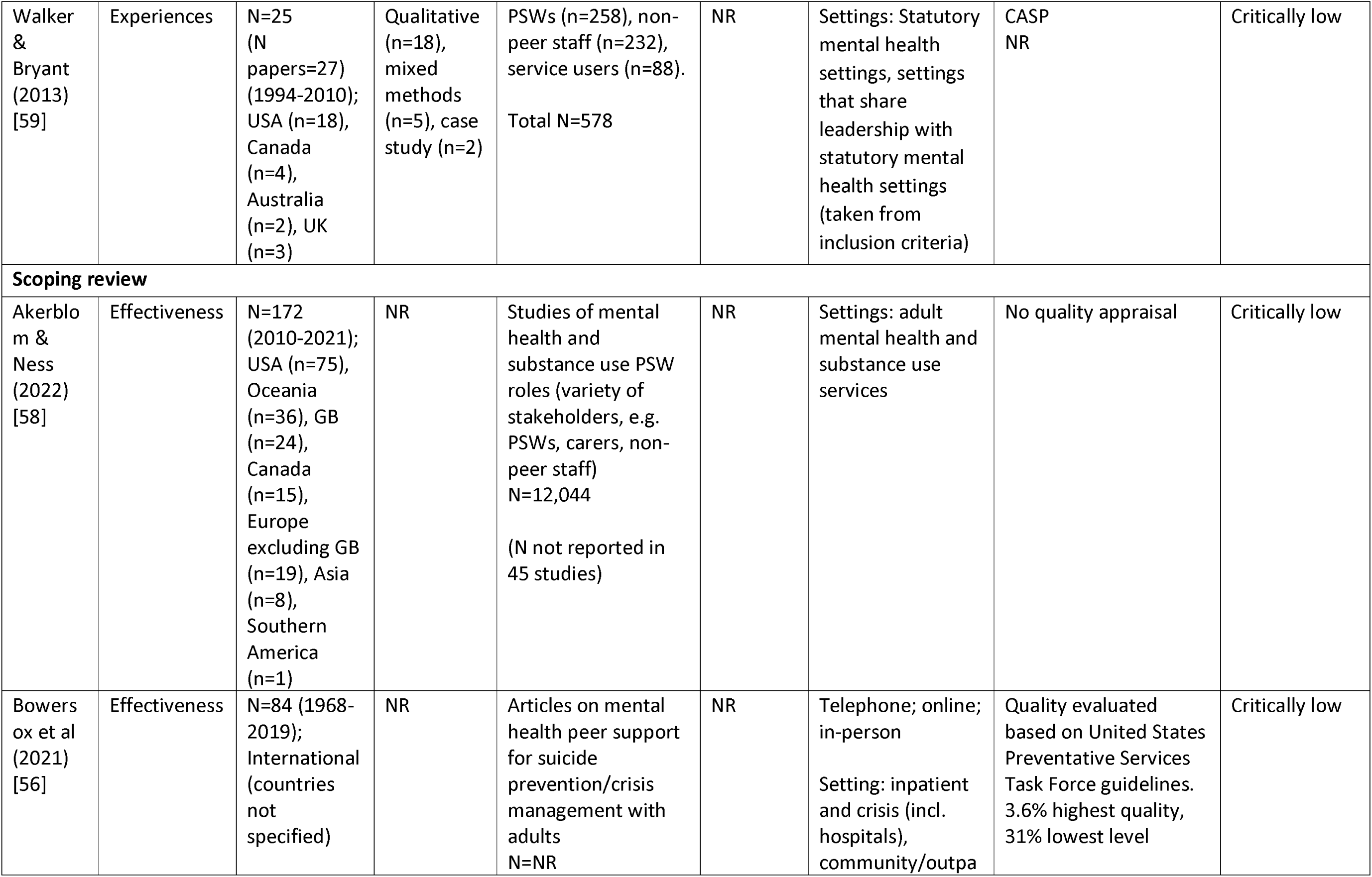
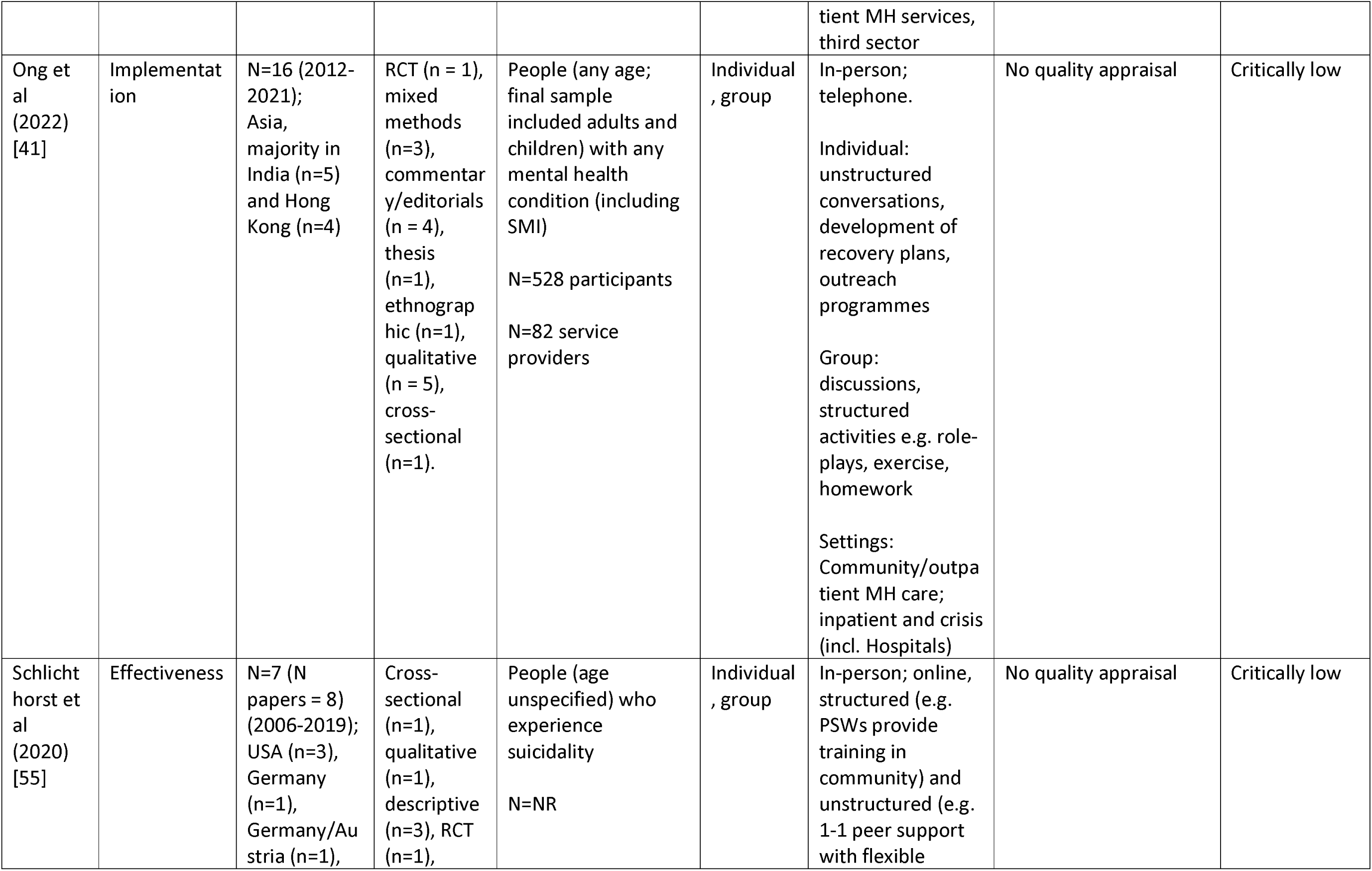
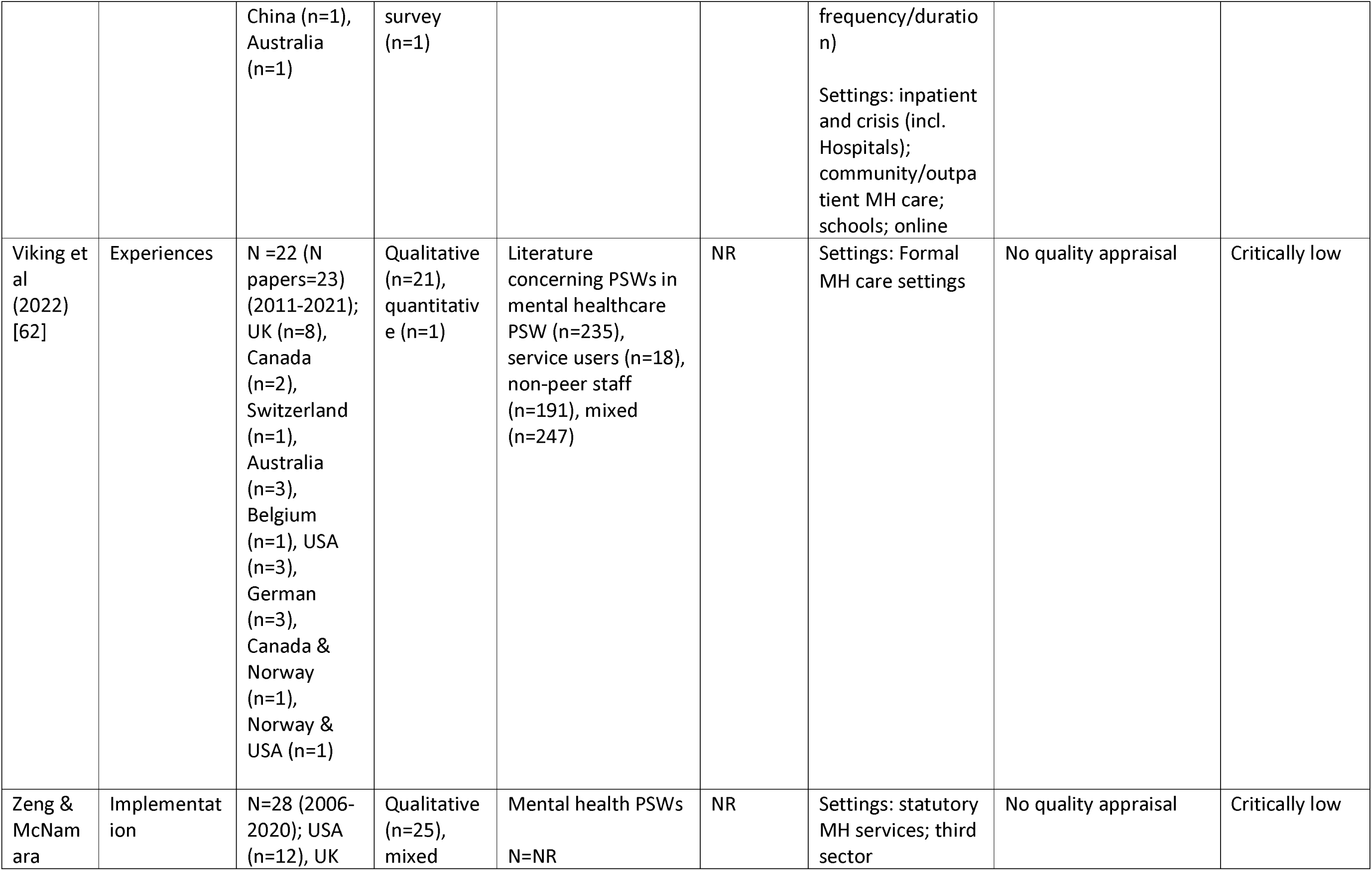
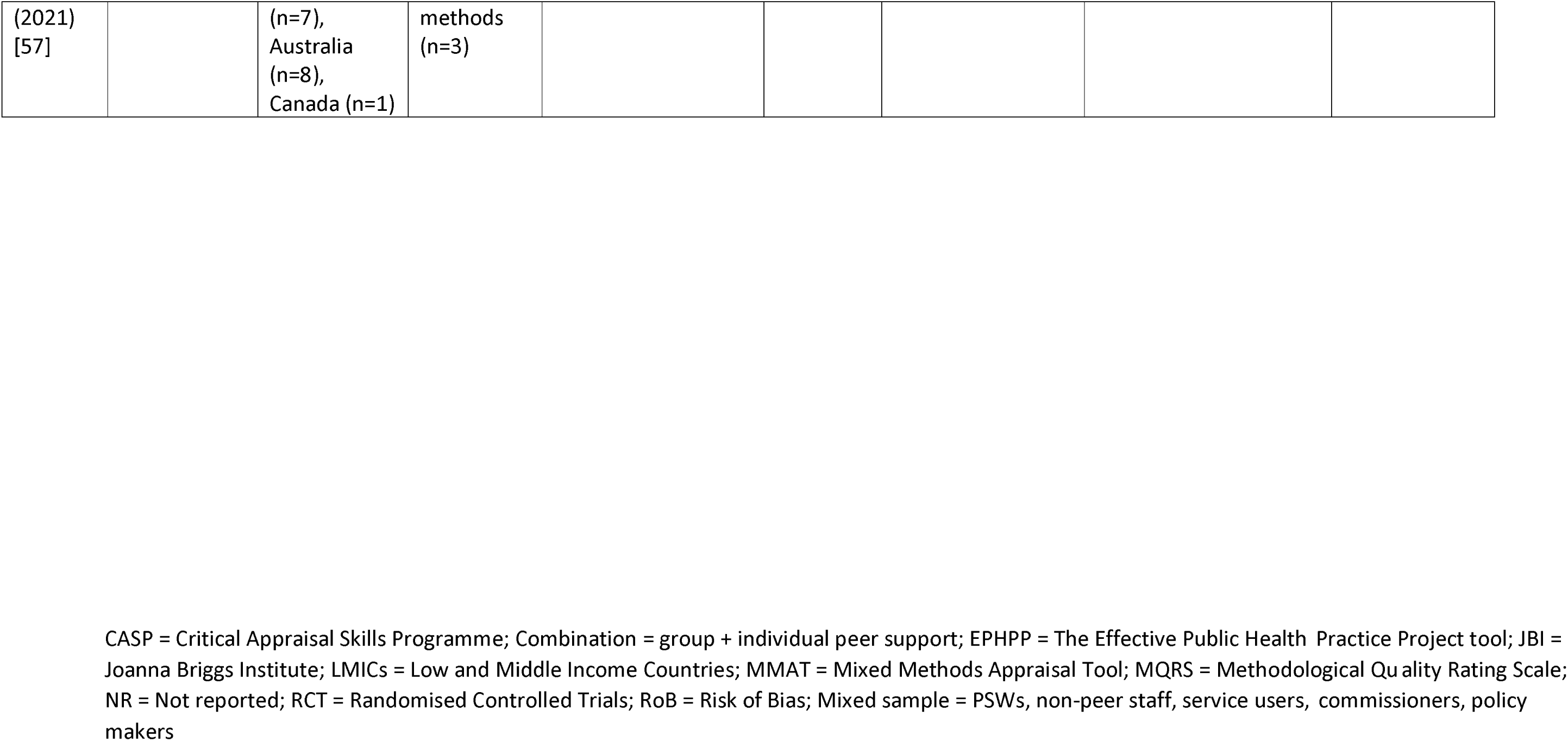
Characteristics of included studies.

### Quality appraisal of included reviews

Most reviews were appraised as low or critically low (97%) quality and one review was appraised as high quality. The most common weaknesses were in critical domains concerning registering protocols before commencement of the review (21 studies), justification of excluding individual studies (28 studies), and considering risk of bias when interpreting results (13 studies). Reviews without meta-analyses were not scored in the critical domains assessing meta-analytical method or publication bias. There were 13 studies with meta-analyses assessed in these two domains: two of these exhibited one critical weakness and two exhibited two critical weaknesses. As scoping reviews are intended to provide overviews of existing literature regardless of risk of bias [63], scoping reviews were not scored in the critical domain concerning risk of bias assessment techniques (see Appendix 5 for adjustments to quality appraisal for scoping and qualitative reviews). Of the 29 reviews that were eligible to be scored in this domain, 10 exhibited a critical weakness. The review eliciting high confidence was a Cochrane review [21]. No reviews were rated as moderate. AMSTAR 2 ratings are detailed in Table 1 and in full in Appendix 5.

## Results of synthesis

### RQ1: What is the effectiveness (e.g., clinical, social, functional) and cost-effectiveness of paid peer support approaches for mental health?

Effectiveness outcomes were reported in 23 reviews (66% of total). A wide variety of clinical, recovery-oriented and psychosocial effectiveness outcomes were reported across both meta-analysis [21,22,33–36,39,42,43,45,46,49,50] and narrative results [2,21,22,33–35,37–40,42,43,46,48,49,51,52,54–56,58]. Comparator groups also varied across the primary studies included in the reviews, including treatment as Usual (TaU), active controls (e.g., a comparable standard treatment) and waitlist control groups.

All outcomes except for one (Family or carer use of formal community support services; [35]) were service user outcomes, rather than carer, staff, or peer support worker outcomes. Outcomes from systematic reviews with meta-analysis are reported in Tables 2, 3 and 4. Effectiveness results from reviews not including meta-analysis are summarised at the end of this section and reported in full in Appendix 6. Evidence was heterogenous across all outcomes and reviews, with many analyses reporting no effect. In the meta-analysis results, there was often notable heterogeneity. There was limited data on cost and cost-effectiveness, but the evidence available from three systematic reviews without meta-analyses (See Appendix 6) suggested that peer support interventions were low cost and cost-saving [40,46,48].

**Table 2:** Meta-analyses effectiveness results: Clinical outcomes.

| Author (year) | Outcome | N of studies (N of participants ) | Population | Effect measure | Effect size (95% CI), p-value | Heterogeneity, $I^2$, 95% CI, $\chi^2$, df | AMSTAR2 | Summary findings |
| --- | --- | --- | --- | --- | --- | --- | --- | --- |
| Depression |  |  |  |  |  |  |  |  |
| Fang et al. (2022) [45] | Perinatal depression (various end time points) | 16 (3154) | Pregnant women/ women who gave birth within 1 year with diagnosis/ risk of perinatal depression | SMD | -0.39, (-0.54, -0.24); Z=9.42, $p<0.00001$ (results taken from text) | $I^2 = 78\%$; $\chi^2=91.38$ (df=20, $p<0.00001$) | Critically low | **Significant reduction in perinatal depression** |
| Huang et al. (2020) [46] | Depression (post-intervention) | 9 (1617) | Pregnant women/ women who gave birth within 1 year with diagnosis /risk of perinatal depression | SMD, Z | -0.37 (-0.66, -0.08), $p = 0.01$ Z=2.47 ($p=0.01$) | $I^2 = 84\%$, $p < 0.00001$, $\text{Tau}^2 = 0.14$; $\chi^2 = 49.37$ (df = 8, $p < 0.00001$) | Critically low | **Significant reduction in perinatal depression scores** |
| Huang et al. (2020) [46] | Depression 'events' (binary measure - post-intervention) | 7 (1644) | Pregnant women/ women who gave birth within 1 year with diagnosis /risk of perinatal depression | RR, Z | 0.69 (0.49, 0.96), $p = 0.03$ Z=2.22 ($p=0.03$) | $I^2 = 70\%$, $p = 0.003$ $\text{Tau}^2 = 0.11$; $\chi^2 = 20.25$ (df=6; $p=0.003$) | Critically low | **Significant reduction in risk of perinatal depression** |
| Lloyd-Evans et al. (2014) [22] | Depression and anxiety (post-intervention) | 3 (861) | Adults with SMI or those using secondary MH services | SMD | -0.10 (-0.24, 0.03) | $I^2 = 0\%$, $\chi^2 = 1.97$ ($p=0.37$) | Low | No effect |
| Lyons et al. (2021) [37] ** | Depression (post-intervention) | 4 (929) | Adults with any mental health condition (including SMI) | SMD | -0.09 (-0.22, 0.04), $p=0.18$ | $I^2 = 0\%$, $\chi^2=1.11$ (df = 2) | Critically low | No effect |
| Sun et al. (2022) [34] ** | Depression (end-of-treatment) | 5 (372) | Adults and adolescents with any mental health problem including MH service users without reported diagnoses | SMD | -0.05 (-0.26, 0.15), p=0.34 | I<sup>2</sup>=0%, Chi<sup>2</sup>=1.62 (df=1, p=0.76) | Critically low | No effect |
| Depression: follow up |  |  |  |  |  |  |  |  |
| Lloyd-Evans et al. (2014) [22] | Depression and anxiety (6-month follow-up) | 2 (721) | Adults with SMI or those using secondary MH services | SMD | -0.17 (-0.32, -0.03) | I<sup>2</sup>=0%, Chi<sup>2</sup> = 0.15 (p=0.70) | Low | Significant reduction in depression and anxiety |
| Lyons et al. (2021) [37] ** | Depression (3–6-month follow-up) | 3 (674) | Adults with any mental health condition (including SMI) | SMD | -0.12 (-0.27, 0.03), p=0.11 | I<sup>2</sup> = 0%, Chi<sup>2</sup> = 0.95 (df = 2) | Critically low | No effect |
| Clinical recovery |  |  |  |  |  |  |  |  |
| Smit et al. (2022) [50] | Clinical recovery (post-intervention) | 22 (NR) | Adults with any mental health diagnosis | Hedges' g | 0.19 (0.11–0.27), p<0.001 | I<sup>2</sup>= 10%, (95% CI 0–44) | Low | Significant improvement in clinical recovery |
| Clinical recovery: follow up |  |  |  |  |  |  |  |  |
| Smit et al. (2022) [50] | Clinical recovery (6-9 months follow up) | 13 (NR) | Adults with any mental health diagnosis | Hedges' g | 0.17 (0.08–0.26), p=0.002 | I<sup>2</sup>=0% (95% CI 0–57) | Low | Significant improvement in clinical recovery |
| Smit et al. (2022) [50] | Clinical recovery (12–18-month follow up) | 8 (NR) | Adults with any mental health diagnosis | Hedges' g | 0.10 (-0.21,0.40), p=0.48 | I<sup>2</sup>=63% (95% CI 20–83) | Low | No effect |
| Mental health symptoms |  |  |  |  |  |  |  |  |
| Peck et al. (2022) | Symptom severity | 5 (1094) | Adults, majority diagnosed with | SMD, Z | -0.30 (-0.55, -0.04), Z=2.29 | I<sup>2</sup>=75%, Tau<sup>2</sup>= 0.06, Chi<sup>2</sup>=15.77 (df=4, | Critically low | Significant reduction in symptom severity |
| [42] * | (time point not stated) |  | schizophrenia, bipolar disorder or major affective disorders. |  | (p=0.02) | p=.003) |  |  |
| Wang et al. (2022) [35] | Psychotic symptoms (post-family led peer support) | 3 (742) | Adult individuals or family members with SMI | SMD, Z | -1.45, (-2.68, -0.22), Z=2.32, p=0.02 | I<sup>2</sup>=98%, Tau<sup>2</sup>=1.92; Chi<sup>2</sup>=189.37 (df=4, p<0.00001) | Critically low | **Significant reduction in psychotic symptoms** |
| Lloyd-Evans et al. (2014) [22] | Overall psychiatric symptoms (post-treatment) | 3 (753) | Adults with SMI or those using secondary MH services | SMD | -0.07 (-0.39, 0.24) | I<sup>2</sup> = 74%, Chi<sup>2</sup> = 7.83 (p=0.02) | Low | No effect |
| Lloyd-Evans et al. (2014) [22] | Symptoms of psychosis (post-treatment) | 2 (696) | Adults with SMI or those using secondary MH services | SMD | -0.08 (-0.27, 0.03) | Not reported | Low | No effect |
| Lyons et al. (2021) [37] ** | Global symptoms (post-intervention) | 3 (823) | Adults with any mental health condition (including SMI) | SMD | -0.13 (-0.27, 0.01), p=0.07 | I<sup>2</sup> = 0%, Chi<sup>2</sup> = 1.11 (df=2) | Low | No effect |
| Pitt et al. (2013) [33,43] | Mental health symptoms (time point not stated) | 2 (197) | Adults with severe mental health diagnoses | SMD, Z | -0.24 (-0.52, 0.05), Z=1.65 (p=0.1) | I<sup>2</sup>=0%, Tau<sup>2</sup>=0; Chi<sup>2</sup>=0.52 (df=2; p=0.77) | Low | No effect |
| Sun et al. (2022) [34] ** | Anxiety (post-intervention) | 2 (175) | Adults and adolescents with any mental health problem including MH service users without reported diagnoses | SMD | 0.29, (-0.01, 0.58), p=0.06 | I<sup>2</sup>=0%, Chi<sup>2</sup>=0.09 (df=1, p=0.76) | Critically low | No effect |
| Wang et al. (2022) [35] | Psychotic symptoms (post individual-led peer support) | 11 (2651) | Adult individuals or family members with SMI | SMD | -0.30, (-0.73, 0.13), Z=1.38, p=0.17 | I<sup>2</sup>=96%; Tau<sup>2</sup>=0.5; Chi<sup>2</sup>= 270.29 (df=10, p<.00001) | Critically low | No effect |
| Mental health symptoms: follow up |  |  |  |  |  |  |  |  |
| White et al. (2020) [36] * | Psychiatric symptoms (6-24 months follow up) | 6 (857) | Adults using mental health services with any diagnoses | SMD, Z | -0.01 (-0.21, 0.20), Z=0.0, (p=0.961) | I<sup>2</sup>=53%, Chi<sup>2</sup>=10.7, p=0.057 | Critically low | No effect |
| Service use |  |  |  |  |  |  |  |  |
| Wang et al. (2022) [35] | Rehospitalisation (last follow up) | 3 (483) | Adult individuals or family members with SMI | SMD, Z | -1.34, (-1.94, -0.75), Z=4.44, p<0.00001 | I<sup>2</sup>=87%, Tau<sup>2</sup>=0.32; Chi<sup>2</sup>=23.15 (df=3, p<0.0001) | Critically low | **Significant reduction in rehospitalisation** |
| Wang et al. (2022) [35] | Duration of hospitalisation (last follow up) | 3 (483) | Adult individuals or family members with SMI | SMD, Z | -1.48, (-2.56, -0.41), Z=2.70, p=0.007 | I<sup>2</sup>=96%, Tau<sup>2</sup>=1.14; Chi<sup>2</sup>=70.97 (df=3, p<0.0001) | Critically low | **Significant reduction in hospital duration** |
| Wang et al. (2022) [35] | Family/carer use of formal community support services (last follow up) | 4 (483) | Adult individuals or family members with SMI | SMD, Z | -1.38, (2.19, -0.56), Z=3.32, p=0.0009 | I<sup>2</sup>=93%, Tau<sup>2</sup>=0.64, Chi<sup>2</sup>=42.21 (df=3, p<0.00001) | Critically low | **Significant reduction in use of community support services** |
| Lloyd-Evans et al. (2014) [22] | Duration of admission (post-treatment) | 3 (255) | Adults with SMI or those using secondary MH services | SMD | -0.22 (-0.72, 0.28) p=0.03 | I<sup>2</sup> = 72%, Chi<sup>2</sup> = 7.16 | Low | No effect |
| Pitt et al. (2013) | Length of stay (time) | 2 (119) | Adults with severe mental health | MD, Z | -13.41 (-32.09, 5.27), Z=1.41 | I<sup>2</sup>=28.6%, Tau<sup>2</sup>=89.38; | Low | No effect |
| [33,43] | point not stated) |  | diagnoses |  | (p=0.16) | Chi2=1.4 (df=1, p=0.24) |  |  |
| Service use: follow up |  |  |  |  |  |  |  |  |
| White et al. (2020) [36] * | Risk of hospitalisation (3-24 months follow up) | 5 (497) | Adults using mental health services with any diagnoses | RR, Z | 0.86 (0.66, 1.13), Z=1.1 (p=0.27) | I<sup>2</sup>=38%, Chi<sup>2</sup>=6.5, p=0.170 | Critically low | **Significant reduction in risk of hospitalisation** |
| White et al. (2020) [36] * | Days in hospital (9-24 months follow up) | 5 (453) | Adults using mental health services with any diagnoses | SMD, Z | -0.10 (-0.34, 0.14), Z=0.8, p=0.426 | I<sup>2</sup>=39%, Chi<sup>2</sup> (Q)=10.7, p=0.057 | Critically low | No effect |
| Other clinical outcomes |  |  |  |  |  |  |  |  |
| Sun et al. (2022) [34] ** | Help-seeking (post-intervention) | 2 (114) | Adults and adolescents with any mental health problem including MH service users without reported diagnoses | SMD | 0.46, (0.10, 0.82), p=0.01 | I<sup>2</sup>=0%, Chi<sup>2</sup>=0.93 (df=1, p=0.34) | Critically low | **Significant improvement in help-seeking** |
| Sun et al. (2022) [34] ** | Disclosure-related distress (post-intervention) | 2 (170) | Adults and adolescents with any mental health problem including MH service users without reported diagnoses | SMD | -0.53 (-0.84, -0.23), p=0.0006 | I<sup>2</sup>=0%, Chi<sup>2</sup>=0.25 (df=2, p=0.61) | Critically low | **Significant decrease in disclosure related distress** |
| Wang et al. (2022) [35] | Medication adherence (last follow up) | 5 (371) | Adult individuals or family members with SMI | SMD | -0.22, (-0.43, -0.01); Z=2.08, p=0.04 | I<sup>2</sup>=0%; Tau2=0; Chi2=1.54 (df=4, p=0.82) | Critically low | **Significant improvement in medical adherence** |
| Wang et | Activation | 3 | Adult individuals or | SMD | 0.43, (0.19, | I<sup>2</sup>=19%; Tau2=0.01; | Critically low | **Significant improvement** |
| al. (2022) [35] | (last follow up) | (375) | family members with SMI |  | 0.67); Z=3.46, p=0.0005 | Chi2=2.47 (df=2, p=0.29) |  | **in activation** |
| Sun et al. (2022) [34] ** | Disclosure-related withdrawal (post-intervention) | 3 (281) | Adults and adolescents with any mental health problem including MH service users without reported diagnoses | SMD | -0.10 (-0.33, 0.14), p=0.42 | I<sup>2</sup>=37%, Chi<sup>2</sup>=3.20 (df=2, p=0.20) | Critically low | No effect |
| Wang et al. (2022) [35] | Alcohol use (last follow up) | 3 (257) | Adult individuals or family members with SMI | SMD | -0.23, (-0.49, 0.03); Z=1.73, p=0.08 | I<sup>2</sup>=0%; Tau2=0; Chi2=1.36, (df=2, p=0.51) | Critically low | No effect |
| Other clinical outcomes: follow up |  |  |  |  |  |  |  |  |
| Chien et al. (2019) [21] | Activation (medium term, 1-6 month follow up) | 3 (295) | Adults with schizophrenia or similar SMI | MD, Z | 3.68 (-1.85, 9.22), Z=1.3 (p=0.19) | I<sup>2</sup>=80.32, Tau<sup>2</sup>=18.09, Chi<sup>2</sup>=10.16 (df=2, p=0.01) | High | No effect |

*\*=Review included studies of individual peer support only; \*\*= Review included studies of group peer support only; no stars= Review included studies of either individual or group peer support, or both.*

**Table 3:**
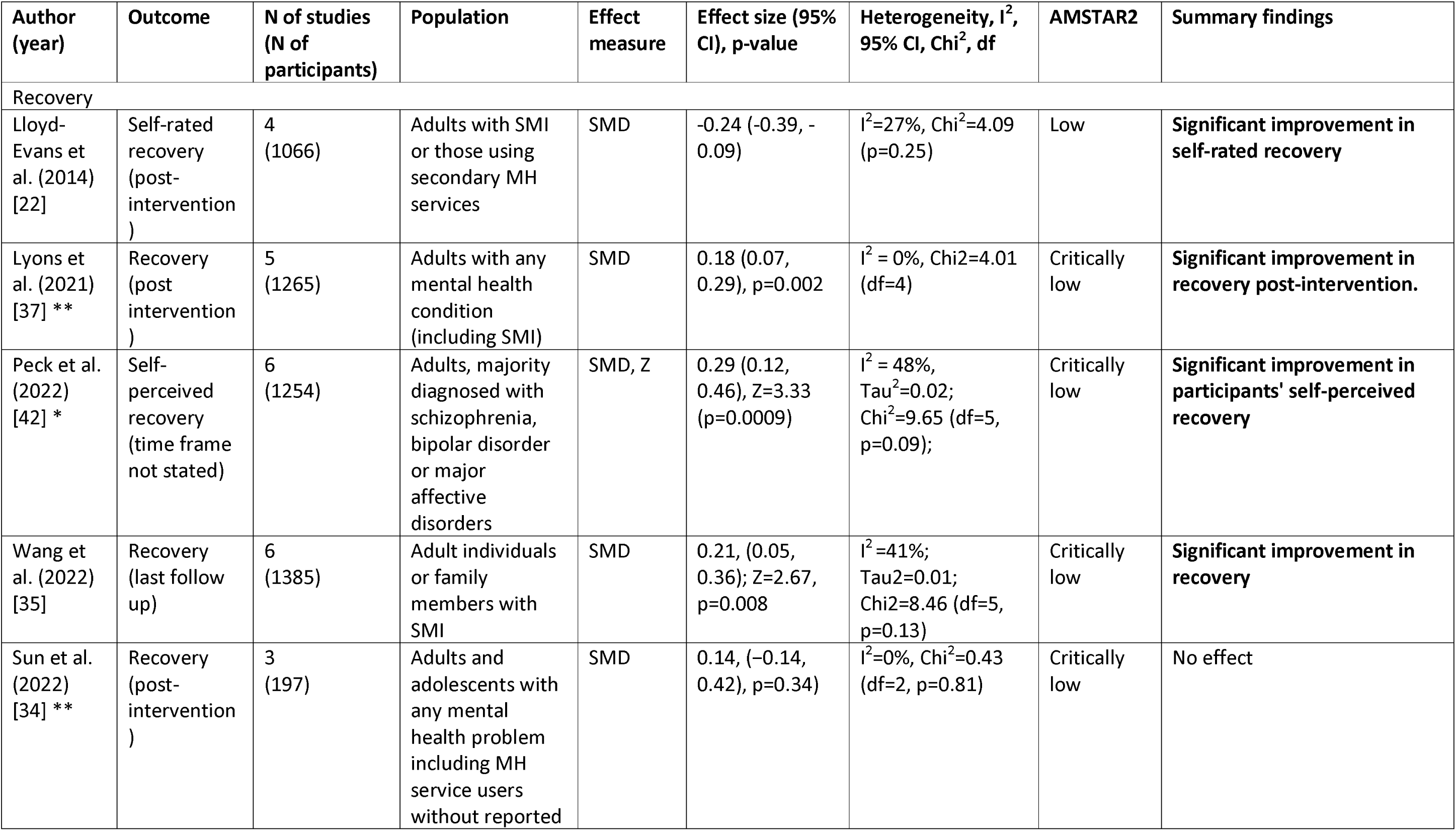
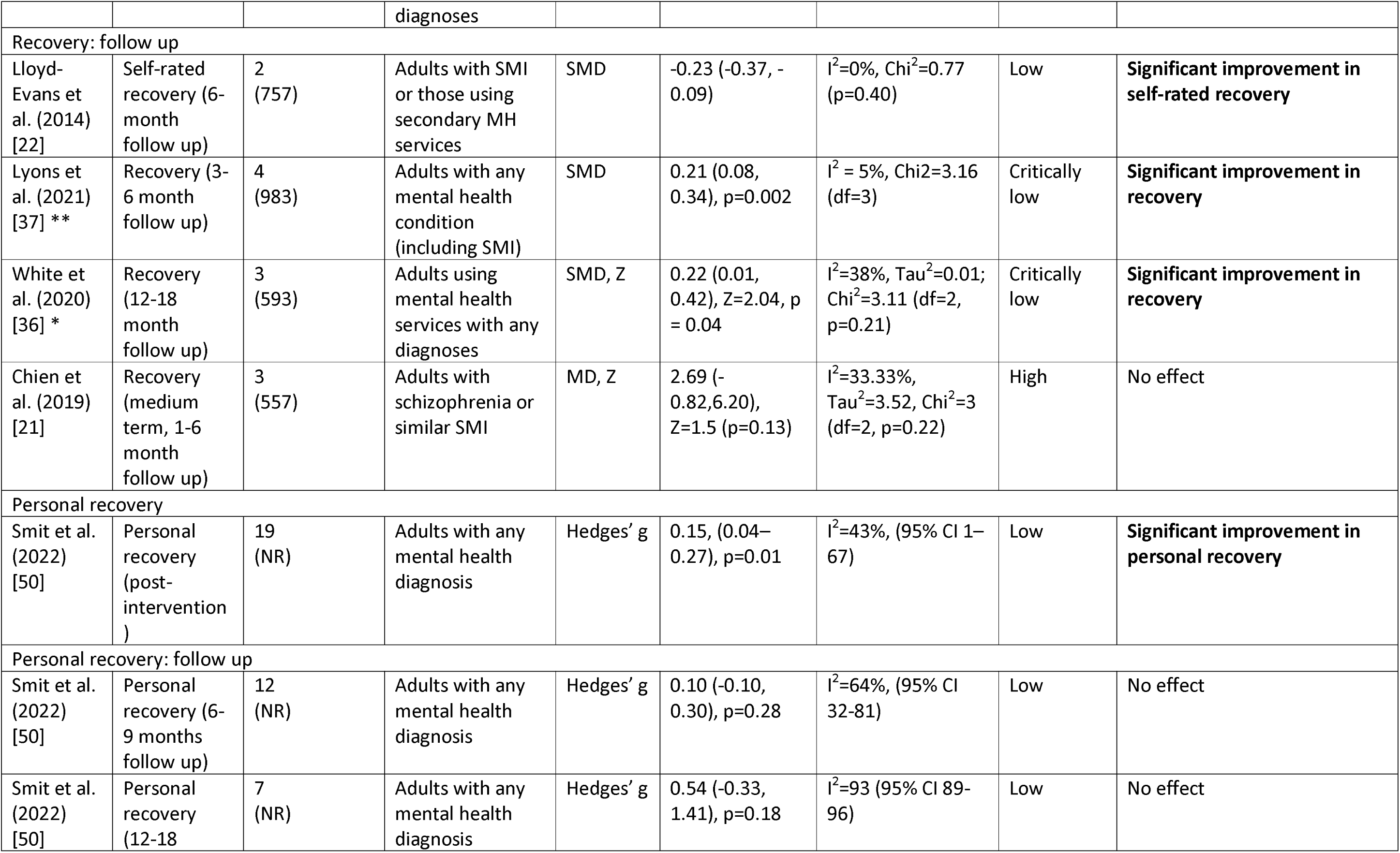
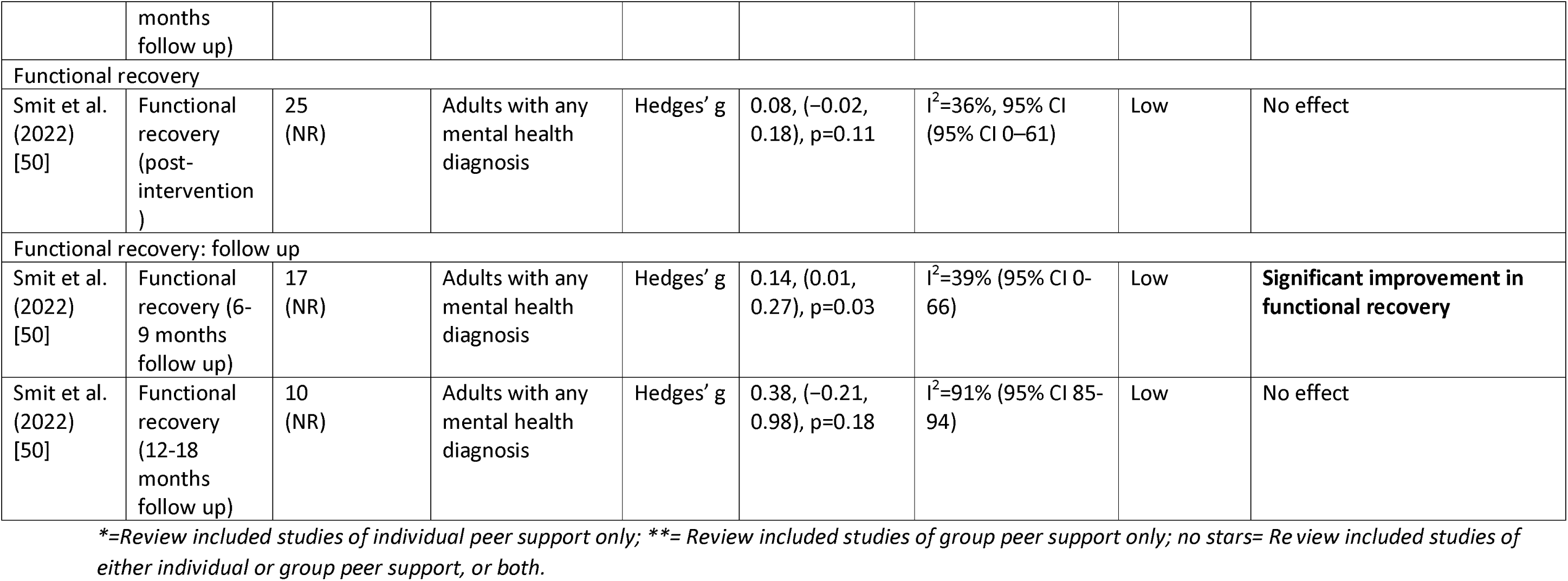
Meta-analyses effectiveness results: Recovery-related outcomes.

**Table 4:**
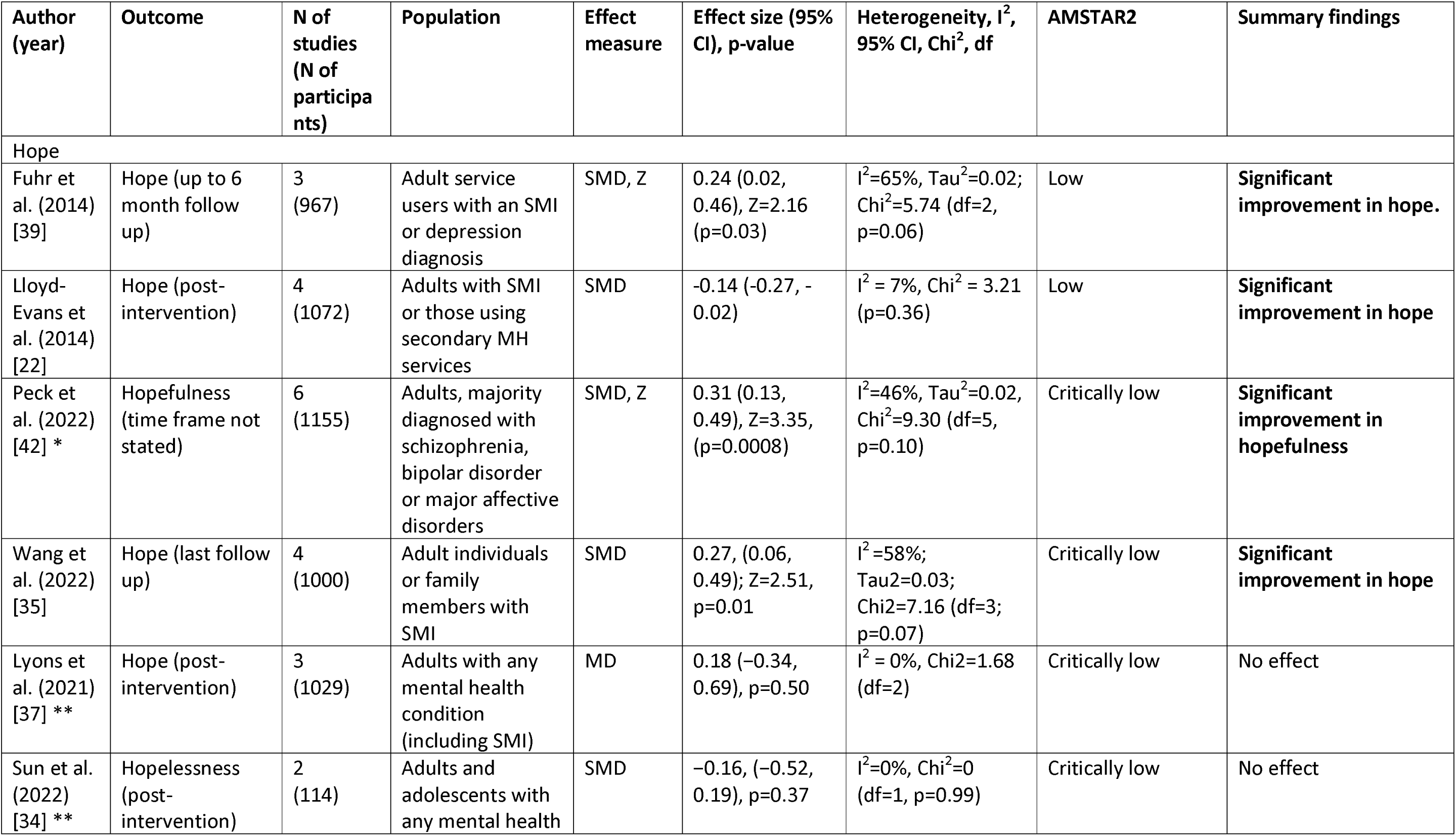
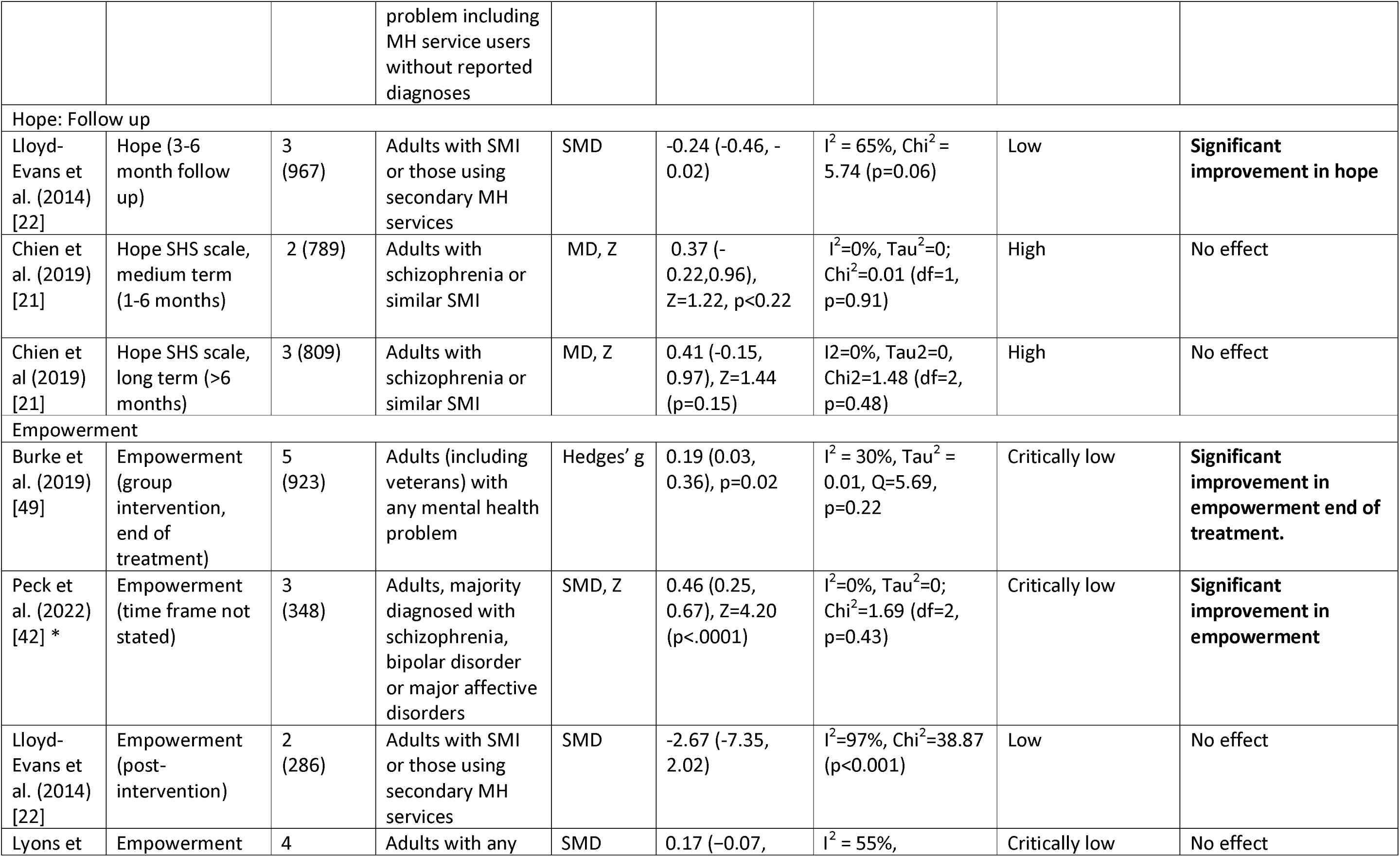
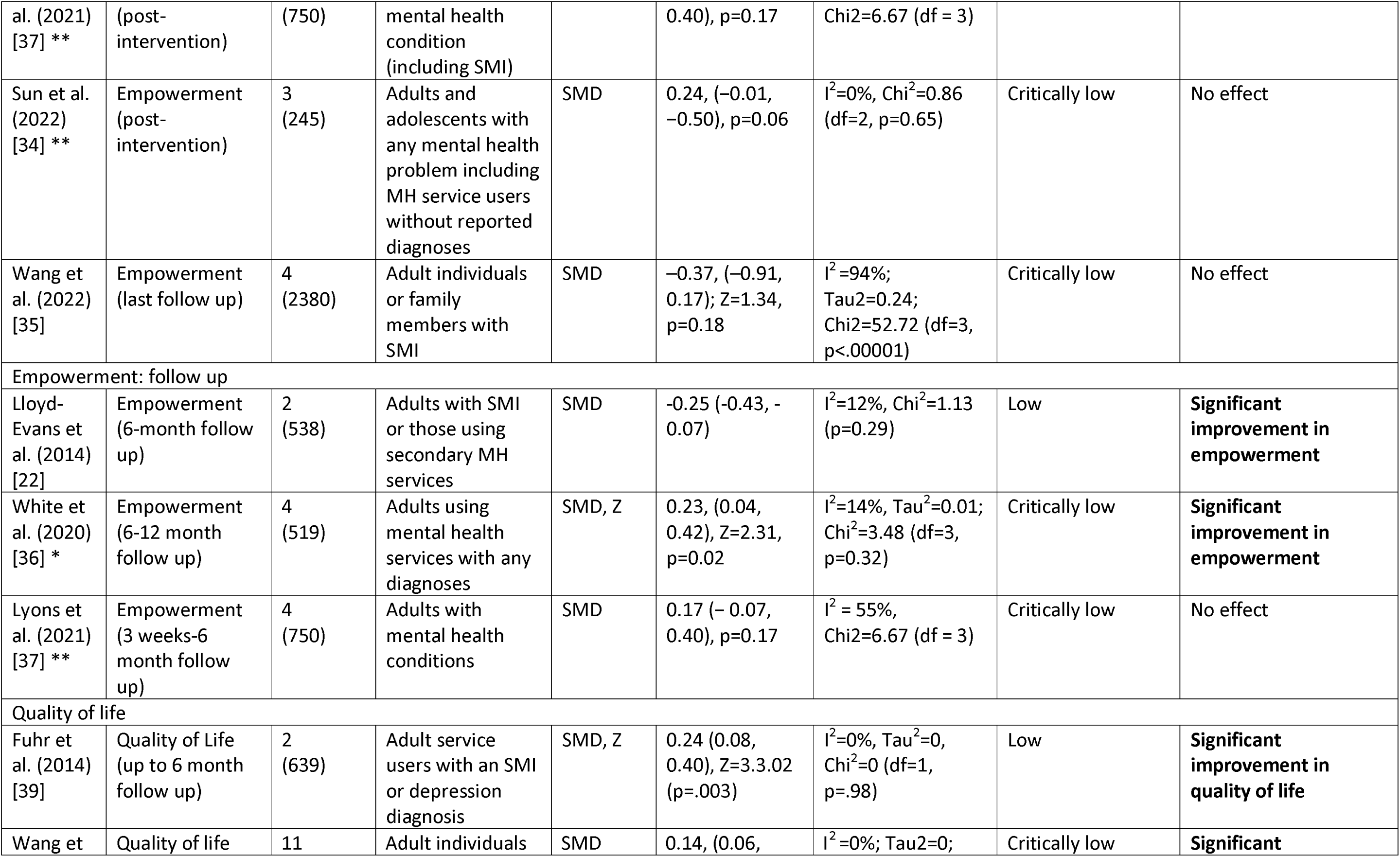
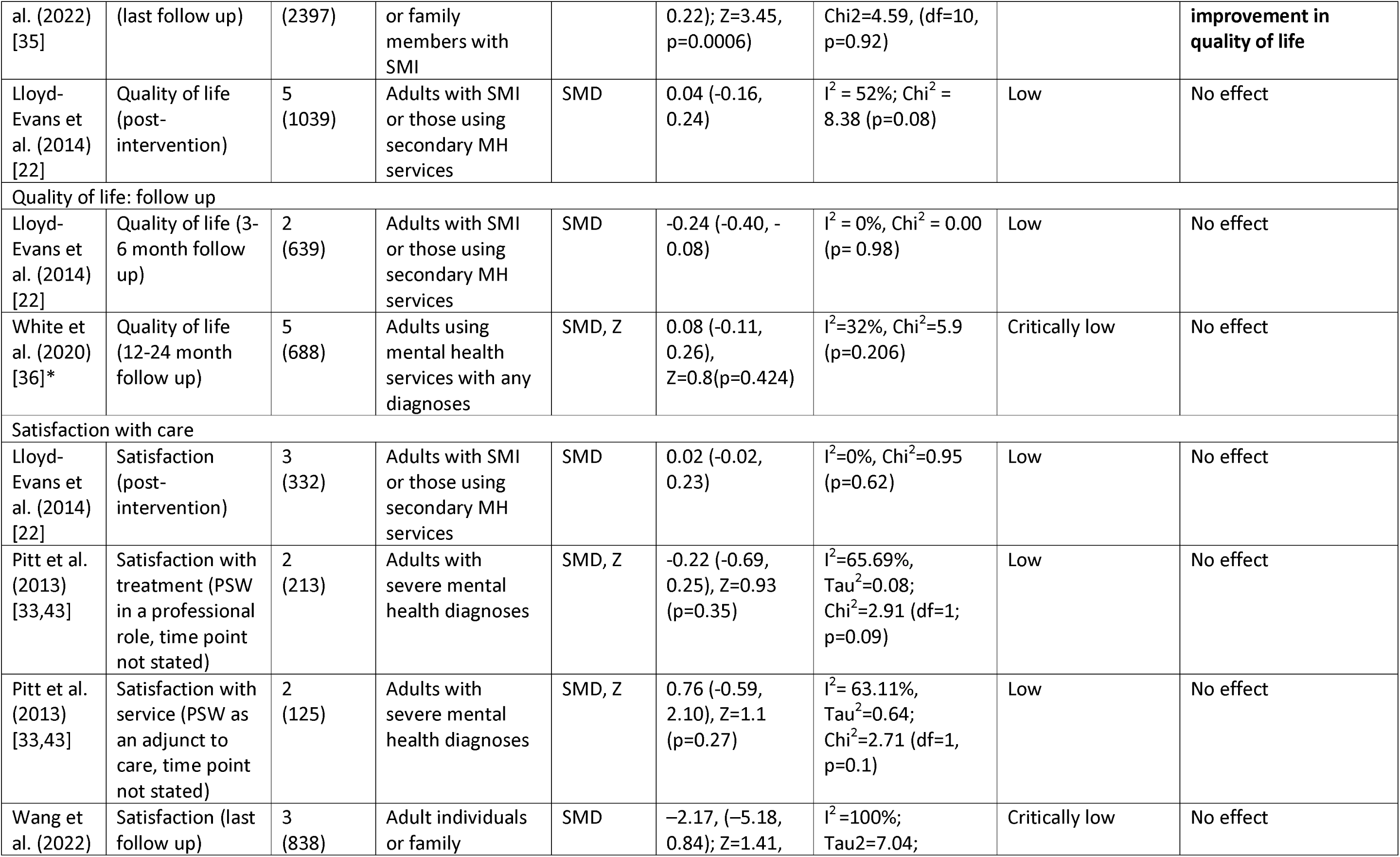
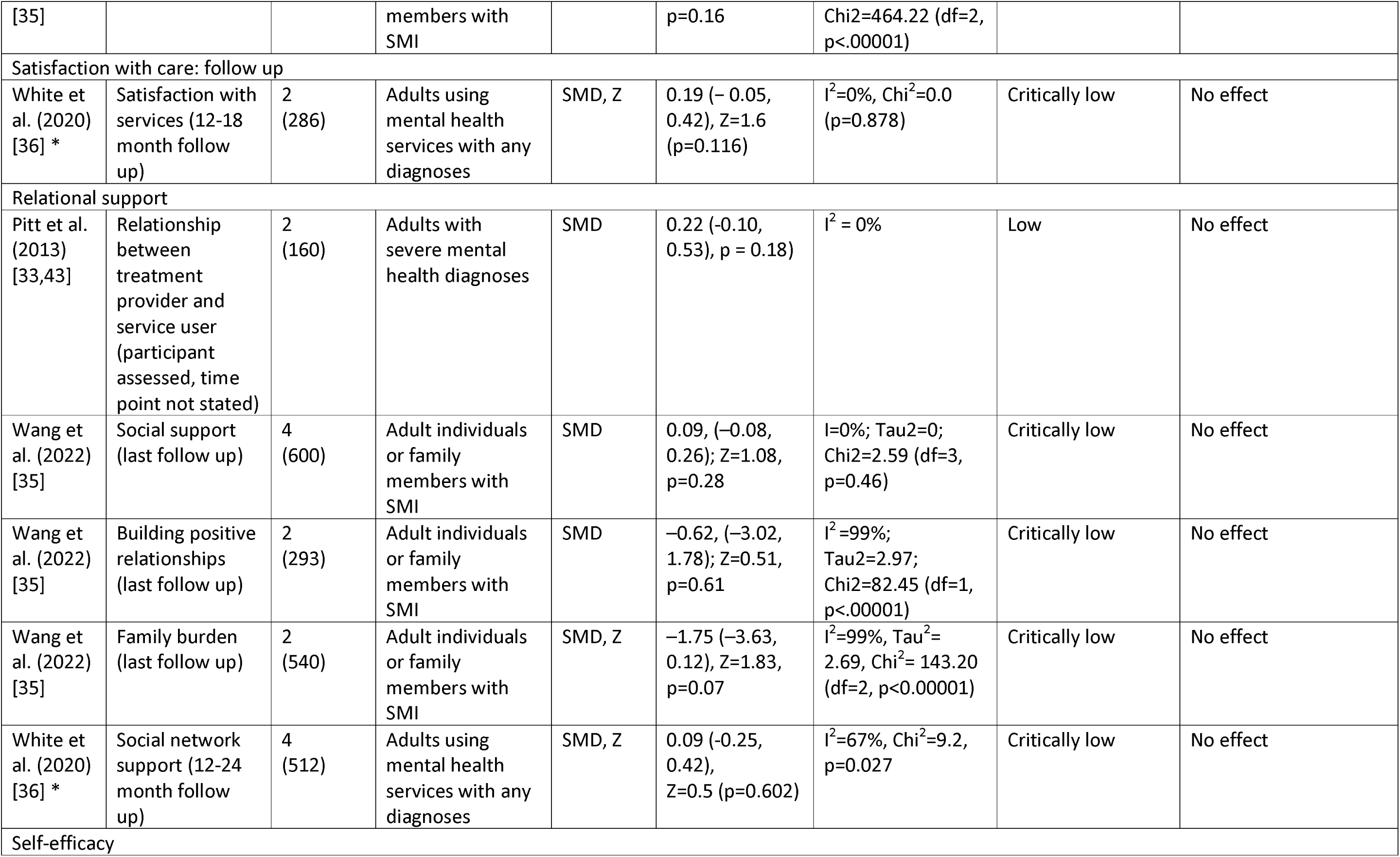
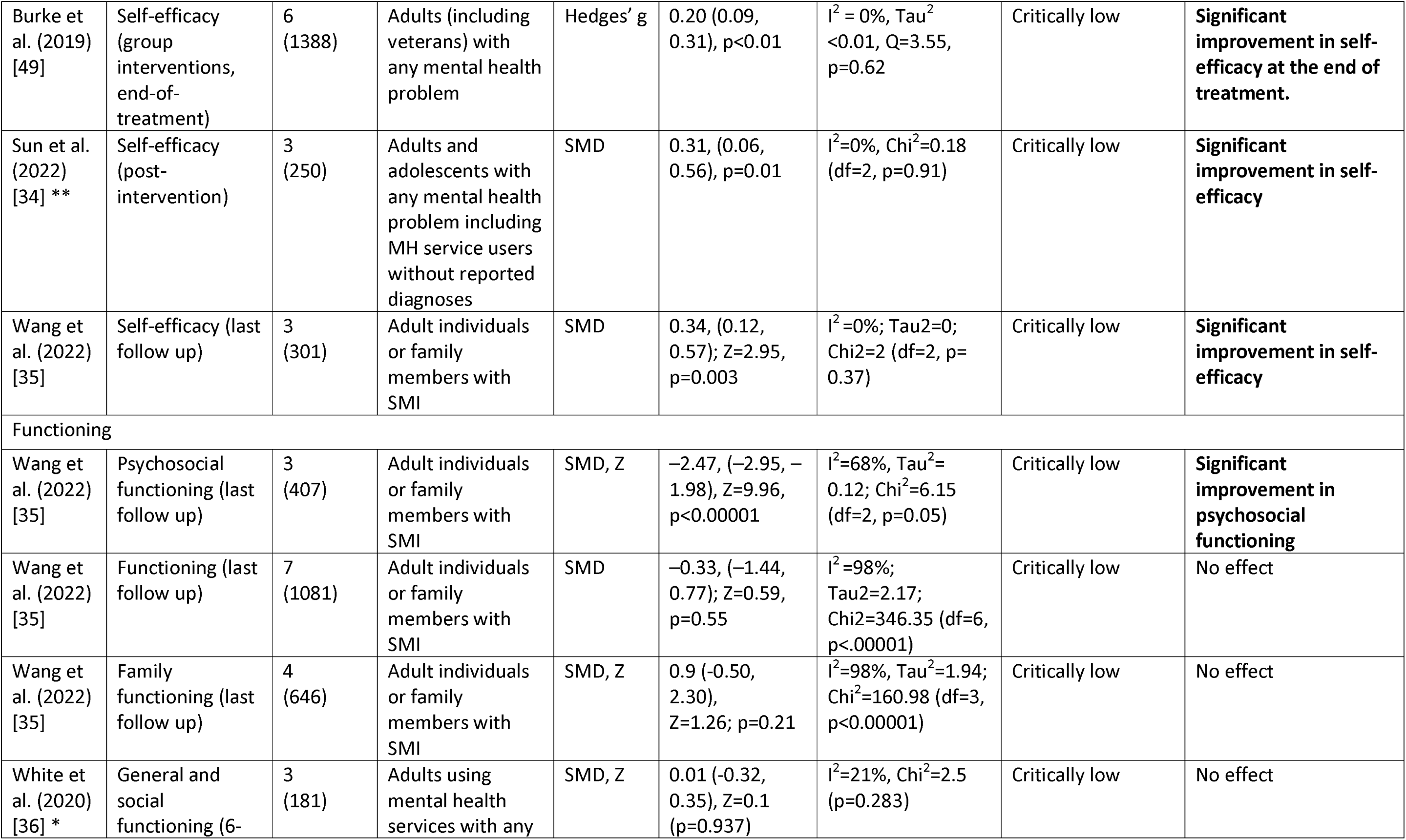
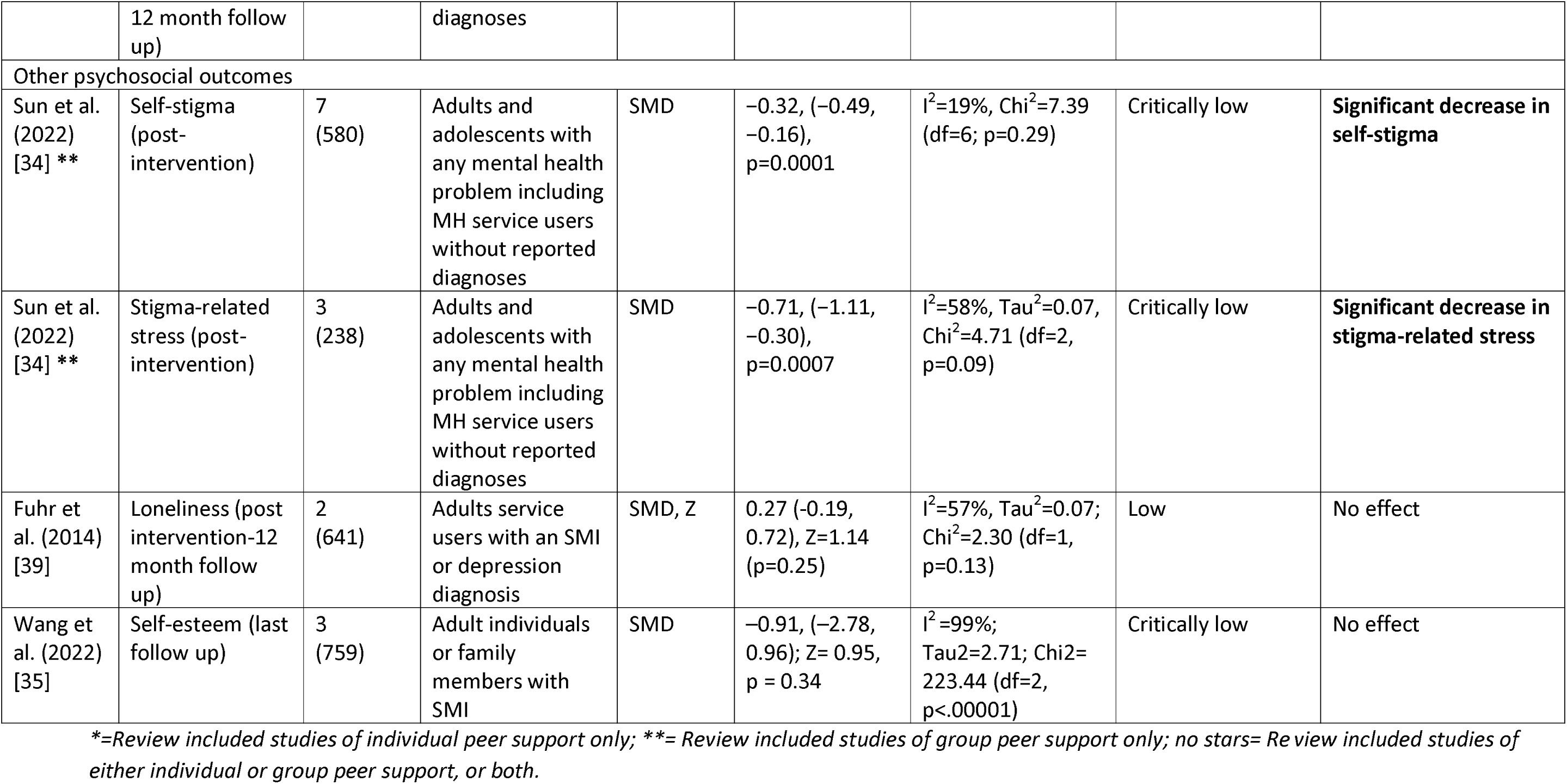
Meta-analyses effectiveness results: Psychosocial outcomes.

**Table 5:** Implementation outcomes by CFIR (Consolidated Framework for Implementation Research) domain.

| Domain | Synthesised data | Reference |
| --- | --- | --- |
| **Innovation**<br>*The “thing” being implemented, e.g., a new clinical treatment, educational program, or city service.* | - High acceptability and feasibility of PSW-led support.<br>- Engaging the community in a co-production approach should be adopted in the design of the peer provision service. | [38,41,44,48,57] |
| **Outer Setting**<br>*The setting in which the Inner Setting exists, e.g., hospital system, school district, state. There may be multiple Outer Settings and/or multiple levels within the Outer Setting (e.g., community, system, state).* | - Integration of intervention implementation within existing healthcare systems.<br>- National policy initiatives and funding provisions for employing and retaining PSWs.<br>- PSWs having access to a wider peer network.<br>- Interference of work with social security benefits.<br>- Power hierarchies in certain broader cultural contexts.<br>- Difficulties incorporating PSWs in a medical model of mental health care.<br>- A lack of recognised certification for peer workers. | [24,41,44,48,60] |
| **Inner Setting**<br>*The setting in which the innovation is implemented, e.g., hospital, school, city. There may be multiple Inner Settings and/or multiple levels within the Inner Setting, e.g., unit, classroom, team.* | - Strong leadership and support from leadership at the highest level.<br>- Importance of a workplace culture emphasising recovery-orientated practice.<br>- Employers being flexible and understanding of needs of PSWs.<br>- A supportive, accepting and trusting workplace culture where PSWs occupy a central position within service network and fit in well with other staff members.<br>- Trusting culture allows management of risk in a psychologically safe space.<br>- Access to necessary resources e.g., desk space, computer, administrative data and medical records.<br>- Time pressure and high caseloads leading to not enough time with patients.<br>- Not enough funding for PSW role and no or limited remuneration for PSWs.<br>- Effective communication and collaboration between PSWs and other workers.<br>- Organisational openness and readiness to employ PSWs.<br>- Organisations encouraging a “keeping well at workplan” to support their PSWs, especially in times of crisis. | [23,24,41,44,48,57,60] |
| **Individuals**<br>*The roles and characteristics of individuals.* | - Professionalisation and legitimisation of PSW role with performance standards/code of ethics.<br>- The use of rigorous recruitment practices to hire PSWs.<br>- High levels of competency amongst peer-counsellors when delivering interventions and having relevant skills and knowledge e.g., mental health conditions.<br>- Conflicted sense of identity when constructing either 'professional identity' or 'peer worker identity'.<br>- Required recovery status for peer supporters.<br>- PSWs ability to use coping skills and be resilient to avoid potential negative impacts on their wellbeing.<br>- Staff willingness and ability to work with PSWs and accepting them as part of the service.<br>- The use of champions and implementation leaders to drive the set up and maintenance of PSW interventions.<br>- The use of appropriate confidentiality considerations (e.g., removing PSWs details from the service if they had previously been a patient there). | [24,41,44,48,53,60] |
| **Implementation Process**<br>*The activities and strategies used to implement the innovation.* | - Comprehensive training for PSWs delivered prior to starting work and on an ongoing basis.<br>- Training should include practical skills for the PSW role, knowledge, and awareness of mental health conditions.<br>- Training other members of staff to effectively work with PSWs.<br>- Regular clinical supervision for PSWs.<br>- Clear role definition for PSW with appropriate boundaries.<br>- Safeguarding precautions e.g., removal of triggering content; psychiatric assessment and monitoring for PSWs.<br>- Establishing sustainable systems of implementation (e.g., models of cost and supervision) from the outset of the implementation process to sustain PSW engagement over time.<br>- Taking service user and PSW preferences into account when matching based on certain characteristics (e.g., demographics/diagnosis). | [24,38,41,44,48,53,57,60] |

**Table 6:** experiences of peer support (overview of themes)

| Theme | Benefit/ challenge, references | Summary and sample |
| --- | --- | --- |
| **What the PSW role can bring** |  |  |
| **Wellbeing and recovery** | Benefit<br>[23,32,59] | **PSWs** [23,32,59]: PSWs experienced improved wellness and recovery. The role enabled them to reframe and accept their illness and kept them engaged in recovery. They also experienced increased confidence, social networks, self-esteem, self-knowledge, and personal growth, through e.g., using their lived experience to help others, a sense of belonging, learning more about their own mental health and learning from service users. |
|  | Challenge<br>[33,53,58]<br>[23] | **PSW** [23,32]: the role could have a negative impact on PSW wellbeing and recovery, e.g., due to a heavy workload, the role could remind them of their illness and the 'sick' label could stay with PSWs. Service users could be a source of stress, e.g., service users who had a greater level of disturbance than the PSWs own experience.<br>**Mixed\*** [33,53,58]: PSW absenteeism due to illness or relapse increased caseload for non-peer staff. There is a risk that service users and PSWs could experience distress due to exposure to triggering content. There was fear that PSWs recovery process could negatively impact the support provided (service users, PSWs, carers, non-peer staff). |
| **Recovery and role models** | Benefit<br>[23,41,47,53,58,59,62][32] | **PSW** [23,32,53]: PSWs felt mutual benefits from the role. The role aided PSWs personal recovery through, e.g., providing a route back into employment and social inclusion. The importance of PSWs being role models was related to embodying personal recovery so they could be 'the evidence of recovery'.<br>**Service users** [41,53,59]: For service users, PSWs could be role models and give service users hope of recovery, e.g., from working with PSWs, service users experienced increased hope, motivation, better social communication skills, a sense of belonging and improved mental health symptoms. PSWs could show service users that life beyond illness is possible. Service users valued PSWs sharing their knowledge and felt empowered as they gained knowledge on mental health. Gaining knowledge motivated service users to be optimistic and independent in their recovery.<br>**Non-peer staff** [59]: From working with PSWs, non-peer staff developed increased empathy toward people in recovery and a belief in recovery.<br>**Peer-support group members** [47]: Forming relationships in peer support groups was valuable for recovery, e.g., enabled re-evaluation of self and expectations [of motherhood].<br>**Mixed\*** [58,62]: PSWs are role models, give service users hope of recovery, are valued and provide guidance and support to service users through the process of engaging with mental health services, e.g., how to |
|  |  | navigate services. (non-peer staff, PSWs, service users, policy makers, peer programme developers, carers) |
|  | Challenge [59] | **Service users** [59]: Some reported that PSWs are not role models for service users. Reasons included a belief that without formal training and because of their mental health diagnosis PSWs would be ineffective helpers. |
| **Career, social inclusion and identity** | Benefit [23,32,33,59] | **PSWs** [23,32,59]: The PSW role enabled them to contribute through work, which helped maintain recovery. The role offered a route back into employment, gaining skills, financial freedom, structure and stability, improving functioning and increasing social inclusion (e.g., by interacting with non-peer staff, on an equal footing), and social networks PSWs reported increased self-acceptance as they no longer had to hide their mental health issues. The role could also be a steppingstone into further employment.<br>**Mixed\*** [33,59]: PSW roles were rewarding and enabled service users to find a place in the community beyond 'patient'. (Mental health organisations, PSWs, non-peer staff, service users, carers) |
| **Experiential knowledge, normalisation and stigma** | Benefit [41,53,59] | **Service users** [41,53]: For service users, PSW support differed from formal treatment, it normalised and de-medicalised service user experiences. This difference felt person centred leading service users to reconnect with 'real life' situations, e.g., rebuilding relationships. Lack of judgement from PSWs reduced stigma around service users' experiences of an eating disorder. The sense of a 'shared experience' helped service users feel they were 'getting back to normal'. Service users valued peer support services and appreciated PSWs experiential knowledge, perceiving them to be more insightful than non-peer staff as they were viewed as role models in recovery, promoting empowerment and hope for service users. PSW services were trusted, making service users feel comfortable and accepted when attending activities.<br>**Mental health organisations** [59]: For organisations, PSW roles decreased mental health stigma and set a positive example to other sectors. |
|  | Challenge [41] | **Service users** [41]: Some service users and members of the public found it challenging to view PSWs as mental health professionals due to concerns on their mental health history. Some service users perceived the knowledge of PSWs to be of lower value than that by healthcare professionals and should not be fully trusted. |
| **Isolation and validation** | Benefit [47] | **Peer support group members** [47]: Having their experiences, e.g., that mothering in illness is difficult, validated by other mothers made life 'less difficult'. |
|  | Challenge [47] | **Peer support group members** [47]: Meeting other mothers could lead to increased isolation, where their experiences were contrasting, e.g., feeling that others are happy when they are not. |
| **Rapport and empathy with service users** | Benefit | **Service users** [41,59]: Service users built rapport easier with PSW than non-peer staff due to PSWs having less professional distance and being 'street smart'. Service users felt that PSWs were more approachable and |
|  | [41,59] | caring than non-peer staff, enabling them to open up and share concerns. Service users perceived greater empathy from PSWs, especially regarding adverse effects from medications |
| **Bridge** | Benefit [62] | **Mixed\*** [62]: PSWs function as a bridge between service users and non-peer staff and within the organisation, by building trust-based pathways, supporting the service user across the fragmented care system. (non-peer staff, PSWs, service users, policy makers, peer programme developers) |
| **Pioneer and expectations** | Challenge [62] | **Mixed\*** [62]: PSWs were pioneers which led to expectations and pressure, i.e., no room for failure which would reduce future PSW opportunities. (non-peer staff, PSWs, service users, policy makers, peer programme developers) |
| **Complementary role, expertise and becoming part of the team** | Benefit [62] | **Mixed\*** [62]: Non-peer staff recognised the valuable contribution of PSWs and PSWs fit with various perspectives, becoming a team member. E.g., they provided psychosocial support, were sources of experiences, fresh insights, and information, and had time to do tasks that others may not, e.g., time to just talk to patients. Collaborating with PSWs could improve recovery-oriented care. PSWs may acquire different knowledge about service users than non-peer staff, e.g., about drug abuse. (non-peer staff, PSWs, service users, policy makers, peer programme developers) |
|  | Challenge [62] | **Mixed\*** [62]: PSWs may lack a broader perspective on mental health beyond their own experience. (non-peer staff, PSWs, service users, policy makers, peer programme developers) |
| **Confusion over the PSW role** |  |  |
| **Role ambiguity** | Benefit [62] | **Mixed\*** [62]: When PSWs were introduced, their role was ambiguous. This was positive as it gave flexibility to define the role (non-peer staff, PSWs, service users, policy makers, peer programme developers). |
|  | Challenge [32,38,42,43, 53,54] | **PSWs** [23,32]: A lack of clarity about the PSW job description meant that PSWs felt confused in their role which affected their confidence, perception of competence, with ramifications for their recovery and uncertainty in their responsibilities to service users. A lack of clarity also led PSWs to feel the role was tokenistic, and to feel uncertain about where to seek support.<br>**Service users** [41][53]: Some service users perceived a lack of clarity on the PSWs' roles: PSWs were viewed as informal staff who were replaceable, leading to negative perceptions of the PSW services. Some service users perceived peer support to be tokenistic, which led to the content of the PSW intervention 'feeling irrelevant'.<br>**Mixed\*** [33,61,62]: PSWs found their role ambiguous making them anxious to demonstrate their value. PSWs felt they received insufficient training and were expected to develop the role over time, this hampered service delivery, creating the perception that PSWs were tokenistic. Non-peer staff were unsure of the PSW role, leading to a lack of support from non-peer staff. (PSW, non-peer staff, service users, carers, policy |
|  |  | makers, peer programme developers) |
| **Disclosure of peer status** | Challenge<br>[41,61,62][32] | **PSW** [32,41]: PSWs differed in how comfortable they felt in disclosing their recovery story. For some PSWs sharing their story was connected to their personal recovery. Some PSWs expressed fears of being socially excluded and labelled as “mentally ill” thus would avoid sharing their experiences because they believed service users would not trust them or value their knowledge. PSWs also expressed concern about getting jobs outside of mental health due to their peer worker identity.<br>**Mixed\*** [61,62]: There was confusion over when/with whom to disclose lived experience. For example, disclosure was important to educate team on alternative views but may require discretion within professional relationships. But ‘professionalism’ may not challenge existing boundaries which could change culture. Some PSWs felt vulnerable and were reluctant to disclose but disclosure could build trust with service users and enabled PSWs to be recovery role models. (PSW, service users, policy makers, peer programme developers, non-peer staff, mental health organisations) |
| **Boundaries** | Challenge<br>[23,58,59,61] | **PSWs** [23,59]: the transition from service user to PSW and knowing where to draw the line between friend and service provider, was challenging. Working as a PSW in substance abuse could lead to disconnection from their own recovery communities due to ethical concerns when sharing in support groups, putting the PSWs recovery at risk.<br>**Mixed\*** [58,61]: whether PSWs should relate to service users as friends (seen as unprofessional) or service users. Some PSWs would not share service user information with agency staff due to concern about violating friendship. (Service users, PSWs, carers, non-peer staff) |
| **Role conflict and professionalisation** | Challenge<br>[32,59,61,62] | **PSWs** [32]: for PSWs dual identity as a service user and service provider could be a source of stress and impact on relationships and boundaries. For example, PSWs could more closely connect with service users with similar difficulties to their own but this could have an emotional impact and could be triggering for PSWs leading to a recurrence of their own mental health issues. PSWs found the dual identity particularly difficult where PSWs were working in a team that previously cared for them.<br>**Mixed\*** [59,61,62]: The transition from patient to staff is challenging. For example, non-peer staff may be concerned about the PSW becoming unwell, especially if they were previously a patient at the facility, making PSWs feel that they’re being treated like patients. PSWs can be ‘unwilling’ to give up their consumer perspective to adopt ‘professional beliefs and roles’, e.g., training was questioned as leading to professionalisation and interference with the advantage of being a PSW. (PSW, service users, policy makers, peer programme developers, non-peer staff, mental health organisations). |
| **Organisational challenges and impact** |  |  |
| **Lack of support and training** | Challenge [23,58,62] [32] | **PSW** [23,32]: PSWs experienced a lack of support and training, potentially related to unclear job descriptions. PSWs struggled to develop the skills for their roles, including to work with service users with more complex needs than their own experiences. PSWs reported their supervision felt superficial, and problems in their relationship with their supervisors, e.g., due to PSWs not feeling that they had enough autonomy.<br>**Mixed\*** [23,58,62]: It was felt that lived experience wasn't solely sufficient to work in interprofessional teams. Some PSWs were positive about certification, others felt that certification could conflict with the grassroots, user-led ethos. Supervision and support were often not offered to PSWs. Risks might arise due to PSWs lack of training and support. Organisations needed to train PSWs and non-peer staff about the value of peer support and develop/implement guidelines. (PSW, non-peer staff, service users, carers, policy makers, peer programme developers). |
| **The value of the PSW role and low pay** | Challenge [23,59,61,62][32] | **PSWs** [23,59][32]: The value of the PSW role was linked to low pay. There were concerns about low pay, few hours and working overtime without compensation. Low pay contributed to role dissatisfaction with PSWs viewing themselves as 'cheap labour'. However, some PSWs felt that they were well compensated.<br>**Mixed\*** [61,62]: PSWs received low pay. This was difficult as they wanted jobs that freed them from disability income. Low pay contributed to role dissatisfaction and suggested the job was new, not valued or unclear. PSWs felt pay correlated with legitimacy and tokenism. Reasons for low pay were hourly pay, PSW not requiring certification, stigma from non-peer staff about 'the capacity for people with mental health conditions to work'. (non-peer staff, PSWs, service users, policy makers, peer programme developers) |
| **Workload** | Challenge [62] | **Mixed\*** [62]: PSW workload could be overwhelming. This could jeopardise other staff relationships, also under pressure from their own workload. Being given so many varying tasks (e.g., household tasks, meetings) the role could lose its distinctiveness. This was added to by a lack of understanding of the PSW role. (non-peer staff, PSWs, service users, policy makers, peer programme developers) |
| Colleagues and stigma | Challenge<br>[23,41,44,59,62][32] | **PSWs** [23,59][32]: Although PSWs reported feeling accepted in their teams, some PSWs could experience negative and rejecting non-peer staff attitudes, e.g., treated as a patient, rather than a colleague, talking inappropriately or joking about people with mental health issues, PSWs not invited to social events. PSWs felt excluded, experienced tokenism and stigma, this could lead to isolation and self-stigma.<br>**Non-peer staff** [59]: There was fear that 'cheap labour' provided by PSWs might lead to less non peer staff positions.<br>**Mixed\*** [41,44,62]: PSW roles could be a threat to other professionals' roles e.g., nurses suspicious they may be replaced. Non-peer staff were uneasy about working with people they had previously treated or PSWs seeing medical records e.g., of other PSWs.<br>Concerns from healthcare professionals and policymakers over effectiveness and safety of peer-support led to a lack of support and hostility from non-peer staff. Hence PSWs were accorded less respect and fewer responsibilities, with doubts consequently cast over their credibility.<br>PSWs felt uncomfortable talking about their role due to stigma, they challenged stigma by taking on more responsibility. Hierarchies in teams undermined PSWs feeling equal in meetings, they needed to find their voice to challenge clinically dominant ways of thinking. (PSW, service users, policy makers, peer programme developers, non-peer staff, mental health organisations, unspecified (in one study)) |
| Challenges for healthcare staff/organisations | Challenge<br>[33,59] | **Mixed\*** [33,59]: Non-peer staff felt there were expectations to support, train and supervise PSWs, increasing their workload. Some staff found it challenging to have different 'providers' [PSWs] in the team.<br>Confidentiality, disclosure and increased sick time of PSWs compared to non-peer workers were issues for organizations. (Service users, PSWs, carers, non-peer staff, mental health organisations) |
| Treatment models | Challenge<br>[23] | **PSW** [23]: PSWs are part of the newer recovery model and had trouble integrating into the traditional treatment model, e.g., where doctors held majority of power and decision making for service users but spent the least time with service users. PSWs expected to contest the traditional treatment model in support of a recovery focus (e.g., by their presence or in some cases by being openly challenging), this led to friction. If organisations are not prepared for PSWs the role doesn't provide stable employment. |
| Other |  |  |
| Offering treatment choice | [58] | **Mixed\*** [58]: Service users should have opportunities to choose among PSWs as service providers. (service users, PSWs, carers, non-peer staff). |

\*Note. For 'mixed' samples the specific sample that stated the theme is unknown (e.g., PSW or non-peer staff or both).
PSW = Peer Support Worker

### Results from meta-analyses

Here we report results on clinical outcomes, recovery-related outcomes and psychosocial outcomes from systematic reviews with meta-analysis.

### Clinical outcomes

For depression outcomes, evidence from two reviews with meta-analyses suggested that peer support is effective in improving perinatal depression [45,46]. Three reviews of peer support for adults and adolescents with mental health problems including those with SMI diagnoses reported no effect on depression post-intervention [22,34,37], where two of these reviews looked at group-based peer support alone [34,37]. Two of these reviews reported follow-up results; one review of group peer support for adults with any mental health condition continued to find no effect at 3-6 months follow up [37], while the other involving adults with SMI reported improvements in depression and anxiety at 6 months follow up, despite reporting no effect at post-intervention [22]. One review [50], measured clinical recovery in adults with any mental health diagnosis, reporting improvements post-intervention and at 6-9 month follow up, but no improvement at 12-18 month follow up.

Most evidence regarding mental health symptom severity among adults and adolescents with mental health diagnoses or who were using mental health services suggested no effect [22,33–35,37,43], other than for perinatal depression as previously summarised. One review [42] of individual peer support for adults with primarily SMI diagnoses reported improvements in symptom severity, while another involving adults with SMI [35] reported symptom improvements following family-led peer support, but no improvement following individual-led peer support. Results for service use varied depending on the measure, for example, peer support was associated with reduced risk of hospitalisation [35], including after a follow up period [36], but no effect was found regarding length of stay [33,43].

All reviews providing meta-analytic evidence relevant to this question were rated low or critically low quality, except from one high quality review [21] which found no effect of peer support on patient activation between 1-6 month follow up (a person’s perceived ability to manage their illness and their approach to healthcare) in adults with schizophrenia diagnoses or similar SMI.

### Recovery-related outcomes

Of the seven reviews with meta-analyses reporting data on overall self-reported recovery, five reported improvements in recovery in adults with mental health diagnoses including SMI [22,35–37,42]. Two studies found effects for individual peer support interventions alone [36,42], and one reported an effect for group-based peer support alone [37]. Only two reviews reported no effect [21,34], where one included studies of adults with SMI in both individual and group-based peer support [21], and the other involved studies with adults and adolescents with any mental health problem in group-based peer support alone [34].

Three reviews reported follow-up data showing continued improvements for adults with mental health diagnoses including SMI at follow ups of 6 months [22], 3-6 months [37] and 12-18 months [36], the former and the latter reviewing individual and group peer support, and the second focussing on group peer support alone. One further review reported no improvements at medium-term follow up (1-6 months) [21]. One review of adults with any mental health diagnosis identified improvements in personal recovery post-intervention, but not at 6-9 or 12-18 month follow up; and found no improvements in functional recovery post-intervention or at 12-18 month follow up, but did report improvements at 6-9 month follow up [50].

All reviews providing meta-analytic evidence for these outcomes were rated as critically low or low quality, except for one [21] which was rated high quality. Based on evidence from three studies, this latter review [21] found no effect of peer support on recovery in the medium term for adults with schizophrenia diagnoses or similar SMI.

### Psychosocial outcomes

Evidence regarding hope or hopefulness was mixed. Four reviews with meta-analyses suggested that peer support resulted in improvements in adults with SMI [22,39,42], where one of these studies looked at individual peer support alone [42] and the rest included both individual and group peer support. However, three reviews of studies including SMI and mixed mental health diagnoses samples reported no effect [21,34,37], where two of these reviews focussed on group-based peer support alone [34,37]. One study [22] followed up adults with SMI and those using secondary MH services at 3-6 months and found continued improvements in hope. However, another review investigating longer-term outcomes (over 6 months) in adults with SMI found no effect [21].

Improvements in empowerment were evidenced by two reviews with meta-analyses [42,50] of studies involving adults with any mental health diagnosis including SMI. No effects were reported in four reviews [22,34,35,37]. One of the meta-analyses finding positive effects of peer support on empowerment looked at individual peer support alone [42], whereas two of the meta-analyses with no effect solely involved group-based peer support [34,37]. Three studies reported follow up data. Two showed improvements at 6 months in adults with SMI [22] and at 6-12 month follow up among adults using mental health services with any diagnoses [36]. The other showed no improvements from group-based peer support only in adults with mental health diagnoses including SMI between 3 weeks and 6 months follow up [37].

Quality of life reportedly improved in two reviews with meta-analyses [35,39] of studies involving adults with SMI, while there was no evidence of improvement in one other with an SMI sample [22]. The two studies which reported follow up data continued to find no effect [22,36].

There were improvements in self-efficacy in adults with any mental health problem in all three reviews with meta-analyses reporting this outcome [34,35,49]. Decreases in self-stigma and stigma-related stress in adults and adolescents with any mental health problem were found by one review with meta-analysis of group-based peer support [34]. There was no evidence for peer support improving satisfaction with care [22,33,35,36,43] or relational outcomes (including social support and network), and building relationships (both personally and with staff) [33,35,36,43].

All reviews providing meta-analytic evidence for these outcomes were rated as critically low or low quality, except one high quality review [21] which found no effect of peer support on hope in adults with schizophrenia diagnoses or similar SMI in the medium or long term.

### Summary of results from systematic reviews without meta-analysis

Effectiveness results from systematic reviews without meta-analyses are tabulated in full in Appendix 6. These reviews presented mixed results pertaining to clinical outcomes including depression, anxiety, eating disorder pathology, and psychosis. However, two scoping reviews reported evidence of peer support in improving suicidal ideation [55,56]. Evidence was deemed inconclusive regarding the impact of peer support on indicators of service use, where three reviews failed to find evidence for peer support [21,22,33,43], three reported mixed results [2,40,52], and one found evidence for improvements associated with peer support [38]. More consistent evidence was found indicating peer support improves outcomes related to recovery [2,35,38,40,42,51]. For most psychosocial outcomes, systematic reviews presented mixed evidence, for example different effects were found by one high quality review for empowerment, hope and self-efficacy, depending on what measures were used [21]. Despite mixed effects being reported overall for the impact of peer support on satisfaction with care, one review cited some possible associated moderating factors such as the number of conversations had between peer supporter and recipient [46]. Evidence was marginally less mixed for relational outcomes, such as strength of interpersonal relationships and sense of community, as the majority (three) of relevant reviews found evidence in support of peer support [21,40,56], although one review found this did not persist long-term [21].

### RQ 2: What influences the implementation of peer support approaches for mental health?

Implementation was investigated in nine reviews [23,24,38,41,44,48,53,57,60]. Table 3 shows an overview of implementation outcomes by CFIR domain [31]. All reviews relevant to this research question were rated as critically low quality based on the adapted AMSTAR 2 rating scale (see Appendix 5).

#### Innovation

Studies reported generally high acceptability and feasibility of PSW-led interventions [38,41,44,48]. When planning a peer-led service, co-producing the design of peer support provision with the community and stakeholders was found to be key [57].

#### Outer setting

The existence of national policy and funding provisions for employing and retaining PSWs facilitated PSW-led care [41,44,57], as did integration of interventions within existing healthcare systems [48]. However, barriers included power hierarchies [41], difficulties incorporating PSWs in medical mental health care models [24,41,44], interference of work with welfare benefits [60] and a lack of recognised PSW certification [60].

#### Inner setting

A workplace culture emphasising recovery-orientated practice [24,57], and organisational openness and readiness to employ PSWs [41], was important. Facilitators included strong leadership and support at the highest level [44], and flexible and understanding employers, especially in times of crisis [57]. A key facilitator was a supportive, accepting and trusting workplace culture where PSWs occupy a central position and fit in well with other staff members [24]. A trusting culture allowed the management of risk in a psychologically safe space [57]; Effective communication and collaboration between PSWs and other workers facilitated this [24], whilst stigmatising staff attitudes were a barrier [60]. It was easier to implement PSW in a more collaborative and less hierarchical service [57]. There were practical facilitators and barriers for PSWs also, such as access to desk space or administrative data [24,44], time restraints, high caseloads [23,24] and insufficient funding for PSW role [24,48].

#### Individuals

The professionalisation and legitimisation of the PSW role was seen as important, with associated performance standards and/or a code of ethics [24] which was linked to rigorous recruitment practices, ensuring parity in the recruitment of PSWs and other staff [44]. A further facilitator was high levels of competency amongst peer-counsellors when delivering interventions and having relevant skills and knowledge e.g., mental health conditions [56]. PSWs were often required to have recovered from their mental health difficulties [53] and be able to use their coping skills and resilience to avoid potential negative impacts on their wellbeing [24]. PSWs reported a conflicted sense of identity between being a ‘peer’ with experience of mental health problems and a ‘professional’ as a barrier to their work [60]. The use of champions and implementation leaders to drive the set up and maintenance of PSW interventions was reported as a facilitator [44], as was staff willingness and ability to work with PSWs and accept them as part of the service [24].

#### Implementation process

Studies emphasised the importance of comprehensive training for PSWs delivered both prior to starting work and on an ongoing basis, alongside regular clinical supervision [24,44,48,53] supporting the management of any problems encountered [57]. PSW roles should be clearly defined [24,60] and training should also be delivered to other members of staff to help them work effectively with PSWs [44]. Establishing sustainable models of cost and supervision from the outset was key for the longevity of PSW [48].

### RQ 3: What are the experiences of peer support approaches for mental health (e.g., of acceptability) from the perspective of peer support workers, healthcare practitioners, service users, carers?

Experiences of both the benefits and challenges of peer support were reported in 11 reviews [23,32,33,41,44,47,53,58,59,61,62] from a range of perspectives: PSWs [23,32,41,53,59], service users [41,53,59], non-peer staff [59], peer support group members [47] and mixed samples which consisted of combinations of PSWs, service users, non-peer staff, carers, mental health organisations, policy makers and peer programme developers [23,33,41,44,53,58,59,61,62]. In one review it was unclear whose perspective was being presented [44], although this review only contributed to one theme. All reviews providing evidence for this research question were rated as critically low quality based on the adapted AMSTAR 2 rating scale (see Appendix 5). We identified 3 overarching themes: i) what the PSW role can contribute, ii) confusion over the PSW role, and iii) organisational challenges and impact. Table 4 gives an overview of the overarching themes and subthemes (with more detail in Appendix 7). The following provides an overview of each overarching theme from the perspective of the different samples (i.e. PSWs, service users, mixed samples).

### What the PSW role can bring

#### Perspective of PSWs

PSWs experienced improved wellness and recovery from working in the role, reporting increased self-esteem, personal growth, and social networks [23,32,53,59]. They benefited in a variety of ways, e.g., the role provided a route back into employment, improving functioning and social inclusion, and allowed them to learn more about their own mental health [23,32]. PSWs also reported increased self-acceptance as they no longer had to hide their mental health issues [32]. The role was therefore often reported to be mutually beneficial for PSWs and service users [32,53]. PSWs felt it was important that they were role models for service users, being ‘the evidence of recovery’ [32]. However, working as a PSW could also have a negative impact on the PSWs’ wellbeing and recovery [23,32]. Reasons for this included the role reminding them of their mental health condition and the ‘sick’ label staying with them [23].

#### Perspective of service users

For service users, PSWs could be role models, giving them hope of recovery [41,53,59]. PSW support normalised and de-medicalised service user experiences [53]. Lack of judgement from PSWs reduced feelings of self-stigma for service users [53]. Service users felt empowered by and valued gaining experiential knowledge from PSWs, perceiving them to be more insightful than non-peer staff, and trusting their services [41]. Service users also built rapport more easily with PSWs than non-peer staff, feeling they were more approachable and had greater empathy than non-peer staff [41,59]. However, some service users reported that PSWs are not role models and found it challenging to view them as professionals or fully trust their knowledge, due to their lack of training and concerns about their mental health history [41,59].

#### Perspective of non-peer staff

From working with PSWs non-peer staff developed increased empathy towards service users and a belief in recovery [59].

#### Perspective of peer support group members

Forming relationships in peer support groups and having their experiences validated by others was valuable for recovery [47]. However, group members could feel isolated when other members’ experiences contrasted with their own [47].

#### Perspective of mixed samples

PSWs were perceived to be role models, providing valuable support to service users and giving them hope of recovery [58,62]. Working as a PSW could enable service users to find a role in the community, beyond the identity of being a ‘patient’ [59]. PSWs could build trust-based pathways to function as a bridge between service users and non-peer staff [62]. Within teams, working with PSWs could improve recovery-oriented care and PSWs carried out various roles, such as providing psychosocial support, advocating for service users, providing insights based on their lived experiences [62]. For mental health organisations, PSW roles decreased stigma towards mental health problems and set a positive example [59]. However, there were fears that the PSWs’ mental health condition could impact the provided support, such as increased PSW absenteeism which could increase non-peer staff caseloads and concerns that service users’ and PSWs’ could experience distress due to exposure to difficult (‘triggering’) content [33,53,58]. PSWs experienced pressure due to the perception that they were pioneers, leading to expectations, e.g. failure could reduce future PSW opportunities [62]. There was also concern that PSWs lacked mental health knowledge, beyond their own experience [62].

### Confusion over the PSW role

#### Perspective of PSWs

A lack of clarity about the PSW job description led PSWs to feel the role was undervalued and tokenistic and meant they felt confused in their role. This impacted their perception of competence which affected their recovery and led to uncertainty in their responsibilities with service users [23,32]. PSWs also found the transition from service user to PSW and knowing where to draw the line between friend and service provider to be challenging [23,59]. Linked to this, their dual identity as a service user and provider could be a source of stress. For example, it meant they could closely connect with service users who had similar difficulties to their own, but this could also be triggering and lead to a recurrence of the PSWs’ own mental health issues [32]. PSWs expressed varying views on disclosing their recovery story [32,41]. For some, sharing elements of their story was linked to their own personal recovery [32]. However, other PSWs felt fearful of disclosure, e.g., they were concerned about being labelled ‘mentally ill’ and service users not trusting them [41].

#### Perspective of service users

A lack of clarity on the PSW role could lead service users to view the role as informal, leading to negative perceptions of the PSW services. Perceptions of tokenism of peer support could lead to the content of the PSW intervention ‘feeling irrelevant’ [41].

#### Perspective of mixed samples

PSWs and non-peer staff found the PSW role to be ambiguous, e.g. the role was not clearly defined [61] and job descriptions were ‘vague’[62]. Although this gave flexibility to define the role [62], it also led to challenges. Some PSWs felt they were expected to develop the role over time and received insufficient training, which hampered service delivery and could result in perceptions that PSWs were tokenistic [33,61,62]. Uncertainty about the role also led to a lack of support from non-peer staff [61]. Relatedly, there was confusion for PSWs over when/with whom to disclose their lived experience [61,62]. Some PSWs felt vulnerable and were reluctant to disclose, but disclosure could build trust with service users, enabled PSWs to be recovery role models, and could educate non-peer staff on alternative views [61,62]. Disclosure was also felt to require discretion when fitting with professional relationships. However, ‘professionalisation’ of PSWs may not challenge the existing boundaries (e.g. traditional hospital-based boundaries which could make it difficult for the sharing of lived experience to be valuable), when challenging these boundaries could change culture [61,62]. The transition for PSWs from patient to staff was challenging, e.g., non-peer staff were concerned about the PSW becoming unwell, making PSWs feel like they’re being treated like patients [61,62]. There were issues around boundaries, including whether PSWs should relate to service users as friends or service users [61].

### Organisational challenges and impact

#### Perspective of PSWs

PSWs experienced a lack of support and training for their role, potentially related to unclear job descriptions, and insufficient supervision [23,32]. This meant that PSWs struggled to develop the skills for their roles, including to work with service users with more complex needs than their own experiences [23]. Although there were some contrasting views, PSWs were concerned that they received low pay which made them feel that they were not valued, and they perceived themselves to be ‘cheap labour’ [23,32,59]. Some PSWs felt accepted in their teams however others experienced negative and rejecting non-peer staff attitudes [23,32,59]. For example, PSWs reported not being invited to social events and being treated like patients [59]. Consequently, some PSWs felt excluded, that their roles were tokenistic and experienced self-stigma [23,32]. PSWs as part of the newer recovery model reported challenges around integrating into traditional treatment models, e.g. where doctors spent the least time with service users but held the majority of power and decision making for service users. PSWs were expected to contest the traditional treatment model in support of a recovery focus, e.g. by their presence or in some cases being openly challenging, and this clash between old and new treatment models could lead to friction [23].

#### Perspective of non-peer staff

There was a fear that the workforce could be undermined by ‘cheap labour’ provided by PSWs and may lead to fewer non-peer staff positions [59].

#### Perspective of mixed samples

PSWs often received low pay, which led to role dissatisfaction for PSWs, suggesting the job was tokenistic or the role was unclear [61,62]. One reason for low pay was due to PSWs not requiring certification (i.e. specific qualifications, which e.g. a social worker would require) [61]. Some PSWs were positive about certification but others felt it could conflict with the grassroots ethos of peer support. However, there was the view that lived experience was not solely sufficient to work in interprofessional teams [62]. Despite this, supervision and support were often not offered to PSWs leading to risks [58,62].

There were challenges in PSW relationships with non-peer staff which could lead to a lack of support and hostility from non-peer staff. Non-peer staff felt threatened that they may be replaced by PSWs [62], were uneasy about working with people they previously treated [44], were concerned about the effectiveness of peer support [41], and felt expectations to support PSWs, increasing their workload [33]. This undermined the role of PSWs, e.g., they were subsequently given fewer responsibilities [41]. For PSWs, they wanted to challenge stigma by taking on more responsibility but high, varying workloads could jeopardise relationships with non-peer staff and team hierarchies hindered their ability to challenge clinically dominant ways of thinking [62].

### Other

#### Perspective of mixed samples

A final theme was the perception that service users should be able to choose among PSWs as service providers [58]

## Discussion

### Key findings

Our umbrella review of 35 reviews explored the effectiveness, implementation, and experiences of peer support for mental health.

Effectiveness was reported in 23 reviews. Many reviews reporting effectiveness data reported no effect of peer support on a range of outcomes, mirroring the findings from other reviews [10,65] including those focusing on other types of peer support (e.g., online peer support for young people) [66]. However, there was consistent evidence from meta-analyses that peer support may improve the clinical outcomes of perinatal depression and risk of hospitalisation of adults with severe mental illness, as well as recovery outcomes, and self-efficacy and stigma-related outcomes. Mixed meta-analytic results were found for the clinical outcomes of overall psychiatric symptoms in adults with SMI, psychosis symptoms, length of hospital stay, and patient activation; and for psychosocial outcomes such as hope, empowerment, and quality of life. There was no meta-analytic evidence for improvements in relational support. Evidence from systematic reviews without meta-analysis similarly gave a mixed picture regarding psychosocial and clinical outcomes, but indicated more consistent evidence that peer support has a positive impact on recovery, suicidal ideation, and, to some degree, satisfaction with care.

Many possible sources of heterogeneity across the included reviews could contribute to the mixed findings in this study, such as low-quality methodologies, differences in the populations included, and poor specification of peer support roles or the content of interventions delivered. One important potential contributor to our mixed results is that the primary studies contributing to the included reviews often varied in the type of control groups they considered, for example studies with treatment as usual, active controls, and waitlist controls were often reviewed within the same paper. As such, it was not possible to determine whether peer support is effective in comparison to certain types of care provision but not others. In a similar vein, we could not perform subgroup analysis to determine whether specific forms of peer support are more effective on certain populations as most reviews with meta-analyses involved a combination of different formats and a range of participant groups. Nevertheless, there was some indication that differences in the format of peer support may impact its effectiveness on empowerment, as the two meta-analyses involving individual peer support alone found a positive effect on empowerment, but the two looking at group-based peer support alone did not. However, further research is needed to adequately address such questions.

Although this overview of quantitative evidence does not give unequivocal support for peer support on a variety of outcomes, the mixed results must be understood not only in the context of heterogeneity of the quantitative research conducted thus far, but with regards to the qualitative evidence documenting strong support for this intervention (as discussed in more detail below). Given that the implementation of peer support in mental health services is still rare and highly variable, many of the trials conducted thus far may have tested peer support in environments where it is not fully embedded in the organisation and culture. Indeed, peer support may have positive impacts on the operation of mental health services that have not been measured as quantitative outcomes in existing trials – such as a stronger culture of person-centred care. More consistent quantitative results demonstrating the benefit of peer support may increasingly emerge as it becomes better integrated in the mental health care system. We identified several factors reported to be important for the successful implementation of peer support, which were summarised and structured using the CFIR. These factors included adequate training and supervision for peer support workers, a recovery-oriented workplace structure, strong leadership, and a supportive and trusting workplace culture with effective collaboration between PSWs and non-peer staff. Barriers to peer support being implemented effectively included a lack of time, resources, and appropriate funding, and a lack of recognised PSW certification. Policy, research and campaign groups have advocated implementation approaches in line with these findings, for example, ImROC (implementing Recovery through Organisational Change) [15,67], who support peer support implementation globally and international competence frameworks from New Zealand [68,69], outline recovery-focus as a core principle of peer support and emphasise the importance of training and ongoing professional development; peer support practice guidelines in the USA outline the importance of and give guidelines for supervision [70]. Formalised career pathways for PSWs [71] may help to address some of the identified barriers to effective implementation of peer support work, although these are still early in their development [67].

Experiences of peer support were from a range of perspectives (e.g., PSWs, service users, non-peer staff) and were organised under three main themes. The benefits of peer support for PSWs, service users and non-peer staff were expressed in many reviews, however there were also conflicting and challenging experiences of the role. The mental health experience of PSWs was viewed as valuable, but also subject to some stigmatising views. For PSWs, the role could improve their personal wellness and recovery, providing a route back into employment and improving functioning, and provide service users with role models of recovery. The reciprocal benefits of peer support have also been highlighted as an advantage of peer support in resources developed by NHS England [20]. However, PSWs reported the ‘sick’ label stayed with them in the role, with non-peer staff at times concerned that PSWs mental health would impact their work, and some service users reported that they found it challenging to trust PSWs knowledge due to their lack of training and mental health history. A key experience, which became the core of our second theme, was the ambiguity of the PSW job description, including lack of clarity over boundaries with service users and when to disclose PSWs recovery stories. This ambiguity meant that the role was flexible, but also led to the perception that it was tokenistic and left PSWs feeling confused which impacted their own recovery. IMROC recommend the prioritisation of clear roles when implementing peer support [67]. Professional accreditation can counter the view of peer support as tokenistic, e.g., the UK Peer Support Competence Framework outlined by the Royal College of Psychiatrists [72] and the Canadian Peer support Accreditation and Certification, a national standard endorsing peer support work as a valuable career, developed in 2017 by PSWs themselves [73]. The final theme ‘organisational challenges and impact’ included experiences such as PSWs receiving inadequate support, training, and supervision, and receiving low pay, leaving them feeling undervalued. Some non-peer staff attitudes were also a reported issue; while some PSWs felt accepted within teams, others experienced negative and rejecting non-peer staff attitudes, such as being treated as patients and not being invited to staff social events. Organisations should prepare, structurally and culturally, for the introduction of PSWs in order to ensure PSW wellbeing and reduce the risk of absences due to sickness [67,74].

### Strengths and limitations

We conducted a comprehensive search of several relevant databases and identified a large number of reviews for inclusion, providing the first detailed summary of review findings relating to effectiveness, implementation and experiences of peer support. We also had consistent involvement of researchers with lived experience of mental health and peer support delivery and receipt throughout the design, data screening and extraction, analysis and synthesis, and manuscript drafting for this paper, which allowed lived experience priorities and experiences to guide our approaches to data and our decision making throughout.

We aimed to focus our review on paid peer support, however this information was underreported in the reviews, and even when reported, interventions were often grouped with peer support interventions that did not fully meet our eligibility criteria (e.g., were unpaid). We also synthesised data from studies where payment status of peer support workers was ambiguous, i.e., not reported. This limits our ability to draw firm conclusions around paid peer support specifically, as a significant portion of the data synthesised was from studies investigating unpaid or voluntary peer support. Another limitation was the lack of involvement of people with lived experience in the included reviews, with involvement reported in only one review [55]. Given the service user-led origins of peer support, future reviews should ensure involvement of people with lived experience. Most included reviews were appraised by the AMSTAR 2 as low or critically low (97%) quality with only one review appraised as high quality. Although the low quality of reviews is a limitation, we aimed to report an overview of all current evidence for peer support to inform policy makers and healthcare practitioners, therefore to maximise the evidence base we synthesised the reviews scored as ‘critically low quality’. Our ratings are also in line with a prior umbrella review of peer support which rated 87% of reviews as critically low quality and the remainder as low quality, but reported outcomes from all reviews [65].

Beyond the aforementioned limitations regarding variation in studies within each review, there is also a loss of granular detail through the umbrella review process of summarising data across reviews, which themselves contain many studies which have been summarised. The person-centred nature of peer support may mean that there are meaningful outcomes for the service user which are not easily captured in standard outcome measurement tools or recognised as clinically significant. Variation in peer support roles across studies may have contributed to the contradictions in our findings for RQ3, e.g., the challenges around PSW roles being ambiguous, but also the reported benefits of a flexible role.

A strength of our review was our broad inclusion criteria, for example, for qualitative data on experiences of peer support we reported data from the perspectives of service users, non-peer staff and PSWs. Though some data was reported separately by role, there were studies where experiences were reported together, and these perspectives were difficult to disentangle. Finally, we did not conduct a formal meta-synthesis of the qualitative experiences data, therefore some detail may have been missed.

### Implications for practice

Peer support may be effective at improving some clinical outcomes, self-efficacy, and recovery outcomes for some people and could augment the standard service range. Certain groups may benefit from peer support more than others; evidence was strongest for depression outcomes within perinatal populations, but extremely variable for other populations. Peer support may differ in effectiveness depending on population needs and characteristics. PSWs need adequate pay, clear role descriptions and guidelines (e.g., about boundaries and disclosure), ongoing training and supervision, and opportunities for progression. Attitudes about peer support held by non-peer staff may significantly support or impede the implementation and experience of PSWs, and non-peer staff may require training about PSW roles and how to work collaboratively with PSWs. Culture, hierarchical structure and staff acceptability of peer support impact implementation and experience of peer support – structural and cultural change may be required for peer support to succeed, e.g., ensuring a recovery-oriented care model is operating in the service.

### Implications for policy

Successful implementation of PSWs in healthcare settings is likely to require a coproduction approach with clearly defined PSW roles, a receptive hierarchical structure and staff, strong leadership, and appropriate training (for PSWs and staff) with clinical and/or peer supervision alongside safeguarding. Issues relating to cost, lack of time and lack of resources are key considerations for service providers aiming to implement PSW that is sustained and effective within services. Additionally, Services could benefit from clear, coproduced guidelines, outlining the steps that are most likely to lead to successful PSW implementation.

### Implications for research

Future primary and secondary research could usefully explore the differences in efficacy, implementation, and experiences in paid PSW over time as it becomes more established; an important distinction as there are likely to be differences in these outcomes as the role of PSW develops. Such studies could consider using more personalised outcome measures such as goal based outcome measurement [75]. Current PSW roles are still poorly defined and PSW content, including PSW variations (such as whether PSWs should deliver structured or more loosely structured, informal interventions, or whether interventions should vary according to need and context) need further exploration. Realist investigations around what works for whom, how and in which contexts would uncover more fine-grained detail on the specific contexts and mechanisms that explain these differences. Very few reviews included in this umbrella review reported lived-experience researcher leadership or involvement in the undertaking of the study. It is imperative for future research in this area to appropriately reflect the priorities of those who are directly involved in PSW, either as providers or as service users. As the number of PSWs increases and more formalised roles are created, positive impact may not be restricted to outcomes of those supported by PSWs, but also to the functioning of services at an organisational level [67]. Further research is needed to evaluate how teams function with and without PSWs in order to understand how they may impact experiences through changes at a system level [67].

#### Lived Experience Commentary, written by LM and KM

This study provides a useful summary of the available research on peer support. By providing an overarching review of 35 reviews including 426 available studies, the paper brings together the knowledge on a topic of growing importance and understanding of the experiences, effectiveness, and implementation of peer support. However, this evidence is limited to ‘paid peer support workers’ included in data from academic literature of systematic reviews.

The nature of an umbrella review means that the systematic reviews themselves are synthesised, limiting our ability to look at specific details in the primary studies, for example to look for evidence of lived experience involvement or co-authorship or demographics of participants. The papers within the review are likely to have originated from traditionally funded research enquiries, and an umbrella review potentially magnifies academic or clinical perspectives over user voices and interests. While this is a frustration in any mental-health related topic, this is particularly concerning in relation to peer support, with its origins in our user-led history.

The roots in user-led peer support are also overlooked when limiting the studies to paid peer support work. Although they might use the same language of mutuality and reciprocity, the two feel different. We are hesitant to suggest that we would prefer the skills and expertise of our supporters to be voluntary and unpaid; we strongly believe their expertise should be valued and funded. But there is something magical about informal peer support which can be lost when it is over-policed in bureaucratic cultures. Additionally, with studies included in the review dating back to 1979, we question how relevant these studies are in informing England’s evolving peer support landscape.

A crucial area of future research is exploring what type of peer support works best for whom and in what circumstances, and how we can deliver this. Furthermore, we need to better understand how NHS cultures can be supported to value the expertise that originates in our lived experience, including the marginalized experiences which have been disproportionately represented in mental health services.

## Data Availability

All data produced in the present study are available upon reasonable request to the authors

