## Supplement for "The effectiveness, implementation, and experiences of peer support approaches for mental health: a systematic umbrella review"

**Appendix 1:** Prisma checklist [1]

| **Section and Topic** | **Item #** | **Checklist item** | **Location where item is reported** |
| --- | --- | --- | --- |
| **TITLE** | | |  |
| Title | 1 | Identify the report as a systematic review. | Title |
| **ABSTRACT** | | |  |
| Abstract | 2 | See the PRISMA 2020 for Abstracts checklist. | Abstract |
| **INTRODUCTION** | | |  |
| Rationale | 3 | Describe the rationale for the review in the context of existing knowledge. | Introduction |
| Objectives | 4 | Provide an explicit statement of the objective(s) or question(s) the review addresses. | Introduction |
| **METHODS** | | |  |
| Eligibility criteria | 5 | Specify the inclusion and exclusion criteria for the review and how studies were grouped for the syntheses. | Eligibility criteria; Synthesis methods; Protocol |
| Information sources | 6 | Specify all databases, registers, websites, organisations, reference lists and other sources searched or consulted to identify studies. Specify the date when each source was last searched or consulted. | Information sources and search strategy |
| Search strategy | 7 | Present the full search strategies for all databases, registers and websites, including any filters and limits used. | Appendix 2 |
| Selection process | 8 | Specify the methods used to decide whether a study met the inclusion criteria of the review, including how many reviewers screened each record and each report retrieved, whether they worked independently, and if applicable, details of automation tools used in the process. | Selection process |
| Data collection process | 9 | Specify the methods used to collect data from reports, including how many reviewers collected data from each report, whether they worked independently, any processes for obtaining or confirming data from study investigators, and if applicable, details of automation tools used in the process. | Data extraction |
| Data items | 10a | List and define all outcomes for which data were sought. Specify whether all results that were compatible with each outcome domain in each study were sought (e.g. for all measures, time points, analyses), and if not, the methods used to decide which results to collect. | Eligibility criteria, Outcome measures; Synthesis methods |
|  | 10b | List and define all other variables for which data were sought (e.g. participant and intervention characteristics, funding sources). Describe any assumptions made about any missing or unclear information. | Data extraction |
| Study risk of bias assessment | 11 | Specify the methods used to assess risk of bias in the included studies, including details of the tool(s) used, how many reviewers assessed each study and whether they worked independently, and if applicable, details of automation tools used in the process. | Quality appraisal of included reviews |
| Effect measures | 12 | Specify for each outcome the effect measure(s) (e.g. risk ratio, mean difference) used in the synthesis or presentation of results. | Synthesis methods |
| Synthesis methods | 13a | Describe the processes used to decide which studies were eligible for each synthesis (e.g. tabulating the study intervention characteristics and comparing against the planned groups for each synthesis (item #5)). | Data extraction |
|  | 13b | Describe any methods required to prepare the data for presentation or synthesis, such as handling of missing summary statistics, or data conversions. | NA |
|  | 13c | Describe any methods used to tabulate or visually display results of individual studies and syntheses. | Synthesis methods |
|  | 13d | Describe any methods used to synthesize results and provide a rationale for the choice(s). If meta-analysis was performed, describe the model(s), method(s) to identify the presence and extent of statistical heterogeneity, and software package(s) used. | Synthesis methods |
|  | 13e | Describe any methods used to explore possible causes of heterogeneity among study results (e.g. subgroup analysis, meta-regression). | NA – no meta-analysis |
|  | 13f | Describe any sensitivity analyses conducted to assess robustness of the synthesized results. | NA – no meta-analysis |
| Reporting bias assessment | 14 | Describe any methods used to assess risk of bias due to missing results in a synthesis (arising from reporting biases). | Quality appraisal of included reviews |
| Certainty assessment | 15 | Describe any methods used to assess certainty (or confidence) in the body of evidence for an outcome. | NA |
| **RESULTS** | | |  |
| Study selection | 16a | Describe the results of the search and selection process, from the number of records identified in the search to the number of studies included in the review, ideally using a flow diagram. | Figure 1 |
|  | 16b | Cite studies that might appear to meet the inclusion criteria, but which were excluded, and explain why they were excluded. | Appendix 3 |
| Study characteristics | 17 | Cite each included study and present its characteristics. | Table 1 |
| Risk of bias in studies | 18 | Present assessments of risk of bias for each included study. | Appendix 5 |
| Results of individual studies | 19 | For all outcomes, present, for each study: (a) summary statistics for each group (where appropriate) and (b) an effect estimate and its precision (e.g. confidence/credible interval), ideally using structured tables or plots. | Tables 2-4 |
| Results of syntheses | 20a | For each synthesis, briefly summarise the characteristics and risk of bias among contributing studies. | Table 1; Characteristics of included reviews; Quality appraisal of included reviews |
|  | 20b | Present results of all statistical syntheses conducted. If meta-analysis was done, present for each the summary estimate and its precision (e.g. confidence/credible interval) and measures of statistical heterogeneity. If comparing groups, describe the direction of the effect. | NA – no statistical syntheses or meta-analyses |
|  | 20c | Present results of all investigations of possible causes of heterogeneity among study results. | NA – no meta-analysis |
|  | 20d | Present results of all sensitivity analyses conducted to assess the robustness of the synthesized results. | NA |
| Reporting biases | 21 | Present assessments of risk of bias due to missing results (arising from reporting biases) for each synthesis assessed. | Appendix 5 |
| Certainty of evidence | 22 | Present assessments of certainty (or confidence) in the body of evidence for each outcome assessed. | NA |
| **DISCUSSION** | | |  |
| Discussion | 23a | Provide a general interpretation of the results in the context of other evidence. | Key findings |
|  | 23b | Discuss any limitations of the evidence included in the review. | Strengths and limitations |
|  | 23c | Discuss any limitations of the review processes used. | Strengths and limitations |
|  | 23d | Discuss implications of the results for practice, policy, and future research. | Implications for practice; Implications for policy; Implications for research |
| **OTHER INFORMATION** | | |  |
| Registration and protocol | 24a | Provide registration information for the review, including register name and registration number, or state that the review was not registered. | Methods – Study design and protocol |
|  | 24b | Indicate where the review protocol can be accessed, or state that a protocol was not prepared. | Methods – Study design and protocol |
|  | 24c | Describe and explain any amendments to information provided at registration or in the protocol. | NA |
| Support | 25 | Describe sources of financial or non-financial support for the review, and the role of the funders or sponsors in the review. | Other: Funding |
| Competing interests | 26 | Declare any competing interests of review authors. | Competing interest statement |
| Availability of data, code and other materials | 27 | Report which of the following are publicly available and where they can be found: template data collection forms; data extracted from included studies; data used for all analyses; analytic code; any other materials used in the review. | Main article; Supplement |

**Appendix 2:** full search strategy

Present the full search strategies for all databases, registers and websites, including any filters and limits used.

**Embase, Medline, Psychinfo**

Limit: 2012 to current

(((peer or peer-based or "peer based" or consumer) adj (support* or worker$ or advisor$ or facilitator$ or facilitation or facilitated or provider$ or provision or specialist$ or led or employee$ or delivery or delivered or conducted or managed or directed or staff or companion or directed or service$)) or "lived experience$").ti,ab,kw.

AND

(systematic or structured or evidence or trials or studies).ti. and ((review or overview or update* or summary).ti. or review.pt.)

OR

meta-analysis.pt. or (meta-analys* or meta analys* or metaanalys* or meta synth* or meta-synth* or metasynth*).ti,ab,kf,hw.

OR

 ((systematic or meta) adj2 (analys* or review)).ti,kf. or ((systematic* or quantitativ* or methodologic*) adj5 (review* or overview*)).ti,ab,kf,sh. or (quantitativ$ adj5 synthesis$).ti,ab,kf,hw.

OR

 ("integrative research review*" or "research integration").tw. or "scoping review?".ti,kf. or (review.ti,kf,pt. and ("trials as topic" or "studies as topic").hw.) or (evidence adj3 review*).ti,ab,kf.

OR

Review.pt. and (medline or medlars or embase or pubmed or scisearch or psychinfo or psycinfo or psychlit or psyclit or cinahl or "electronic database*" or "bibliographic database*" or "computeri#ed database*" or "online database*" or pooling or pooled or "mantel haenszel" or peto or dersimonian or "der simonian" or "fixed effect" or (hand adj2 search*) or (manual* adj2 search*)).tw,hw.

AND

(mental or psychiatr* or "affective disorder$" or schizophreni* or psychosis or psychotic or depress* or anxiety or bipolar or "obsessive compulsive" or OCD or panic or GAD or "eating disorder" or anorexi* or bulimi* or "mood disorder$" or dysthymi$ or "personality disorder$" or "posttraumatic stress" or trauma or "post-traumatic stress" or PTSD or suicid* or ((substance or drug? or alcohol or cocaine or opioid or marijuana) adj2 (use* or misuse* or addict* or disorder? or abuse*))).ti,ab,kw.

**The Cochrane Database of Systematic Reviews**

Jan 2012-December 2022

(((peer or peer-based or "peer based" or consumer) adj (support* or worker$ or advisor$ or facilitator$ or facilitation or facilitated or provider$ or provision or specialist$ or led or employee$ or delivery or delivered or conducted or managed or directed or staff or companion or directed or service$)) or "lived experience$") in Title Abstract Keyword

AND

(systematic or structured or evidence or trials or studies) and ((review or overview or update* or summary) or review) OR meta-analysis or (meta-analys* or meta analys* or metaanalys* or meta synth* or meta-synth* or metasynth*)

OR

((systematic or meta) adj2 (analys* or review)) or ((systematic* or quantitativ* or methodologic*) adj5 (review* or overview*)) or (quantitativ$ adj5 synthesis$)

OR

("integrative research review*" or "research integration") or "scoping review?" or (review and ("trials as topic" or "studies as topic")) or (evidence adj3 review*)

OR

Review and (medline or medlars or embase or pubmed or scisearch or psychinfo or psycinfo or psychlit or psyclit or cinahl or "electronic database*" or "bibliographic database*" or "computeri#ed database*" or "online database*" or pooling or pooled or "mantel haenszel" or peto or dersimonian or "der simonian" or "fixed effect" or (hand adj2 search*) or (manual* adj2 search*)) in Title Abstract Keyword

AND

(mental or psychiatr* or "affective disorder$" or schizophreni* or psychosis or psychotic or depress* or anxiety or bipolar or "obsessive compulsive" or OCD or panic or GAD or "eating disorder" or anorexi* or bulimi* or "mood disorder$" or dysthymi$ or "personality disorder$" or "posttraumatic stress" or trauma or "post-traumatic stress" or PTSD or suicid* or ((substance or drug? or alcohol or cocaine or opioid or marijuana) adj2 (use* or misuse* or addict* or disorder? or abuse*))) in Title Abstract Keyword - (Word variations have been searched)

**The Campbell Collaboration**

This database enables basic searches, therefore the following key words were used:

"peer support" OR "peer based" OR "peer led"

**Appendix 3:** excluded studies following full text screening, with reasons

| **Reference** | **Reason for exclusion** |
| --- | --- |
| Ali et al (2015) [2] | Not a paid peer support approach |
| Anderson et al (2020) [3] | Wrong population |
| Cabassa et al (2017) [4] | Wrong outcome |
| Campos et al (2014) [5] | Wrong outcome |
| Charles et al (2021) [6] | Wrong outcome |
| Charles et al (2020) [7] | Wrong outcome |
| Davidson et al (2012) [8] | Not a review |
| De Ven et al (2021) [9] | Wrong population |
| Donovan (2022) [10] | Not a review |
| Du Plessis (2020) [11] | Not a review |
| Eddie et al (2019) [12] | No quality appraisal |
| Evans et al (2021) [13] | Not a published paper |
| Gillard & Holley (2018) [14] | Not a review |
| Gopalan et al (2017) [15] | Wrong outcome |
| Grant et al (2018) [16] | Not a review |
| Griffiths & Bailey (2015) [17] | Not a paid peer support approach |
| Shalaby & Agyapong (2020) [18] | Not a review |
| Happell et al (2014) [19] | Not a paid peer support approach |
| Higgins et al (2022) [20] | Not a paid peer support approach |
| Ho et al (2022) [21] | Not a paid peer support approach |
| Jordan et al (2022) [22] | Wrong outcome |
| Kelly et al (2020) [23] | Not a paid peer support approach |
| Kia et al (2021) [24] | Not a paid peer support approach |
| King et al (2018) [25] | No quality appraisal |
| King & Fazel (2021) [26] | Not a paid peer support approach |
| Leung et al (2022) [27] | Not a paid peer support approach |
| Mahlke et al (2015) [28] | Not a review |
| Marshall et al (2015) [29] | No quality appraisal |
| Mercer et al (2021) [30] | Not a paid peer support approach |
| Miler et al (2020) [31] | Not a paid peer support approach |
| Milton et al (2017) [32] | Not a review |
| Miyamoto & Sono (2012) [33] | Duplicate |
| Mowbray et al (2021) [34] | No quality appraisal |
| Mulder & de Rooy (2018) [35] | Not a paid peer support approach |
| O’Connor et al (2017) [36] | Not a review |
| Opie et al (2022) [37] | Wrong outcome |
| Price et al (2022) [38] | Not a review |
| Richard et al (2022) [39] | Not a paid peer support approach |
| Roach (2018) [40] | Not a paid peer support approach |
| Rusch & Kosters (2021) [41] | No quality appraisal |
| Sartore et al (2021) [42] | Wrong population |
| Satinsky et al (2021) [43] | Wrong outcome |
| Schmutte et al (2020) [44] | Not a published paper |
| Scholz et al (2017) [45] | Not a paid peer support approach |
| Selseng et al (2021) [46] | Not a paid peer support approach |
| Shorey & Chua (2022) [47] | Not a paid peer support approach |
| Suresh et al (2021) [48] | Wrong population |
| Tracy & Wallace (2016) [49] | No quality appraisal |
| Tse et al (2020) [50] | Not a review |
| Tweed et al (2021) [51] | Not a paid peer support approach |
| Vally & Abrahams (2016) [52] | Not a paid peer support approach |
| Yamashita et al (2022) [53] | Not a paid peer support approach |
| Yoon et al (2022) [54] | Not a paid peer support approach |
| Zwaiman et al (2022) [55] | Wrong population |
| Davidson et al (2012) [8] | Not a review |
| Lloyd-Evans et al (2014) [56] | Duplicate |

**Appendix 4:** study overlap

|  | Bailie, 2015 | Burke 2019 | Chien 2019 | Fang 2022 | Fuhr 2014 | Huang 2020 | Lloyd Evans 2014 | Lyons 2021 | Peck 2022 | Pitt 2013 a | Pitt 2013 b | Smit 2022 | Sun 2022 | Wang 2022 | White 2020 | Bassuk 2016 | Chinman 2014 | Du Plessis 2020 | *Fortuna 2020 | Gaiser 2021 | Ibrahim 2020 | Lewis & Foye, 2022 | *Miyamoto & Sono 2012 | Mutschler 2022 | Pelizzer & Wade 2022 | Reif 2014 | Triece 2022 | Vandewalle 2016 | Jones 2014 | Walker & Bryant 2013 | Akerblom & Ness 2022 | *Bowersox 2021 | Ong 2022 | Schlichthorst 2020 | Viking 2022 | Zeng & McNamara 2021 | N reviews included |
| --- | --- | --- | --- | --- | --- | --- | --- | --- | --- | --- | --- | --- | --- | --- | --- | --- | --- | --- | --- | --- | --- | --- | --- | --- | --- | --- | --- | --- | --- | --- | --- | --- | --- | --- | --- | --- | --- |
| Adamson, 2019 |  |  |  |  |  |  |  |  |  |  |  |  |  |  |  |  |  |  |  |  |  | x |  |  | X |  |  |  |  |  |  |  |  |  |  |  | 2 |
| Agrawal, Capponi, 2016 |  |  |  |  |  |  |  |  |  |  |  |  |  |  |  |  |  |  |  |  |  |  |  |  |  |  |  |  |  |  | x |  |  |  |  |  | 1 |
| Ahmad, 2020 |  |  |  |  |  |  |  |  |  |  |  |  |  |  |  |  |  |  |  |  |  |  |  |  |  |  | x |  |  |  |  |  |  |  |  |  | 1 |
| Ahmed, 2015 |  |  |  |  |  |  |  |  |  |  |  |  |  |  |  |  |  |  |  |  | x |  |  |  |  |  |  | X |  |  |  |  |  |  |  |  | 2 |
| Akena, 2020 |  |  |  |  |  |  |  |  |  |  |  |  |  |  |  |  |  |  |  |  |  |  |  |  |  |  | x |  |  |  |  |  |  |  |  |  | 1 |
| Alvarez-Jimenez, 2013 |  |  |  |  |  |  |  |  |  |  |  |  |  |  |  |  |  |  | x |  |  |  |  |  |  |  |  |  |  |  |  |  |  |  |  |  | 1 |
| Andreas, 2010 |  |  |  |  |  |  |  |  |  |  |  |  |  |  |  |  |  |  |  |  |  |  |  |  |  | x |  |  |  |  |  |  |  |  |  |  | 1 |
| Armitage, 2010 |  |  |  |  |  |  |  |  |  |  |  |  |  |  |  |  |  |  |  |  |  |  |  |  |  | x |  |  |  |  |  |  |  |  |  |  | 1 |
| Armstrong, 1995 |  |  |  |  |  |  |  |  |  |  |  |  |  |  |  |  |  |  |  |  | x |  |  |  |  |  |  |  |  | x |  |  |  |  |  |  | 2 |
| Aschbrenner, 2016a |  |  |  |  |  |  |  |  |  |  |  |  |  |  |  |  |  |  | x |  |  |  |  |  |  |  |  |  |  |  |  |  |  |  |  |  | 1 |
| Aschbrenner, 2016b |  |  |  |  |  |  |  |  |  |  |  |  |  |  |  |  |  |  | x |  |  |  |  |  |  |  |  |  |  |  |  |  |  |  |  |  | 1 |
| Aschbrenner, 2015 |  |  |  |  |  |  |  |  |  |  |  |  |  |  |  |  |  |  | x |  | x |  |  |  |  |  |  |  |  |  |  |  |  |  |  |  | 2 |
| Asad, 2015 |  |  |  |  |  |  |  |  |  |  |  |  |  |  |  |  |  |  |  |  |  |  |  |  |  |  |  |  |  |  |  |  |  |  | x |  | 1 |
| Asad and Chriem 2016 |  |  |  |  |  |  |  |  |  |  |  |  |  |  |  |  |  | x |  |  | x |  |  |  |  |  |  | X |  |  |  |  |  |  | x |  | 4 |
| Atif, 2017 |  |  |  |  |  |  |  |  |  |  |  |  |  |  |  |  |  |  |  |  |  |  |  |  |  |  | x |  |  |  |  |  |  |  |  |  | 1 |
| Atif, 2019a |  |  |  |  |  |  |  |  |  |  |  |  |  |  |  |  |  |  |  |  |  |  |  |  |  |  | x |  |  |  |  |  |  |  |  |  | 1 |
| Balogun-Mwangi, 2017 |  |  |  |  |  |  |  |  |  |  |  |  |  |  |  |  |  |  |  |  | x |  |  |  |  |  |  |  |  |  |  |  |  |  |  |  | 1 |
| Barbic, Krupa, Armstrong, 2009 |  | X |  |  |  |  | X |  | X |  |  |  |  | X |  |  |  |  |  |  |  |  |  |  |  |  |  |  |  |  |  |  |  |  |  |  | 4 |
| Barker, 2014 |  |  |  |  |  |  |  |  |  |  |  |  |  |  |  |  |  |  |  |  |  |  |  |  |  |  |  |  |  |  |  | x |  |  |  |  | 1 |
| Barkway, 2012 |  |  |  |  |  |  |  |  |  |  |  |  |  |  |  |  |  |  |  |  | x |  |  |  |  |  |  |  |  |  |  |  |  |  |  |  | 1 |
| Barr, Townsend, 2020 |  |  |  |  |  |  |  |  |  |  |  |  |  |  |  |  |  |  |  |  |  |  |  |  |  |  |  |  |  |  | x |  |  |  |  |  | 1 |
| Bates, 2008 |  |  |  |  |  |  |  |  |  |  |  |  |  |  |  |  |  |  |  |  |  |  |  | x |  |  |  |  |  |  |  |  |  |  |  | x | 2 |
| Bennetts, 2013 |  |  |  |  |  |  |  |  |  |  |  |  |  |  |  |  |  | x |  |  |  |  |  |  |  |  |  |  |  |  |  |  |  |  |  |  | 1 |
| Beehler, 2014 |  |  |  |  |  |  |  |  |  |  |  |  |  |  |  |  |  |  |  |  |  |  |  |  |  |  |  |  |  |  | x |  |  |  |  |  | 1 |
| Ben-Zeev 2018 |  |  |  |  |  |  |  | X |  |  |  |  |  |  |  |  |  |  |  |  |  |  |  |  |  |  |  |  |  |  |  |  |  |  |  |  | 1 |
| Bernstein, 2005 |  |  |  |  |  |  |  |  |  |  |  |  |  |  |  | x |  |  |  |  |  |  |  |  |  | x |  |  |  |  |  |  |  |  |  |  | 2 |
| Berry, 2011 |  |  |  |  |  |  |  |  |  |  |  |  |  |  |  |  |  | x |  |  |  |  |  |  |  |  |  |  |  |  |  |  |  |  |  | x | 2 |
| Besio, 1993 |  |  |  |  |  |  |  |  |  |  |  |  |  |  |  |  |  |  |  |  |  |  |  |  |  |  |  |  |  | x |  |  |  |  |  |  | 1 |
| Beveridge, 2019 |  |  |  |  |  |  |  |  |  |  |  |  |  |  |  |  |  |  |  |  |  | x |  |  | X |  |  |  |  |  |  |  |  |  |  |  | 2 |
| Biagianti 2016 |  |  |  |  |  |  |  |  |  |  |  |  |  |  |  |  |  |  | x |  |  |  |  |  |  |  |  |  |  |  |  |  |  |  |  |  | 1 |
| Biggar & Neal, 1996 |  |  |  |  |  |  |  |  |  |  |  |  |  |  |  |  |  |  |  |  |  |  |  |  |  |  |  |  |  |  |  | x |  |  |  |  | 1 |
| Boevink, 2014 |  |  |  |  |  |  |  |  |  |  |  |  |  |  |  |  |  |  |  |  | x |  |  |  |  |  |  |  |  |  |  |  |  |  |  |  | 1 |
| Boevink, Kroon, Van Vugt, Delespaul, van Os, 2016 |  | X |  |  |  |  |  |  |  |  |  | X |  |  |  |  |  |  |  |  |  |  |  |  |  |  |  |  |  |  |  |  |  |  |  |  | 2 |
| Bocking, 2017 |  |  |  |  |  |  |  |  |  |  |  |  |  |  |  |  |  |  |  |  |  |  |  |  |  |  |  |  |  |  | x |  |  |  |  |  | 1 |
| Boisvert, 2008 |  |  |  |  |  |  |  |  |  |  |  |  |  |  |  |  |  |  |  |  |  |  |  |  |  | x |  |  |  |  |  |  |  |  |  |  | 1 |
| Bouchery, 2018 |  |  |  |  |  |  |  |  |  |  |  |  |  |  |  |  |  |  |  | x |  |  |  |  |  |  |  |  |  |  |  |  |  |  |  |  | 1 |
| Boyd, 2005 |  |  |  |  |  |  |  |  |  |  |  |  |  |  |  |  |  |  |  |  |  |  |  |  |  | x |  |  |  |  |  |  |  |  |  |  | 1 |
| Bracke, 2008 |  |  |  |  |  |  |  |  |  |  |  |  |  |  |  |  |  |  |  |  |  |  | x |  |  |  |  |  |  |  |  |  |  |  |  |  | 1 |
| Brasier, Roennfeldt, 2022 |  |  |  |  |  |  |  |  |  |  |  |  |  |  |  |  |  |  |  |  |  |  |  |  |  |  |  |  |  |  | x |  |  |  |  |  | 1 |
| Bright, 1999 |  |  |  |  |  |  |  |  |  | X |  |  |  |  |  |  |  |  |  |  |  |  |  |  |  |  |  |  |  |  |  |  |  |  |  |  | 1 |
| Burr, 2020 |  |  |  |  |  |  |  |  |  |  |  |  |  |  |  |  |  |  |  |  |  |  |  |  |  |  |  |  |  |  |  |  |  |  | x |  | 1 |
| Byrne, 2013 |  |  |  |  |  |  |  |  |  |  |  |  |  |  |  |  |  | x |  |  |  |  |  |  |  |  |  |  |  |  |  |  |  |  |  |  | 1 |
| Byrne, 2016 |  |  |  |  |  |  |  |  |  |  |  |  |  |  |  |  |  |  |  |  |  |  |  | x |  |  |  |  |  |  |  |  |  |  |  |  | 1 |
| Byrne, 2018 |  |  |  |  |  |  |  |  |  |  |  |  |  |  |  |  |  |  |  |  |  |  |  |  |  |  |  |  |  |  | x |  |  |  |  |  | 1 |
| Cabral, 2014 |  |  |  |  |  |  |  |  |  |  |  |  |  |  |  |  |  |  |  |  | x |  |  | x |  |  |  | x |  |  |  |  |  |  |  |  | 3 |
| Carandang, 2019 |  |  |  |  |  |  |  |  |  |  |  |  |  |  |  |  |  |  |  |  |  |  |  |  |  |  | x |  |  |  |  |  |  |  |  |  | 1 |
| Carandang, 2020 |  |  |  |  |  |  |  |  |  |  |  |  |  |  |  |  |  |  |  |  |  |  |  |  |  |  | X |  |  |  |  |  |  |  |  |  | 1 |
| Cardi, 2019 |  |  |  |  |  |  |  |  |  |  |  |  |  |  |  |  |  |  |  |  |  | x |  |  | x |  |  |  |  |  |  |  |  |  |  |  | 2 |
| Carlston, 2001 |  |  |  |  |  |  |  |  |  |  |  |  |  |  |  |  |  |  |  |  |  |  |  |  |  |  |  |  |  | x |  |  |  |  |  |  | 1 |
| Castellano, 2012 |  |  |  |  |  |  |  |  |  |  |  |  |  |  |  |  |  |  |  |  |  |  |  |  |  |  |  |  |  |  |  | x |  |  |  |  | 1 |
| Castellanos, 2018 |  |  |  |  |  |  |  |  |  |  |  |  |  |  |  |  |  |  |  | x |  |  |  |  |  |  |  |  |  |  | x |  |  |  |  |  | 2 |
| Castelein, Bruggeman et al. 2008 |  | X | x |  |  |  |  |  |  |  |  | X |  | X |  |  |  |  |  |  |  |  | x |  |  |  |  |  |  |  |  |  |  |  |  |  | 5 |
| Chang and Liu 2014 |  |  |  |  |  |  |  |  |  |  |  |  |  |  |  |  |  | x |  |  |  |  |  |  |  |  |  |  |  |  |  |  |  |  |  |  | 1 |
| Chen 2000 |  |  |  | x |  |  |  |  |  |  |  |  |  |  |  |  |  |  |  |  |  |  |  |  |  |  |  |  |  |  |  |  |  |  |  |  | 1 |
| Cheng & Yen, 2021 |  |  |  |  |  |  |  |  |  |  |  |  |  |  |  |  |  |  |  |  |  |  |  |  |  |  |  |  |  |  | x |  |  |  |  |  | 1 |
| Chenet, 2000 |  |  |  |  |  | x |  |  |  |  |  |  |  |  |  |  |  |  |  |  |  |  |  |  |  |  |  |  |  |  |  |  |  |  |  |  | 1 |
| Chibanda, 2014 |  |  |  | x |  |  |  |  |  |  |  |  |  |  |  |  |  |  |  |  |  |  |  |  |  |  | x |  |  |  |  |  |  |  |  |  | 2 |
| Chien et al 2008 |  |  |  |  |  |  |  |  |  |  |  |  |  | X |  |  |  |  |  |  |  |  |  |  |  |  |  |  |  |  |  |  |  |  |  |  | 1 |
| Chien & Chan, 2004 |  |  |  |  |  |  |  |  |  |  |  |  |  | X |  |  |  |  |  |  |  |  |  |  |  |  |  |  |  |  |  |  |  |  |  |  | 1 |
| Chien & Thompson, 2013 |  |  |  |  |  |  |  |  |  |  |  |  |  | X |  |  |  |  |  |  |  |  |  |  |  |  |  |  |  |  |  |  |  |  |  |  | 1 |
| Chien, 2018 |  |  |  |  |  |  |  |  |  |  |  |  |  | X |  |  |  |  |  |  |  |  |  |  |  |  |  |  |  |  |  |  |  |  |  |  | 1 |
| Chinman, 2001 |  |  |  |  |  |  |  |  |  |  |  |  |  |  |  |  |  |  |  |  |  |  | x |  |  |  |  |  |  |  |  |  |  |  |  |  | 1 |
| Chinman, 2006 |  |  |  |  |  |  |  |  |  |  |  |  |  |  |  |  |  |  |  |  | x |  |  |  |  |  |  |  |  |  |  |  |  |  |  | x | 2 |
| Chinman, 2008 |  |  |  |  |  |  |  |  |  |  |  |  |  |  |  |  |  |  |  |  | X |  |  |  |  |  |  | x |  | x |  |  |  |  |  | x | 4 |
| Chinman, 2010 |  |  |  |  |  |  |  |  |  |  |  |  |  |  |  |  |  |  |  |  | X |  |  | x |  |  |  |  |  |  |  |  |  |  |  | x | 3 |
| Chinman, 2012 |  |  |  |  |  |  |  |  |  |  |  |  |  |  |  |  |  |  |  |  | X |  |  | x |  |  |  |  |  |  |  |  |  |  |  |  | 2 |
| Chinman et al 2013 |  |  |  |  |  |  | x |  |  |  |  |  |  | X | X |  |  |  |  | x |  |  |  |  |  |  |  |  |  |  |  |  |  |  |  |  | 4 |
| Chinman et al 2015 |  | X |  |  |  |  |  |  |  |  |  |  |  |  |  |  |  |  |  |  | X |  |  |  |  |  |  |  |  |  |  |  |  |  |  |  | 2 |
| Chinman, 2018 |  |  |  |  |  |  |  |  |  |  |  |  |  |  |  |  |  |  |  | x |  |  |  |  |  |  |  |  |  |  |  |  |  |  |  |  | 1 |
| Chisholm and Petrakis, 2020 |  |  |  |  |  |  |  |  |  |  |  |  |  |  |  |  |  |  |  |  |  |  |  |  |  |  |  |  |  |  |  |  |  |  |  | x | 1 |
| Clarke, 2000 |  |  |  |  |  |  | x |  |  | X |  |  |  |  | X |  | X |  |  |  |  |  |  |  |  |  |  |  |  |  |  |  |  |  |  |  | 4 |
| Clarke, 2004 |  |  |  |  |  |  |  |  |  |  | x |  |  |  |  |  |  |  |  |  |  |  |  |  |  |  |  |  |  |  |  |  |  |  |  |  | 1 |
| Cleary et al 2018 |  |  |  |  |  |  |  |  |  |  |  |  |  |  |  |  |  | x |  |  |  |  |  |  |  |  |  |  |  |  |  |  |  |  | x |  | 2 |
| Collins, 2016 |  |  |  |  |  |  |  |  |  |  |  |  |  |  |  |  |  |  |  |  |  |  |  |  |  |  |  |  |  |  | x |  |  |  | x |  | 2 |
| Collins, Alla, 2019 |  |  |  |  |  |  |  |  |  |  |  |  |  |  |  |  |  |  |  |  |  |  |  |  |  |  |  |  |  |  | x |  |  |  |  |  | 1 |
| Colson, 2009 |  |  |  |  |  |  |  |  |  |  |  |  |  |  |  |  |  |  |  |  |  |  |  |  |  |  |  |  |  | x |  |  |  |  |  |  | 1 |
| Conley et al 2020 |  |  |  |  |  |  |  |  |  |  |  |  | x |  |  |  |  |  |  |  |  |  |  |  |  |  |  |  |  |  |  |  |  |  |  |  | 1 |
| Conner et al. 2015 |  | X |  |  |  |  |  |  |  |  |  |  |  |  |  |  |  |  |  |  |  |  |  |  |  |  |  |  |  |  |  |  |  |  |  |  | 1 |
| Conner, 2018 |  |  |  |  |  |  |  |  |  |  |  |  |  |  |  |  |  |  |  |  | x |  |  |  |  |  |  |  |  |  |  |  |  |  |  |  | 1 |
| Cook et al. 2009 |  | X |  |  |  |  |  |  | X |  |  |  |  |  |  |  |  |  |  |  |  |  |  |  |  |  |  |  |  |  |  |  |  |  |  |  | 2 |
| Cook 2011 |  |  |  |  |  |  | x |  |  |  |  |  |  |  |  |  |  |  |  |  |  |  |  |  |  |  |  |  |  |  |  |  |  |  |  |  | 1 |
| Cook 2012a |  |  | x |  | X |  | x | x | X |  |  | X |  | X |  |  | X |  |  |  |  |  |  |  |  |  |  |  |  |  |  |  |  |  |  |  | 8 |
| Cook 2012b |  |  | x |  | x |  |  | x | X |  |  | X |  | X |  |  | X |  |  |  |  |  |  |  |  |  |  |  |  |  |  |  |  |  |  |  | 7 |
| Cook 2012c |  |  |  |  |  |  |  |  | X |  |  |  |  | X |  |  | X |  |  |  |  |  |  |  |  |  |  |  |  |  |  |  |  |  |  |  | 3 |
| Cook 2013 |  |  |  |  |  |  |  |  | X |  |  |  |  |  |  |  |  |  |  |  |  |  |  |  |  |  |  |  |  |  |  |  |  |  |  |  | 1 |
| Corrigan, 2006 |  |  |  |  |  |  |  |  |  |  |  |  |  |  |  |  |  |  |  |  |  |  | x |  |  |  |  |  |  |  |  |  |  |  |  |  | 1 |
| Corrigan 2015 |  |  |  |  |  |  |  | x |  |  |  |  | x |  |  |  |  |  |  |  |  |  |  |  |  |  |  |  |  |  |  |  |  |  |  |  | 2 |
| Corrigan 2017 |  |  |  |  |  |  |  |  |  |  |  | x |  |  |  |  |  |  |  |  |  |  |  |  |  |  |  |  |  |  | x |  |  |  |  |  | 2 |
| Corrigan 2018 |  |  |  |  |  |  |  |  |  |  |  | X |  |  |  |  |  |  |  | x |  |  |  |  |  |  |  |  |  |  |  |  |  |  |  |  | 2 |
| Coulthard, 2013 |  |  |  |  |  |  |  |  |  |  |  |  |  |  |  |  |  |  |  |  | x |  |  |  |  |  |  |  |  |  |  |  |  |  |  |  | 1 |
| Craig 2004 |  |  |  |  |  |  | X |  |  | x | X | X |  |  | X |  | X |  |  |  |  |  |  |  |  |  |  |  |  |  |  |  |  |  |  |  | 6 |
| Crisanti, 2019 |  |  |  |  |  |  |  |  |  |  |  |  |  |  |  |  |  |  |  | x |  |  |  |  |  |  |  |  |  |  | x |  |  |  |  |  | 2 |
| Croft, Isvan, 2015 |  |  |  |  |  |  |  |  |  |  |  |  |  |  |  |  |  |  |  | x |  |  |  |  |  |  |  |  |  |  |  |  |  |  |  |  | 1 |
| Daigle, 2007 |  |  |  |  |  |  |  |  |  |  |  |  |  |  |  |  |  |  |  |  |  |  |  |  |  |  |  |  |  |  |  | x |  |  |  |  | 1 |
| Davidow, 2018 |  |  |  |  |  |  |  |  |  |  |  |  |  |  |  |  |  |  |  |  |  |  |  |  |  |  |  |  |  |  |  |  |  | x |  |  | 1 |
| Davidson, 2004 |  |  |  |  | x |  | x |  |  |  |  | X |  | X |  |  | X |  |  |  |  |  |  |  |  |  |  |  |  |  |  |  |  |  |  |  | 5 |
| Davidson, 2012 |  |  |  |  |  |  |  |  |  |  |  |  |  |  |  |  |  |  |  |  |  |  |  |  |  |  |  |  |  |  |  |  |  |  |  | x | 1 |
| Davies, Gray, 2014 |  |  |  |  |  |  |  |  |  |  |  |  |  |  |  |  |  |  |  |  |  |  |  |  |  |  |  |  |  |  | x |  |  |  |  |  | 1 |
| Davison, 2001 |  |  |  |  |  |  |  |  |  |  |  |  |  |  |  |  |  |  |  |  |  |  |  |  |  |  |  |  |  | x |  |  |  |  |  |  | 1 |
| Dawson, 2018 |  |  |  |  |  |  |  |  |  |  |  |  |  |  |  |  |  |  |  |  |  | x |  |  |  |  |  |  |  |  |  |  |  |  |  |  | 1 |
| De Man, 1991 |  |  |  |  |  |  |  |  |  |  |  |  |  |  |  |  |  |  |  |  |  |  |  |  |  |  |  |  |  |  |  | x |  |  |  |  | 1 |
| Debyser, 2018 |  |  |  |  |  |  |  |  |  |  |  |  |  |  |  |  |  |  |  |  | X |  |  |  |  |  |  |  |  |  |  |  |  |  | x |  | 2 |
| Deering, 2011 |  |  |  |  |  |  |  |  |  |  |  |  |  |  |  |  |  |  |  |  |  |  |  |  |  | x |  |  |  |  |  |  |  |  |  |  | 1 |
| Delman and Klodnick 2017 |  |  |  |  |  |  |  |  |  |  |  |  |  |  |  |  |  |  |  |  | x |  |  |  |  |  |  |  |  |  |  |  |  |  |  | x | 2 |
| Dennis 2003a |  |  |  |  |  | X |  |  |  |  |  | X |  |  |  |  |  |  |  |  |  |  |  |  |  |  |  |  |  |  |  |  |  |  |  |  | 2 |
| Dennis 2003b |  |  |  |  |  | x |  |  |  |  |  |  |  |  |  |  |  |  |  |  |  |  |  |  |  |  |  |  |  |  |  |  |  |  |  |  | 1 |
| Dennis, 2009 |  |  |  | x | x | x |  |  |  |  |  | X |  |  |  |  |  |  |  |  |  |  |  |  |  |  |  |  |  |  |  |  |  |  |  |  | 4 |
| Denison Day 2019 |  |  |  |  |  |  |  |  |  |  |  |  |  |  |  |  |  |  |  |  |  | x |  |  |  |  |  |  |  |  |  |  |  |  |  |  | 1 |
| Dhaliwal, 2009 |  |  |  |  |  |  |  |  |  |  |  |  |  |  |  |  |  |  |  |  |  |  |  |  |  |  |  |  |  |  |  | x |  |  |  |  | 1 |
| Dixon, 1994 |  |  |  |  |  |  |  |  |  |  |  |  |  |  |  |  |  |  |  |  |  |  |  |  |  |  |  |  |  | x |  |  |  |  |  |  | 1 |
| Dixon 2011 |  |  |  |  |  |  |  |  |  |  |  |  |  | X |  |  |  |  |  |  |  |  |  |  |  |  |  |  |  |  |  |  |  |  |  |  | 1 |
| Doherty, 2004 | x |  |  |  |  |  |  |  |  |  |  |  |  |  |  |  |  |  |  |  |  |  |  |  |  |  |  |  |  | x |  |  |  |  |  |  | 2 |
| Dowling, 2006 |  |  |  |  |  |  |  |  |  |  |  |  |  |  |  |  |  |  |  |  |  |  |  |  |  |  |  |  |  |  |  | x |  |  |  |  | 1 |
| Dragatsi and Alvarez, 2012 |  |  |  |  |  |  |  |  |  |  |  |  |  |  |  |  |  |  |  |  |  |  |  |  |  |  |  |  |  |  |  |  |  |  |  | x | 1 |
| Druss 2010 |  |  | x |  | x |  |  |  |  |  |  |  |  | X |  |  | X |  |  |  |  |  |  |  |  |  |  |  |  |  |  |  |  |  |  |  | 4 |
| Druss, 2018 |  |  |  |  |  |  |  |  |  |  |  |  |  |  |  |  |  |  |  | x |  |  |  |  |  |  |  |  |  |  |  |  |  |  |  |  | 1 |
| Duant, 2019 |  |  |  |  |  |  |  |  |  |  |  |  |  |  |  |  |  |  |  |  |  |  |  |  |  |  | x |  |  |  |  |  |  |  |  |  | 1 |
| Dyble et al 2014 |  |  |  |  |  |  |  |  |  |  |  |  |  |  |  |  |  | x |  |  |  |  |  |  |  |  |  |  |  |  |  |  |  |  |  |  | 1 |
| Dyble 2012 | x |  |  |  |  |  |  |  |  |  |  |  |  |  |  |  |  |  |  |  |  |  |  |  |  |  |  |  |  |  |  |  |  |  |  |  | 1 |
| Edmundson, 1982 |  |  |  |  |  |  | x |  |  |  |  |  |  |  |  |  |  |  |  |  |  |  |  |  |  |  |  |  |  |  |  |  |  |  |  |  | 1 |
| Edwards, 2018 |  |  |  |  |  |  |  |  |  |  |  |  |  |  |  |  |  |  |  |  |  |  |  |  |  |  |  |  |  |  |  |  |  | x |  |  | 1 |
| Ehrlich, 2020 |  |  |  |  |  |  |  |  |  |  |  |  |  |  |  |  |  |  |  |  |  |  |  |  |  |  |  |  |  |  |  |  |  |  | x |  | 1 |
| Eichenberg, 2006 |  |  |  |  |  |  |  |  |  |  |  |  |  |  |  |  |  |  |  |  |  |  |  |  |  |  |  |  |  |  |  |  |  | x |  |  | 1 |
| Eichenberg, 2017 |  |  |  |  |  |  |  |  |  |  |  |  |  |  |  |  |  |  |  |  |  |  |  |  |  |  |  |  |  |  |  | x |  |  |  |  | 1 |
| Eisen et al 2012 |  | X | x |  |  |  |  | x |  |  |  |  |  |  |  |  |  |  |  |  |  |  |  |  |  |  |  |  |  |  |  |  |  |  |  |  | 3 |
| Elias and Upton-Davis, 2015 |  |  |  |  |  |  |  |  |  |  |  |  |  |  |  |  |  |  |  |  |  |  |  |  |  |  |  |  |  |  |  |  |  |  | x |  | 1 |
| Ellison, 2020 |  |  |  |  |  |  |  |  |  |  |  |  |  |  |  |  |  |  |  | x |  |  |  |  |  |  |  |  |  |  |  |  |  |  |  |  | 1 |
| Enright & Parsons, 1976 |  |  |  |  |  |  |  |  |  |  |  |  |  |  |  |  |  |  |  |  |  |  |  |  |  |  |  |  |  |  |  | x |  |  |  |  | 1 |
| Fallin-Bennett, 2020 |  |  |  |  |  |  |  |  |  |  |  |  |  |  |  |  |  |  |  |  |  |  |  |  |  |  |  |  |  |  | x |  |  |  |  |  | 1 |
| Fan, 2018 |  |  |  |  |  |  |  |  |  |  |  |  |  |  |  |  |  |  |  |  |  |  |  |  |  |  |  |  |  |  |  |  | X |  |  |  | 1 |
| Fan, 2019 |  |  |  |  |  |  |  |  |  |  |  |  |  |  |  |  |  |  |  |  |  |  |  |  |  |  |  |  |  |  |  |  | x |  |  |  | 1 |
| Felton, 1995 |  |  |  |  |  |  |  |  |  |  |  |  |  |  |  |  |  |  |  |  |  |  | x |  |  |  |  |  |  |  |  |  |  |  |  |  | 1 |
| Field, 2013 |  |  |  | x |  |  |  |  |  |  |  | X |  |  |  |  |  |  |  |  |  |  |  |  |  |  |  |  |  |  |  |  |  |  |  |  | 2 |
| Finnerty, 2018 |  |  |  |  |  |  |  |  |  |  |  |  |  |  |  |  |  |  | X |  |  |  |  |  |  |  |  |  |  |  |  |  |  |  |  |  | 1 |
| Finnerty, 2019 |  |  |  |  |  |  |  |  |  |  |  |  |  |  |  |  |  |  | X |  |  |  |  |  |  |  |  |  |  |  |  |  |  |  |  |  | 1 |
| Finney, 2015 |  |  |  |  |  |  |  |  |  |  |  |  |  |  |  |  |  |  |  |  |  |  |  |  |  |  |  |  |  |  |  | x |  |  |  |  | 1 |
| Fisk, 2000 |  |  |  |  |  |  |  |  |  |  |  |  |  |  |  |  |  |  |  |  |  |  |  |  |  |  |  |  |  | x |  |  |  |  |  |  | 1 |
| Forchuk, 2005 |  |  |  |  | x |  |  |  |  |  |  |  |  | X |  |  |  |  |  |  |  |  |  |  |  |  |  |  |  |  |  |  |  |  |  |  | 2 |
| Fortuna, 2018 |  |  |  |  |  |  |  |  |  |  |  |  |  |  |  |  |  |  | x |  |  |  |  |  |  |  |  |  |  |  | x |  |  |  |  |  | 2 |
| Francis, 2002 |  |  |  |  |  |  |  |  |  |  |  |  |  |  |  |  |  |  |  |  | x |  |  |  |  |  |  | X |  |  |  |  |  |  |  |  | 2 |
| Franke, 2010 |  |  |  |  |  |  |  |  |  |  |  |  |  |  |  |  |  |  |  |  |  |  |  | x |  |  |  |  |  |  |  |  |  |  |  | x | 2 |
| Frost, 2011 |  |  |  |  |  |  |  |  |  |  |  |  |  |  |  |  |  |  |  |  | x |  |  |  |  |  |  |  |  |  |  |  |  |  |  |  | 1 |
| Fuhr, 2019 |  |  |  | x |  | x |  |  |  |  |  |  |  |  |  |  |  |  |  |  |  |  |  |  |  |  | x |  |  |  |  |  |  |  |  |  | 3 |
| Fukui, 2010 |  | X |  |  |  |  |  |  | X |  |  |  |  |  |  |  |  |  |  |  |  |  |  |  |  |  |  |  |  |  |  |  |  |  |  |  | 2 |
| Fukui, 2011 |  |  |  |  |  |  |  |  | X |  |  |  |  |  |  |  |  |  |  |  |  |  |  |  |  |  |  |  |  |  |  |  |  |  |  |  | 1 |
| Gagne et al 2018 |  |  |  |  |  |  |  |  |  |  |  |  |  |  |  |  |  | x |  |  |  |  |  |  |  |  |  |  |  |  |  |  |  |  |  |  | 1 |
| Galenter, 1988 |  |  |  |  |  |  |  |  |  |  |  |  |  |  |  |  |  |  |  |  |  |  | x |  |  |  |  |  |  |  |  |  |  |  |  |  | 1 |
| Gates and Akabas, 2007 |  |  |  |  |  |  |  |  |  |  |  |  |  |  |  |  |  |  |  |  |  |  |  | x |  |  |  | x |  | x |  |  |  |  |  | X | 4 |
| Gates, 2010 |  |  |  |  |  |  |  |  |  |  |  |  |  |  |  |  |  |  |  |  |  |  |  | x |  |  |  |  |  |  |  |  |  |  |  | x | 2 |
| Geffen, 2019 |  |  |  |  |  |  |  |  |  |  |  |  |  |  |  |  |  |  |  |  |  |  |  |  |  |  | x |  |  |  |  |  |  |  |  |  | 1 |
| Gjerdingen 2013 |  |  |  | x |  | X |  |  |  |  |  | x |  |  |  |  |  |  |  |  |  |  |  |  |  |  |  |  |  |  |  |  |  |  |  |  | 3 |
| Gidugu, 2015 |  |  |  |  |  |  |  |  |  |  |  |  |  |  |  |  |  |  |  |  |  |  |  |  |  |  |  |  |  |  | x |  |  |  |  |  | 1 |
| Gillard, 2013 |  |  |  |  |  |  |  |  |  |  |  |  |  |  |  |  |  |  |  |  |  |  |  |  |  |  |  | x |  |  |  |  |  |  |  | x | 2 |
| Gillard, Holley 2014a |  |  |  |  |  |  |  |  |  |  |  |  |  |  |  |  |  |  |  |  |  |  |  | x |  |  |  |  |  |  |  |  |  |  | x |  | 2 |
| Gillard, Edwards 2014b |  |  |  |  |  |  |  |  |  |  |  |  |  |  |  |  |  |  |  |  | X |  |  |  |  |  |  |  |  |  |  |  |  |  | x |  | 2 |
| Gillard, 2015 |  |  |  |  |  |  |  |  |  |  |  |  |  |  |  |  |  |  |  |  | x |  |  |  |  |  |  | x |  |  |  |  |  |  | X | X | 4 |
| Gillard, 2015 |  |  |  |  |  |  |  |  |  |  |  |  |  |  |  |  |  |  |  |  |  |  |  |  |  |  |  |  |  |  |  |  |  |  | x |  | 1 |
| Gillard, 2017 |  |  |  |  |  |  |  |  |  |  |  |  |  |  |  |  |  |  |  |  |  |  |  |  |  |  |  |  |  |  |  |  |  |  |  | X | 1 |
| Goldberg 2013 |  |  | x |  |  |  |  |  |  |  |  |  |  | X |  |  |  |  |  |  |  |  |  |  |  |  |  |  |  |  |  |  |  |  |  |  | 2 |
| Gordon, 1979 |  |  |  |  |  |  |  |  |  | x | x |  |  |  |  |  |  |  |  |  |  |  |  |  |  |  |  |  |  |  |  |  |  |  |  |  | 2 |
| Gordon and Bradstreet, 2015 |  |  |  |  |  |  |  |  |  |  |  |  |  |  |  |  |  |  |  |  |  |  |  | x |  |  |  |  |  |  |  |  |  |  |  |  | 1 |
| Grant, 2010 |  |  |  |  |  |  |  |  |  |  |  |  |  |  |  |  |  |  |  |  |  |  |  | x |  |  |  |  |  | x |  |  |  |  |  |  | 2 |
| Gray, 2016 |  |  |  |  |  |  |  |  |  |  |  |  |  |  |  |  |  |  |  |  |  |  |  |  |  |  |  |  |  |  |  |  |  |  |  | x | 1 |
| Gray, 2017 |  |  |  |  |  |  |  |  |  |  |  |  |  |  |  |  |  |  |  |  | x |  |  | x |  |  |  |  |  |  |  |  |  |  |  |  | 2 |
| Greenfield, 2008 |  |  |  |  | x |  |  |  |  |  |  |  |  | X |  |  |  |  |  |  |  |  |  |  |  |  |  |  |  |  |  |  |  |  |  |  | 2 |
| Griffiths, 2012 |  |  |  |  |  |  |  |  |  |  |  | X |  |  |  |  |  |  |  |  |  |  |  |  |  |  |  |  |  |  |  |  |  |  |  |  | 1 |
| Griffiths, Hancock-Johnson, 2017 |  |  |  |  |  |  |  |  |  |  |  |  |  |  |  |  |  |  |  |  |  |  |  |  |  |  |  |  |  |  | x |  |  |  |  |  | 1 |
| Griswold, 2010 |  |  |  |  |  |  |  |  |  |  |  |  |  |  |  |  |  |  |  |  |  |  | x |  |  |  |  |  |  |  |  |  |  |  |  |  | 1 |
| Gucci and Marmo, 2016 |  |  |  |  |  |  |  |  |  |  |  |  |  |  |  |  |  |  | x |  |  |  |  |  |  |  |  |  |  |  |  |  |  |  |  |  | 1 |
| Gulliver, 2011 |  |  |  |  |  |  |  |  |  |  |  |  |  |  |  |  |  |  | x |  |  |  |  |  |  |  |  |  |  |  |  |  |  |  |  |  | 1 |
| Ha 2016 |  |  |  |  |  |  |  |  |  |  |  |  |  |  |  |  |  | x |  |  |  |  |  |  |  |  |  |  |  |  |  |  |  |  |  |  | 1 |
| Hall & Gabor, 2004 |  |  |  |  |  |  |  |  |  |  |  |  |  |  |  |  |  |  |  |  |  |  |  |  |  |  |  |  |  |  |  | x |  |  |  |  | 1 |
| Hall, 2018 |  |  |  |  |  |  |  |  |  |  |  |  |  |  |  |  |  |  |  |  | x |  |  |  |  |  |  |  |  |  |  |  |  |  |  |  | 1 |
| Hamilton, 2015 |  |  |  |  |  |  |  |  |  |  |  |  |  |  |  |  |  |  |  |  |  |  |  | x |  |  |  | x |  |  |  |  |  |  |  |  | 2 |
| Harris, 2020 |  |  |  |  |  |  |  |  |  |  |  |  |  |  |  |  |  |  |  |  |  |  |  |  |  |  |  |  |  |  | x |  |  |  |  |  | 1 |
| Harrison, 2017 |  |  |  |  |  |  |  |  |  |  |  |  |  |  |  |  |  |  |  |  |  |  |  |  |  |  |  |  |  |  | x |  |  |  |  |  | 1 |
| Hebert, 2008 |  |  |  |  |  |  |  |  |  |  |  |  |  |  |  |  |  |  |  |  |  |  |  | x |  |  |  |  |  |  |  |  |  |  |  |  | 1 |
| Hellner, 2021 |  |  |  |  |  |  |  |  |  |  |  |  |  |  |  |  |  |  |  |  |  |  |  |  | x |  |  |  |  |  |  |  |  |  |  |  | 1 |
| Hensley and Dawson, 2017 |  |  |  |  |  |  |  |  |  |  |  |  |  |  |  |  |  |  |  |  |  |  |  |  |  |  |  |  |  |  |  |  |  |  | x |  | 1 |
| Hibbs, 2015 |  |  |  |  |  |  |  |  |  |  |  |  |  |  |  |  |  |  |  |  |  |  |  |  | x |  |  |  |  |  |  |  |  |  |  |  | 1 |
| Higgins, 2017 |  |  |  |  |  |  |  |  |  |  |  |  |  |  |  |  |  |  |  |  | x |  |  |  |  |  |  |  |  |  |  |  |  |  |  |  | 1 |
| Hodsoll 2017 |  |  |  |  |  |  |  |  |  |  |  |  |  |  |  |  |  |  |  |  |  | x |  |  |  |  |  |  |  |  |  |  |  |  |  |  | 1 |
| Holley, 2015 |  |  |  |  |  |  |  |  |  |  |  |  |  |  |  |  |  |  |  |  | x |  |  |  |  |  |  |  |  |  |  |  |  |  |  | x | 2 |
| Hundt 2015 |  |  |  |  |  |  |  |  |  |  |  |  |  |  |  |  |  |  |  |  | x |  |  |  |  |  |  |  |  |  |  |  |  |  |  |  | 1 |
| Hundert, 2021 |  |  |  |  |  |  |  |  |  |  |  |  | x |  |  |  |  |  |  |  |  |  |  |  |  |  |  |  |  |  |  |  |  |  |  |  | 1 |
| Hunkeler, 2000 |  |  |  |  |  |  |  |  |  |  |  |  |  |  | X |  |  |  |  |  |  |  |  |  |  |  |  |  |  |  |  |  |  |  |  |  | 1 |
| Hurley, 2018 |  |  |  |  |  |  |  |  |  |  |  |  |  |  |  |  |  | x |  |  |  |  |  |  |  |  |  |  |  |  |  |  |  |  |  |  | 1 |
| Hymes 2015 |  |  |  |  |  |  |  |  |  |  |  |  |  |  |  |  |  | x |  |  |  |  |  |  |  |  |  |  |  |  |  |  |  |  |  |  | 1 |
| Ja 2009 |  |  |  |  |  |  |  |  |  |  |  |  |  |  |  | x |  |  |  |  |  |  |  |  |  |  |  |  |  |  |  |  |  |  |  |  | 1 |
| Jack, Oller, 2018 |  |  |  |  |  |  |  |  |  |  |  |  |  |  |  |  |  |  |  |  |  |  |  |  |  |  |  |  |  |  | x |  |  |  |  |  | 1 |
| Jewell, Davidson, 2006 |  |  |  |  |  |  |  |  |  |  |  |  |  |  |  |  |  |  |  |  | x |  |  |  |  |  |  |  |  |  |  |  |  |  |  |  | 1 |
| John, 2018 |  |  |  |  |  |  |  |  |  |  |  |  |  |  |  |  |  |  |  |  |  |  |  |  |  |  |  |  |  |  |  |  |  | x |  |  | 1 |
| Johnson, 2018 |  |  |  |  |  |  |  |  | x |  |  | X |  |  | x |  |  |  |  |  | x |  |  |  |  |  |  |  |  |  |  |  |  |  |  |  | 4 |
| Johnson, 2021 |  |  |  |  |  |  |  |  |  |  |  |  |  |  |  |  |  |  |  |  |  |  |  |  |  |  |  |  |  |  | x |  |  |  |  |  | 1 |
| Jonikas, 2013 |  | X |  |  |  |  |  |  |  |  |  |  |  |  |  |  | X |  |  |  |  |  |  |  |  |  |  |  |  |  |  |  |  |  |  |  | 2 |
| Junker, 2005 |  |  |  |  |  |  |  |  |  |  |  |  |  |  |  |  |  |  |  |  |  |  |  |  |  |  |  |  |  |  |  | x |  |  |  |  | 1 |
| Kamalifard, 2013 |  |  |  | x |  |  |  |  |  |  |  |  |  |  |  |  |  |  |  |  |  |  |  |  |  |  |  |  |  |  |  |  |  |  |  |  | 1 |
| Kamon & Turner 2013 |  |  |  |  |  |  |  |  |  |  |  |  |  |  |  | x |  |  |  |  |  |  |  |  |  |  |  |  |  |  |  |  |  |  |  |  | 1 |
| Kandah, 2017 |  |  |  |  |  |  |  |  |  |  |  |  |  |  |  |  |  |  |  |  |  |  |  |  |  |  | x |  |  |  |  |  |  |  |  |  | 1 |
| Kaplan, 2011 |  |  |  |  |  |  | x | x |  |  |  | X |  | X |  |  |  |  | x |  |  |  |  |  |  |  |  |  |  |  |  |  |  |  |  |  | 5 |
| Kaplan 2014 |  |  |  |  |  |  |  |  |  |  |  |  |  |  |  |  |  |  | x |  |  |  |  |  |  |  |  |  |  |  |  |  |  |  |  |  | 1 |
| Kaufman, 1995 |  |  |  |  |  |  |  |  |  | x | X |  |  |  |  |  |  |  |  |  |  |  |  |  |  |  |  |  |  |  |  |  |  |  |  |  | 2 |
| Kelly 2014 |  |  | x |  |  |  |  |  |  |  |  |  |  | X |  |  |  |  |  |  |  |  |  |  |  |  |  |  |  |  |  |  |  |  |  |  | 2 |
| Kelly 2017 |  |  |  |  |  |  |  |  |  |  |  |  |  |  |  |  |  |  |  | x |  |  |  |  |  |  |  |  |  |  |  |  |  |  |  |  | 1 |
| Kemp, 2012 |  |  |  |  |  |  |  |  |  |  |  |  |  |  |  |  |  |  |  |  | x |  |  |  |  |  |  |  |  |  |  |  |  |  |  |  | 1 |
| Kemp & Henderson 2012 |  |  |  |  |  |  |  |  |  |  |  |  |  |  |  |  |  | x |  |  |  |  |  |  |  |  |  | x |  |  |  |  |  |  |  | x | 3 |
| Keogh, 2014 |  |  |  |  |  |  |  |  |  |  |  |  |  |  |  |  |  |  |  |  | x |  |  |  |  |  |  |  |  |  |  |  |  |  |  |  | 1 |
| Kermode, 2020 |  |  |  |  |  |  |  |  |  |  |  |  |  |  |  |  |  |  |  |  |  |  |  |  |  |  | x |  |  |  |  |  |  |  |  |  | 1 |
| Kermode, 2021 |  |  |  |  |  |  |  |  |  |  |  |  |  |  |  |  |  |  |  |  |  |  |  |  |  |  |  |  |  |  |  |  | x |  |  |  | 1 |
| Kidd, 2016 |  |  |  |  |  |  |  |  |  |  |  |  |  |  |  |  |  |  |  |  | x |  |  |  |  |  |  |  |  |  | x |  |  |  |  |  | 2 |
| Kidd, 2021 |  |  |  |  |  |  |  |  |  |  |  |  |  |  |  |  |  |  |  |  |  |  |  |  |  |  |  |  |  |  | x |  |  |  |  |  | 1 |
| Kido, 2017 |  |  |  |  |  |  |  |  |  |  |  |  |  |  |  |  |  |  |  |  | x |  |  |  |  |  |  |  |  |  |  |  |  |  |  |  | 1 |
| Klein, 1998 |  |  |  |  |  |  |  |  |  |  |  |  |  |  | X |  |  |  |  |  |  |  |  |  |  |  |  |  |  |  |  | x |  |  |  |  | 2 |
| Korsbek 2016 |  |  |  |  |  |  |  |  |  |  |  |  |  |  |  |  |  |  | x |  |  |  |  |  |  |  |  |  |  |  |  |  |  |  |  |  | 1 |
| Kral, 2006 |  |  |  |  |  |  |  |  |  |  |  |  |  |  |  |  |  |  |  |  |  |  |  |  |  |  |  |  |  |  |  |  |  | x |  |  | 1 |
| Kroschel and Casey, 2011 |  |  |  |  |  |  |  |  |  |  |  |  |  |  |  |  |  |  |  |  |  |  |  |  |  |  |  |  |  |  |  |  |  |  |  | x | 1 |
| Kuek, 2021 |  |  |  |  |  |  |  |  |  |  |  |  |  |  |  |  |  |  |  |  |  |  |  |  |  |  |  |  |  |  |  |  | x |  |  |  | 1 |
| Lawn, 2008 |  |  |  |  |  |  |  |  |  |  |  |  |  |  |  |  |  |  |  |  |  |  | x |  |  |  |  |  |  |  |  |  |  |  |  |  | 1 |
| Landers, Zhou, 2014 |  |  |  |  |  |  |  |  |  |  |  |  |  |  |  |  |  |  |  | x |  |  |  |  |  |  |  |  |  |  |  |  |  |  |  |  | 1 |
| Lee, 2019 |  |  |  |  |  |  |  |  |  |  |  |  |  |  |  |  |  |  |  |  |  |  |  |  |  |  |  |  |  |  |  |  | X |  |  |  | 1 |
| Letourneau, 2007 |  |  |  |  |  |  |  |  |  |  |  | x |  |  |  |  |  |  |  |  |  |  |  |  |  |  |  |  |  |  |  |  |  |  |  |  | 1 |
| Letourneau, 2011 |  |  |  | x | x | x |  |  |  |  |  | X |  |  |  |  |  |  |  |  |  |  |  |  |  |  |  |  |  |  |  |  |  |  |  |  | 4 |
| Levenson 2003 |  |  |  |  |  |  |  |  |  |  |  |  |  |  |  |  |  |  |  |  |  |  |  |  |  |  |  |  |  |  |  | x |  |  |  |  | 1 |
| Levenson, 2007 |  |  |  |  |  |  |  |  |  |  |  |  |  |  |  |  |  |  |  |  |  |  |  |  |  |  |  |  |  |  |  | x |  |  |  |  | 1 |
| Levenson, 2010 |  |  |  |  |  |  |  |  |  |  |  |  |  |  |  |  |  |  |  |  |  |  |  |  |  |  |  |  |  |  |  | x |  |  |  |  | 1 |
| Li, 2017 |  |  |  |  |  |  |  |  | x |  |  |  |  |  |  |  |  |  |  |  |  |  |  |  |  |  |  |  |  |  |  |  | x |  |  |  | 2 |
| Liu, 2015 |  |  |  |  |  |  |  |  | x |  |  |  |  |  |  |  |  |  |  |  |  |  |  |  |  |  |  |  |  |  |  |  |  |  |  |  | 1 |
| Livingston, 2013 |  | X |  |  |  |  |  |  |  |  |  |  |  |  |  |  |  |  |  |  |  |  |  |  |  |  |  |  |  |  |  |  |  |  |  |  | 1 |
| Lloyd 2017 |  |  |  |  |  |  |  |  |  |  |  |  |  |  |  |  |  |  |  |  |  |  |  |  |  |  |  |  |  |  |  |  |  |  | x |  | 1 |
| Lowe, 2006 |  |  |  |  |  |  |  |  |  |  |  |  |  |  |  |  |  |  |  |  |  | x |  |  |  |  |  |  |  |  |  |  |  |  |  |  | 1 |
| Ludman, 2007 |  |  |  |  | x |  |  |  |  |  |  | X |  |  |  |  |  |  |  |  |  |  |  |  |  |  |  |  |  |  |  |  |  |  |  |  | 2 |
| Ludman, 2016 |  |  |  |  |  |  |  |  |  |  |  |  |  |  |  |  |  |  |  | x |  |  |  |  |  |  |  |  |  |  |  |  |  |  |  |  | 1 |
| Macias, 2015 |  |  |  |  |  |  |  |  |  |  |  |  |  |  |  |  |  |  | x |  |  |  |  |  |  |  |  |  |  |  |  |  |  |  |  |  | 1 |
| Mahlke, 2017 |  | X | x |  |  |  |  |  |  |  |  | x |  | X | X |  |  |  |  |  | x |  |  |  |  |  |  |  |  |  | x |  |  |  |  |  | 7 |
| Mak, 2017 |  |  |  |  |  |  |  |  |  |  |  |  |  |  |  |  |  |  |  |  |  |  |  |  |  |  |  |  |  |  | x |  |  |  |  |  | 1 |
| Mak, 2021 |  |  |  |  |  |  |  |  |  |  |  |  |  |  |  |  |  |  |  |  |  |  |  |  |  |  |  |  |  |  | x |  |  |  |  |  | 1 |
| Mancini, 2005 |  |  |  |  |  |  |  |  |  |  |  |  |  |  |  |  |  |  |  |  |  |  |  |  |  |  |  |  |  | x |  |  |  |  |  |  | 1 |
| Mancini, 2018 |  |  |  |  |  |  |  |  |  |  |  |  |  |  |  |  |  |  |  |  | X |  |  | x |  |  |  |  |  |  |  |  |  |  |  |  | 2 |
| Mancini & Lawson, 2009 |  |  |  |  |  |  |  |  |  |  |  |  |  |  |  |  |  |  |  |  | x |  |  |  |  |  |  |  |  | x |  |  |  |  |  | x | 3 |
| Mangrum, 2008 |  |  |  |  |  |  |  |  |  |  |  |  |  |  |  | x |  |  |  |  |  |  |  |  |  | x |  |  |  |  |  |  |  |  |  |  | 2 |
| Manning & Suire, 1996 | x |  |  |  |  |  |  |  |  |  |  |  |  |  |  |  |  |  |  |  |  |  |  |  |  |  |  |  |  | x |  |  |  |  |  |  | 2 |
| Martin-Calero, 2017 |  |  |  |  |  |  |  |  |  |  |  |  |  |  |  |  |  | x |  |  |  |  |  |  |  |  |  |  |  |  |  |  |  |  |  |  | 1 |
| Maselko, 2020 |  |  |  | x |  |  |  |  |  |  |  |  |  |  |  |  |  |  |  |  |  |  |  |  |  |  | x |  |  |  |  |  |  |  |  |  | 2 |
| Mathias, 2019 |  |  |  |  |  |  |  |  |  |  |  |  |  |  |  |  |  |  |  |  |  |  |  |  |  |  | x |  |  |  |  |  |  |  |  |  | 1 |
| Mathews, 2018 |  |  |  |  |  |  |  |  |  |  |  | X |  |  |  |  |  |  |  |  |  |  |  |  |  |  |  |  |  |  |  |  |  |  |  |  | 1 |
| Mauthner, 1995 |  |  |  |  |  |  |  |  |  |  |  |  |  |  |  |  |  |  |  |  |  |  |  |  |  |  |  |  | x |  |  |  |  |  |  |  | 1 |
| Mazzoni, 2018 |  |  |  | x |  |  |  |  |  |  |  |  |  |  |  |  |  |  |  |  |  |  |  |  |  |  |  |  |  |  |  |  |  |  |  |  | 1 |
| McCarthy 2019 |  |  |  |  |  |  |  |  |  |  |  |  |  |  |  |  |  |  |  |  |  |  |  |  |  |  |  |  |  |  | x |  |  |  |  | x | 2 |
| Meehan, 2002 |  |  |  |  |  |  |  |  |  |  |  |  |  |  |  |  |  |  |  |  |  |  |  |  |  |  |  |  |  | x |  |  |  |  |  |  | 1 |
| Merritt, 2020 |  |  |  |  |  |  |  |  |  |  |  |  |  |  |  |  |  |  |  |  |  |  |  |  |  |  |  |  |  |  | x |  |  |  |  |  | 1 |
| Migdole, 2011 |  |  |  |  |  |  |  |  |  |  |  |  |  |  |  |  |  |  |  |  |  |  |  | x |  |  |  |  |  |  |  |  |  |  | x |  | 2 |
| Miller, 1998 |  |  |  |  |  |  |  |  |  |  |  |  |  |  |  |  |  |  |  |  |  |  |  |  |  |  |  |  |  |  |  | x |  |  |  |  | 1 |
| Min et al 2007 |  |  |  |  |  |  |  |  |  |  |  |  |  |  |  | x |  |  |  |  |  |  |  |  |  | x |  |  |  |  |  |  |  |  |  |  | 2 |
| Mishara, 2012 |  |  |  |  |  |  |  |  |  |  |  |  |  |  |  |  |  |  |  |  |  |  |  |  |  |  |  |  |  |  |  | x |  |  |  |  | 1 |
| Moll, 2009 | x |  |  |  |  |  |  |  |  |  |  |  |  |  |  |  |  |  |  |  | x |  |  |  |  |  |  | x |  | x |  |  |  |  |  | x | 5 |
| Montgomery, 2012 |  |  |  |  |  |  |  |  |  |  |  |  |  |  |  |  |  |  |  |  |  |  |  |  |  |  |  |  | x |  |  |  |  |  |  |  | 1 |
| Moorthi, 2014 |  |  |  |  |  |  |  |  |  |  |  |  |  |  |  |  |  |  |  |  |  |  |  |  |  |  |  |  |  |  |  |  | x |  |  |  | 1 |
| Moran, Russinova, 2012 |  |  |  |  |  |  |  |  |  |  |  |  |  |  |  |  |  | X |  |  |  |  |  |  |  |  |  |  |  |  |  |  |  |  |  |  | 1 |
| Moran et al 2012 | x |  |  |  |  |  |  |  |  |  |  |  |  |  |  |  |  | X |  |  |  |  |  |  |  |  |  |  |  |  |  |  |  |  |  |  | 2 |
| Moran et al 2013 | x |  |  |  |  |  |  |  |  |  |  |  |  |  |  |  |  | x |  |  | x |  |  |  |  |  |  | X |  |  |  |  |  |  |  |  | 4 |
| Moran 2014 |  |  |  |  |  |  |  |  |  |  |  |  |  |  |  |  |  |  |  |  | x |  |  |  |  |  |  |  |  |  |  |  |  |  |  |  | 1 |
| Moran et al. 2017 |  |  |  |  |  |  |  |  |  |  |  |  |  |  |  |  |  |  |  |  | x |  |  |  |  |  |  |  |  |  |  |  |  |  |  |  | 1 |
| Mourra 2014 |  |  |  |  |  |  |  |  |  |  |  |  |  |  |  |  |  |  |  |  | x |  |  |  |  |  |  |  |  |  |  |  |  |  |  |  | 1 |
| Moran, 2018 |  |  |  |  |  |  |  |  |  |  |  |  |  |  |  |  |  |  |  |  |  |  |  |  |  |  |  |  |  |  |  |  | x |  |  |  | 1 |
| Mowbray, 1996 | x |  |  |  |  |  |  |  |  |  |  |  |  |  |  |  |  |  |  |  |  |  |  |  |  |  |  |  |  | x |  |  |  |  |  |  | 2 |
| Mowbray 1998 | x |  |  |  |  |  |  |  |  |  |  |  |  |  |  |  |  | x |  |  |  |  |  |  |  |  |  | x |  | x |  |  |  |  |  |  | 4 |
| Mueller 2018 |  |  |  |  |  |  |  |  |  |  |  |  |  |  |  |  |  |  | x |  |  |  |  |  |  |  |  |  |  |  |  |  |  |  |  |  | 1 |
| Mulfinger 2018 |  |  |  |  |  |  |  |  |  |  |  |  | x |  |  |  |  |  |  |  |  |  |  |  |  |  |  |  |  |  |  |  |  |  |  |  | 1 |
| Mulvale, 2019 |  |  |  |  |  |  |  |  |  |  |  |  |  |  |  |  |  |  |  |  |  |  |  |  |  |  |  |  |  |  |  |  |  |  | x |  | 1 |
| Muralidharan, 2018 |  |  |  |  |  |  |  |  |  |  |  |  |  |  |  |  |  |  | x |  |  |  |  |  |  |  |  |  |  |  |  |  |  |  |  |  | 1 |
| Muralidharan, 2020 |  |  |  |  |  |  |  |  |  |  |  |  |  |  |  |  |  |  |  |  |  |  |  |  |  |  |  |  |  |  | x |  |  |  |  |  | 1 |
| Myers, 2021 |  |  |  |  |  |  |  |  |  |  |  |  |  |  |  |  |  |  |  |  |  |  |  |  |  |  |  |  |  |  | x |  |  |  |  |  | 1 |
| Nannen 2015 |  |  |  |  |  |  |  |  |  |  |  |  |  |  |  |  |  | x |  |  |  |  |  |  |  |  |  |  |  |  |  |  |  |  |  |  | 1 |
| Naslund 2018 |  |  |  |  |  |  |  |  |  |  |  |  |  |  |  |  |  |  | x |  |  |  |  |  |  |  |  |  |  |  |  |  |  |  |  |  | 1 |
| Nelson, 2007 |  |  |  |  |  |  |  |  |  |  |  |  |  |  |  |  |  |  |  |  |  |  | x |  |  |  |  |  |  |  |  |  |  |  |  |  | 1 |
| Nelson, 2019 |  |  |  |  |  |  |  |  |  |  |  |  |  |  |  |  |  |  |  | x |  |  |  |  |  |  |  |  |  |  |  |  |  |  |  |  | 1 |
| Neuner, 2008 |  |  |  |  |  |  |  |  |  |  |  |  |  |  |  |  |  |  |  |  |  |  |  |  |  |  | x |  |  |  |  |  |  |  |  |  | 1 |
| Nicolaidis, Mejia, 2013 |  | X |  |  |  |  |  |  |  |  |  |  |  |  |  |  |  |  |  |  |  |  |  |  |  |  |  |  |  |  |  |  |  |  |  |  | 1 |
| Nicolaidis, Wahab, 2013 |  | X |  |  |  |  |  |  |  |  |  |  |  |  |  |  |  |  |  |  |  |  |  |  |  |  |  |  |  |  |  |  |  |  |  |  | 1 |
| Niederkrotenthaler, 2016 |  |  |  |  |  |  |  |  |  |  |  |  |  |  |  |  |  |  |  |  |  |  |  |  |  |  |  |  |  |  |  | x |  | x |  |  | 2 |
| Noordenbos 2009 |  |  |  |  |  |  |  |  |  |  |  |  |  |  |  |  |  |  |  |  |  | x |  |  |  |  |  |  |  |  |  |  |  |  |  |  | 1 |
| Nossek, 2021 |  |  |  |  |  |  |  |  |  |  |  |  |  |  |  |  |  |  |  |  |  |  |  |  |  |  |  |  |  |  |  |  |  |  | x |  | 1 |
| Nyamathi, 2016 |  |  |  |  |  |  |  |  |  |  |  |  |  |  |  |  |  |  |  | x |  |  |  |  |  |  |  |  |  |  |  |  |  |  |  |  | 1 |
| O’Born, 2019 |  |  |  |  |  |  |  |  |  |  |  |  |  |  |  |  |  |  |  |  |  |  |  |  |  |  |  |  |  |  |  |  |  |  | x |  | 1 |
| O’Connell, 2014 |  |  |  |  |  |  |  |  |  |  |  |  |  |  |  | x |  |  |  |  |  |  |  |  |  |  |  |  |  |  |  |  |  |  |  |  | 1 |
| O’Connell, 2018 |  |  |  |  |  |  |  |  |  |  |  | x |  | x |  |  |  |  |  | x |  |  |  |  |  |  |  |  |  |  | x |  |  |  |  |  | 4 |
| O’Connell, 2020 |  |  |  |  |  |  |  |  |  |  |  |  |  |  |  |  |  |  |  | x |  |  |  |  |  |  |  |  |  |  |  |  |  |  |  |  | 1 |
| O’Donnell, 1999 |  |  |  |  |  |  |  |  |  | X | x |  |  |  |  |  | X |  |  |  |  |  |  |  |  |  |  |  |  |  |  |  |  |  |  |  | 3 |
| O’Leary, 2017 |  |  |  |  |  |  |  |  |  |  |  |  |  |  |  |  |  |  | x |  |  |  |  |  |  |  |  |  |  |  |  |  |  |  |  |  | 1 |
| O’Shea, 2019 |  |  |  |  |  |  |  |  |  |  |  |  |  |  |  |  |  |  | x |  |  |  |  |  |  |  |  |  |  |  |  |  |  |  |  |  | 1 |
| Ockwell, 2012 |  |  |  |  |  |  |  |  |  |  |  |  |  |  |  |  |  |  |  |  |  |  |  |  |  |  |  |  |  |  |  |  |  |  |  | x | 1 |
| Ogundipe, 2019 |  |  |  |  |  |  |  |  |  |  |  |  |  |  |  |  |  |  |  |  |  |  |  |  |  |  |  |  |  |  | x |  |  |  |  |  | 1 |
| Otte, 2019 |  |  |  |  |  |  |  |  |  |  |  |  |  |  |  |  |  |  |  |  |  |  |  |  |  |  |  |  |  |  | x |  |  |  |  |  | 1 |
| Otte, 2020a |  |  |  |  |  |  |  |  |  |  |  |  |  |  |  |  |  |  |  |  |  |  |  |  |  |  |  |  |  |  |  |  |  |  | X |  | 1 |
| Otte, 2020b |  |  |  |  |  |  |  |  |  |  |  |  |  |  |  |  |  |  |  |  |  |  |  |  |  |  |  |  |  |  |  |  |  |  | x |  | 1 |
| Paulson, 1999 |  |  |  |  |  |  |  |  |  |  |  |  |  |  |  |  |  |  |  |  |  |  |  |  |  |  |  |  |  | x |  |  |  |  |  |  | 1 |
| Park, 2020 |  |  |  |  |  |  |  |  |  |  |  |  |  |  |  |  |  |  |  |  |  |  |  |  |  |  |  |  |  |  | x |  |  |  |  |  | 1 |
| Pathare, 2018 |  |  |  |  |  |  |  |  |  |  |  |  |  |  |  |  |  |  |  |  |  |  |  |  |  |  |  |  |  |  |  |  | x |  |  |  | 1 |
| Perez, 2014 |  |  |  |  |  |  |  |  |  |  |  |  |  |  |  |  |  |  |  |  |  |  |  |  | X |  |  |  |  |  |  |  |  |  |  |  | 1 |
| Perlick 2011 |  |  |  |  |  |  |  |  |  |  |  |  | x |  |  |  |  |  |  |  |  |  |  |  |  |  |  |  |  |  |  |  |  |  |  |  | 1 |
| Perrin & Blagden, 2014 |  |  |  |  |  |  |  |  |  |  |  |  |  |  |  |  |  |  |  |  |  |  |  |  |  |  |  |  |  |  |  | x |  |  |  |  | 1 |
| Pfeiffer, 2017 |  |  |  |  |  |  |  |  |  |  |  |  |  |  |  |  |  |  |  | x | x |  |  |  |  |  |  |  |  |  |  |  |  |  |  |  | 2 |
| Pfeiffer, 2018 |  |  |  |  |  |  |  |  |  |  |  |  |  |  |  |  |  |  |  |  |  |  |  |  |  |  |  |  |  |  |  | x |  |  |  |  | 1 |
| Pfeiffer, 2019 |  |  |  |  |  |  |  |  |  |  |  | x |  |  |  |  |  |  |  | x |  |  |  |  |  |  |  |  |  |  |  |  |  | x |  |  | 3 |
| Pfeiffer, 2020 |  |  |  |  |  |  |  |  |  |  |  |  |  |  |  |  |  |  |  | x |  |  |  |  |  |  |  |  |  |  |  |  |  |  |  |  | 1 |
| Pickett, 2012 |  | X |  |  |  |  |  |  | X |  |  |  |  |  |  |  | x |  |  |  |  |  |  |  |  |  |  |  |  |  |  |  |  |  |  |  | 3 |
| Pollitt 2012 |  |  |  |  |  |  |  |  |  |  |  |  |  |  |  |  |  |  |  |  | x |  |  |  |  |  |  |  |  |  |  |  |  |  |  |  | 1 |
| Possemato, 2019 |  |  |  |  |  |  |  |  |  |  |  |  |  |  |  |  |  |  |  |  |  |  |  |  |  |  |  |  |  |  | x |  |  |  |  |  | 1 |
| Proudfoot 2012 |  |  |  |  |  |  | x |  | X |  |  |  |  |  | X |  |  |  | x |  |  |  |  |  |  |  |  |  |  |  |  |  |  |  |  |  | 4 |
| Pudlinski, 1998 |  |  |  |  |  |  |  |  |  |  |  |  |  |  |  |  |  |  |  |  |  |  |  |  |  |  |  |  |  | x |  |  |  |  |  |  | 1 |
| Pudlinski, 2001 |  |  |  |  |  |  |  |  |  |  |  |  |  |  |  |  |  |  |  |  |  |  |  |  |  |  |  |  |  | x |  |  |  |  |  |  | 1 |
| Qian 2015 |  |  | x |  |  |  |  |  |  |  |  |  |  | X |  |  |  |  |  |  |  |  |  |  |  |  |  |  |  |  |  |  |  |  |  |  | 2 |
| Rakis, Monroe, 1989 |  |  |  |  |  |  |  |  |  |  |  |  |  |  |  |  |  |  |  |  |  |  |  |  |  |  |  |  |  |  |  | x |  |  |  |  | 1 |
| Ramchand, 2019 |  |  |  |  |  |  |  |  |  |  |  |  |  |  |  |  |  |  |  |  |  |  |  |  |  |  |  |  |  |  |  | x |  |  |  |  | 1 |
| Ramjan, 2017 |  |  |  |  |  |  |  |  |  |  |  |  |  |  |  |  |  |  |  |  |  |  |  |  | X |  |  |  |  |  |  |  |  |  |  |  | 1 |
| Ramjan, 2018 |  |  |  |  |  |  |  |  |  |  |  |  |  |  |  |  |  |  |  |  |  | x |  |  | X |  |  |  |  |  |  |  |  |  |  |  | 2 |
| Ranzenhofer, 2020 |  |  |  |  |  |  |  |  |  |  |  | X |  |  |  |  |  |  |  |  |  | x |  |  | x |  |  |  |  |  | x |  |  |  |  |  | 4 |
| Raymond, 2009 |  |  |  |  |  |  |  |  |  |  |  |  |  |  |  |  |  |  |  |  |  |  |  |  |  |  |  |  | x |  |  |  |  |  |  |  | 1 |
| Rebeiro Gruhl, 2015 |  |  |  |  |  |  |  |  |  |  |  |  |  |  |  |  |  |  |  |  | x |  |  |  |  |  |  | x |  |  |  |  |  |  |  |  | 2 |
| Repper and Carter 2011 |  |  |  |  |  |  |  |  |  |  |  |  |  |  |  |  |  | x |  |  |  |  |  |  |  |  |  |  |  |  |  |  |  |  |  |  | 1 |
| Repper and Watson, 2012 |  |  |  |  |  |  |  |  |  |  |  |  |  |  |  |  |  |  |  |  |  |  |  |  |  |  |  |  |  |  |  |  |  |  |  | x | 1 |
| Resnick & Rosenheck, 2008 |  | X |  |  |  |  |  |  |  |  |  |  |  |  |  |  |  |  |  |  |  |  |  |  |  |  |  |  |  |  |  |  |  |  |  |  | 1 |
| Reynolds 2004 |  |  | x |  |  |  |  |  |  |  |  |  |  |  |  |  |  |  |  |  |  |  |  |  |  |  |  |  |  |  |  |  |  |  |  |  | 1 |
| Richard, 2009 |  |  |  |  |  |  |  |  |  |  |  |  |  |  |  |  |  |  |  |  |  |  |  |  |  |  |  |  |  | x |  |  |  |  |  |  | 1 |
| Rivera, 2007 |  |  |  |  | x |  | x |  |  | X | x | x |  | X | X |  | x |  |  |  |  |  | x |  |  |  |  |  |  |  |  |  |  |  |  |  | 9 |
| Rodham, 2007 |  |  |  |  |  |  |  |  |  |  |  |  |  |  |  |  |  |  |  |  |  |  |  |  |  |  |  |  |  |  |  | x |  |  |  |  | 1 |
| Rogers et al. 2007 |  | X |  |  |  |  | x |  |  | X | x |  |  | X |  |  |  |  |  |  |  |  |  |  |  |  |  |  |  |  |  |  |  |  |  |  | 5 |
| Rogers 2016 |  |  |  |  |  |  |  |  |  |  |  | X |  |  | X |  |  |  |  | x |  |  |  |  |  |  |  |  |  |  | x |  |  |  |  |  | 4 |
| Rohrbach, 2022 |  |  |  |  |  |  |  |  |  |  |  |  |  |  |  |  |  |  |  |  |  |  |  |  | x |  |  |  |  |  |  |  |  |  |  |  | 1 |
| Rotherham-Borus, 2014 |  |  |  |  |  | X |  |  |  |  |  |  |  |  |  |  |  |  |  |  |  |  |  |  |  |  |  |  |  |  |  |  |  |  |  |  | 1 |
| Rotondi 2005 |  |  |  |  |  |  |  |  |  |  |  |  |  |  |  |  |  |  | x |  |  |  |  |  |  |  |  |  |  |  |  |  |  |  |  |  | 1 |
| Rotondi 2010 |  |  |  |  |  |  |  |  |  |  |  |  |  |  |  |  |  |  | x |  |  |  |  |  |  |  |  |  |  |  |  |  |  |  |  |  | 1 |
| Rowe 2007 |  |  | x |  |  |  |  |  |  |  |  |  |  | X |  | x |  |  |  |  |  |  |  |  |  |  |  |  |  |  |  |  |  |  |  |  | 3 |
| Rozanov, 2002 |  |  |  |  |  |  |  |  |  |  |  |  |  |  |  |  |  |  |  |  |  |  |  |  |  |  |  |  |  |  |  | x |  |  |  |  | 1 |
| Rusch 2014 |  | X |  |  |  |  |  | X |  |  |  | X | x |  |  |  |  |  |  |  |  |  |  |  |  |  |  |  |  |  |  |  |  |  |  |  | 4 |
| Rusch 2019 |  |  |  |  |  |  |  |  |  |  |  |  | x |  |  |  |  |  |  |  |  |  |  |  |  |  |  |  |  |  |  |  |  |  |  |  | 1 |
| Russinova et al 2014 |  | X |  |  |  |  |  | x |  |  |  | X | x |  |  |  |  |  |  |  |  |  |  |  |  |  |  |  |  |  |  |  |  |  |  |  | 4 |
| Salvatore, 2010 |  |  |  |  |  |  |  |  |  |  |  |  |  |  |  |  |  |  |  |  |  |  |  |  |  |  |  |  |  |  |  |  |  | x |  |  | 1 |
| Salyers et al 2009 |  |  |  |  |  |  |  |  | X |  |  |  |  |  |  |  |  |  |  |  |  |  |  |  |  |  |  |  |  | x |  |  |  |  |  |  | 2 |
| Salyers et al 2010 |  |  |  |  |  |  |  |  | X |  |  |  |  |  |  |  |  |  |  |  |  |  |  |  |  |  |  |  |  |  |  |  |  |  |  |  | 1 |
| Salyers 2017 |  |  |  |  |  |  |  |  |  |  |  |  |  |  |  |  |  |  | x |  |  |  |  |  |  |  |  |  |  |  |  |  |  |  |  |  | 1 |
| Salzer et al 2016 |  | X |  |  |  |  |  |  |  |  |  | X |  |  | x |  |  |  |  | x |  |  |  |  |  |  |  |  |  |  |  |  |  |  |  |  | 4 |
| Salzer and shear 2002 | x |  |  |  |  |  |  |  |  |  |  |  |  |  |  |  |  |  |  |  |  |  |  |  |  |  |  |  |  |  |  |  |  |  |  |  | 1 |
| Samudre, 2016 |  |  |  |  |  |  |  |  |  |  |  |  |  |  |  |  |  |  |  |  |  |  |  |  |  |  |  |  |  |  |  |  | x |  |  |  | 1 |
| Samuels, 2018a |  |  |  |  |  |  |  |  |  |  |  |  |  |  |  |  |  |  |  | x |  |  |  |  |  |  |  |  |  |  |  |  |  |  |  |  | 1 |
| Samuels, 2018b |  |  |  |  |  |  |  |  |  |  |  |  |  |  |  |  |  |  |  | x |  |  |  |  |  |  |  |  |  |  |  |  |  |  |  |  | 1 |
| Sanders, 1998 |  |  |  |  |  |  |  |  |  |  |  |  |  |  |  |  |  |  |  |  |  |  |  |  |  | x |  |  |  |  |  |  |  |  |  |  | 1 |
| Sandoval 2019 |  |  |  |  |  |  |  |  |  |  |  |  |  |  |  |  |  |  | x |  |  |  |  |  |  |  |  |  |  |  |  |  |  |  |  |  | 1 |
| Scanlan, 2017 |  |  |  |  |  |  |  |  |  |  |  |  |  |  |  |  |  |  |  |  |  |  |  |  |  |  |  |  |  |  | x |  |  |  |  |  | 1 |
| Schlosser, 2018 |  |  |  |  |  |  |  |  |  |  |  |  |  |  |  |  |  |  | x |  |  |  |  |  |  |  |  |  |  |  |  |  |  |  |  |  | 1 |
| Scott, 2011 |  |  |  |  |  |  |  |  |  |  |  |  |  |  |  |  |  |  |  |  |  |  |  |  |  |  |  | x |  |  |  |  |  |  |  |  | 1 |
| Seeley, 2016 |  |  |  |  |  |  |  |  |  |  |  |  |  |  | x |  |  |  |  |  |  |  |  |  |  |  |  |  |  |  |  |  |  |  |  |  | 1 |
| Segal, Silverman, 2002 |  |  |  |  |  |  |  |  |  |  |  |  |  |  |  |  |  |  |  |  |  |  | x |  |  |  |  |  |  |  |  |  |  |  |  |  | 1 |
| Segal, Silverman, Temkin 2010 |  | X |  |  |  |  |  |  |  |  |  |  |  |  |  |  |  |  |  |  |  |  |  |  |  |  |  |  |  |  |  |  |  |  |  |  | 1 |
| Segal, Silverman, Temkin 2011 |  | X |  |  |  |  | x |  |  |  |  |  |  |  |  |  |  |  |  |  |  |  |  |  |  |  |  |  |  |  |  |  |  |  |  |  | 2 |
| Sells 2006 |  |  |  |  |  |  | x |  |  | X | x |  |  |  | X |  | x |  |  |  |  |  | X |  |  |  |  |  |  |  |  |  |  |  |  |  | 6 |
| Sells 2008 |  |  | x |  | x |  |  |  |  |  |  |  |  | X |  |  |  |  |  |  |  |  | X |  |  |  |  |  |  |  |  |  |  |  |  |  | 4 |
| Sells, 2020 |  |  |  |  |  |  |  |  |  |  |  |  |  |  |  |  |  |  |  |  |  |  |  |  |  |  |  |  |  |  | x |  |  |  |  |  | 1 |
| Shaw, 2021 |  |  |  |  |  |  |  |  |  |  |  |  |  |  |  |  |  |  |  |  |  |  |  |  |  |  |  |  |  |  | x |  |  |  |  |  | 1 |
| Sheehan, 2018 |  |  |  |  |  |  |  |  |  |  |  |  |  |  |  |  |  |  |  |  |  |  |  |  |  |  |  |  |  |  |  |  |  |  |  | x | 1 |
| Shepardson, 2019 |  |  |  |  |  |  |  |  |  |  |  |  |  |  |  |  |  |  |  |  |  |  |  | x |  |  |  |  |  |  |  |  |  |  |  |  | 1 |
| Sheth, 2019 |  |  |  |  |  |  |  |  |  |  |  |  |  |  |  |  |  |  |  |  |  |  |  |  |  |  |  |  |  |  |  |  | x |  |  |  | 1 |
| Shorey, 2019 |  |  |  | x |  |  |  |  |  |  |  | X |  |  |  |  |  |  |  |  |  |  |  |  |  |  |  |  |  |  |  |  |  |  |  |  | 2 |
| Siantz, 2016 |  |  |  |  |  |  |  |  |  |  |  |  |  |  |  |  |  |  |  |  |  |  |  | x |  |  |  |  |  |  |  |  |  |  |  |  | 1 |
| Siantz, 2017 |  |  |  |  |  |  |  |  |  |  |  |  |  |  |  |  |  |  |  |  |  |  |  |  |  |  |  |  |  |  |  |  |  |  |  | x | 1 |
| Siatnz, 2018 |  |  |  |  |  |  |  |  |  |  |  |  |  |  |  |  |  |  |  |  | x |  |  |  |  |  |  |  |  |  |  |  |  |  |  |  | 1 |
| Sikander, 2019 |  |  |  | x |  | X |  |  |  |  |  |  |  |  |  |  |  |  |  |  |  |  |  |  |  |  | x |  |  |  |  |  |  |  |  |  | 3 |
| Silver, 2004 |  |  |  |  |  |  |  |  |  |  |  |  |  |  |  |  |  |  |  |  |  |  |  |  |  |  |  |  |  | x |  |  |  |  |  |  | 1 |
| Simmons, 2018 |  |  |  |  |  |  |  |  |  |  |  |  |  |  |  |  |  |  |  |  |  |  |  | x |  |  |  |  |  |  |  |  |  |  |  |  | 1 |
| Simon 2011 |  |  |  |  |  |  | x |  |  |  |  |  |  |  | X |  |  |  | x |  |  |  |  |  |  |  |  |  |  |  |  |  |  |  |  |  | 3 |
| Simpson, 2013 |  |  |  |  |  |  |  |  |  |  |  |  |  |  |  |  |  |  |  |  |  |  |  |  |  |  |  |  |  |  |  |  |  |  | x |  | 1 |
| Simpson, 2014 |  |  |  |  |  |  |  |  |  |  |  |  |  |  | x |  |  |  |  |  | x |  |  |  |  |  |  | x |  |  | x |  |  |  |  |  | 4 |
| Simpson 2018 |  |  |  |  |  |  |  |  |  |  |  |  |  |  |  |  |  | x |  |  |  |  |  |  |  |  |  |  |  |  |  |  |  |  |  |  | 1 |
| Sinclair 2018 |  |  |  |  |  |  |  |  |  |  |  |  |  |  |  |  |  | x |  |  |  |  |  |  |  |  |  |  |  |  |  |  |  |  |  |  | 1 |
| Singer 2011 |  |  |  |  |  |  |  |  |  |  |  |  |  |  |  |  |  | x |  |  |  |  |  |  |  |  |  |  |  |  |  |  |  |  |  |  | 1 |
| Singla, 2020 |  |  |  |  |  |  |  |  |  |  |  |  |  |  |  |  |  |  |  |  |  |  |  |  |  |  | x |  |  |  |  |  |  |  |  |  | 1 |
| Sledge 2011 |  |  |  |  |  |  | x |  |  | x | x |  |  | X | X |  | x |  |  |  |  |  |  |  |  |  |  |  |  |  |  |  |  |  |  |  | 6 |
| Smelson et al. 2013 |  |  |  |  |  |  |  |  |  |  |  |  |  |  |  | x |  |  |  |  |  |  |  |  |  |  |  |  |  |  |  |  |  |  |  |  | 1 |
| Smith, 2017 |  |  |  |  |  |  |  |  |  |  |  |  |  |  |  |  |  |  |  |  | x |  |  |  |  |  |  |  |  |  |  |  |  |  |  |  | 1 |
| Smith-Merry, 2015 |  |  |  |  |  |  |  |  |  |  |  |  |  |  |  |  |  |  |  |  |  |  |  |  |  |  |  |  |  |  |  |  |  |  |  | x | 1 |
| Solomon 1995 |  |  |  |  | x |  | x |  |  | x | x | X |  |  | X |  | x |  |  |  |  |  | X |  |  |  |  |  |  |  |  |  |  |  |  |  | 8 |
| Soloman & Draine 1995 |  |  |  |  |  |  |  |  |  |  |  |  |  |  |  |  | x |  |  |  | x |  | x |  |  |  |  |  |  |  |  |  |  |  |  |  | 3 |
| Stanford, 2014 |  |  |  |  |  |  |  |  |  |  |  |  |  |  |  |  |  |  |  |  |  |  |  |  |  |  | x |  |  |  |  |  |  |  |  |  | 1 |
| Starnino 2010 |  |  |  |  |  |  |  |  | X |  |  |  |  |  |  |  |  |  |  |  |  |  |  |  |  |  |  |  |  |  |  |  |  |  |  |  | 1 |
| Steigman, 2014 |  |  |  |  |  |  |  |  |  |  |  |  |  |  |  |  |  |  |  |  | x |  |  |  |  |  |  |  |  |  |  |  |  |  |  |  | 1 |
| Steinberg, 2022 |  |  |  |  |  |  |  |  |  |  |  |  |  |  |  |  |  |  |  |  |  |  |  |  | x |  |  |  |  |  |  |  |  |  |  |  | 1 |
| Stewart, 2008 |  |  |  |  |  |  |  |  |  |  |  |  |  |  |  |  |  |  |  |  | x |  |  |  |  |  |  |  |  |  |  |  |  |  |  |  | 1 |
| Storm, 2020 |  |  |  |  |  |  |  |  |  |  |  |  |  |  |  |  |  |  |  |  |  |  |  |  |  |  |  |  |  |  |  |  |  |  | x |  | 1 |
| Straughan & Bucjenham, 2006 |  |  |  |  |  |  |  |  |  |  |  |  |  |  |  |  |  |  |  |  |  |  |  |  |  |  |  |  |  | x |  |  |  |  |  |  | 1 |
| Tammentie, 2004 |  |  |  |  |  |  |  |  |  |  |  |  |  |  |  |  |  |  |  |  |  |  |  |  |  |  |  |  | x |  |  |  |  |  |  |  | 1 |
| Tang, 2018 |  |  |  |  |  |  |  |  |  |  |  |  |  |  |  |  |  |  |  |  |  |  |  |  |  |  |  |  |  |  |  |  | x |  |  |  | 1 |
| Taylor Dorer, 2018 |  |  |  |  |  |  |  |  |  |  |  |  |  |  |  |  |  |  |  |  |  |  |  |  |  |  |  |  |  |  | x |  |  |  |  |  | 1 |
| Thomas, 2016 |  |  |  |  |  |  |  |  |  |  |  |  |  |  |  |  |  |  | x |  |  |  |  |  |  |  |  |  |  |  |  |  |  |  |  |  | 1 |
| Tian, 2013 |  |  |  | x |  | X |  |  |  |  |  |  |  |  |  |  |  |  |  |  |  |  |  |  |  |  |  |  |  |  |  |  |  |  |  |  | 2 |
| Tondora, 2010 |  |  |  |  |  |  |  |  |  |  |  |  |  |  |  |  |  |  |  |  | x |  |  |  |  |  |  |  |  |  |  |  |  |  |  |  | 1 |
| Tracy et al 2011 |  |  |  |  |  |  |  |  |  |  |  |  |  |  |  | x |  |  |  |  |  |  |  |  |  | x |  |  |  |  | x |  |  |  |  |  | 3 |
| Truman, 2002 |  |  |  |  |  |  |  |  |  |  |  |  |  |  |  |  |  |  |  |  |  |  |  |  |  |  |  |  |  | x |  |  |  |  |  |  | 1 |
| Tse, 2012 |  |  |  |  |  |  |  |  |  |  |  |  |  |  |  |  |  |  |  |  |  |  |  |  |  |  |  |  |  |  |  |  | x |  |  |  | 1 |
| Tse, 2017 |  |  |  |  |  |  |  |  |  |  |  |  |  |  |  |  |  |  |  |  | x |  |  |  |  |  |  |  |  |  |  |  | X |  |  |  | 2 |
| Tse, 2019 |  |  |  |  |  |  |  |  |  |  |  |  |  |  |  |  |  |  |  |  |  |  |  |  |  |  |  |  |  |  |  |  | x |  |  |  | 1 |
| Tseris, 2020 |  |  |  |  |  |  |  |  |  |  |  |  |  |  |  |  |  |  |  |  |  |  |  |  |  |  |  |  |  |  | x |  |  |  |  |  | 1 |
| Ussery, 2006 |  |  |  |  |  |  |  |  |  |  |  |  |  |  |  |  |  |  |  |  |  |  |  |  |  |  |  |  |  |  |  | x |  |  |  |  | 1 |
| Van Erp, 2010 |  |  |  |  |  |  |  |  |  |  |  |  |  |  |  |  |  |  |  |  |  |  |  |  |  |  |  | x |  |  |  |  |  |  |  |  | 1 |
| Van Gestel-Timmermans et al 2012 |  | X | x |  | x |  | x |  | X |  |  | x |  | x |  |  |  |  |  |  |  |  |  |  |  |  |  |  |  |  |  |  |  |  |  |  | 7 |
| Van Orden, 2013 |  |  |  |  |  |  |  |  |  |  |  |  |  |  |  |  |  |  |  |  |  |  |  |  |  |  |  |  |  |  |  | x |  |  |  |  | 1 |
| Van Vugt, 2012 |  |  |  |  |  |  |  |  |  |  |  |  |  |  |  |  |  |  |  |  |  |  |  |  |  |  |  |  |  |  | x |  |  |  |  |  | 1 |
| Vayshenker et al. 2016 |  | X |  |  |  |  |  |  |  |  |  |  |  |  |  |  |  |  |  |  |  |  |  |  |  |  |  |  |  |  |  |  |  |  |  |  | 1 |
| Walker and Bryant 2013 |  |  |  |  |  |  |  |  |  |  |  |  |  |  |  |  |  | x |  |  |  |  |  |  |  |  |  |  |  |  |  |  |  |  |  |  | 1 |
| Weis, 2017 |  |  |  | x |  |  |  |  |  |  |  |  |  |  |  |  |  |  |  |  |  |  |  |  |  |  |  |  |  |  |  |  |  |  |  |  | 1 |
| Weller, 2021 |  |  |  |  |  |  |  |  |  |  |  |  |  |  |  |  |  |  |  |  |  |  |  |  |  |  |  |  |  |  |  |  |  |  | x |  | 1 |
| White, 2017 |  |  |  |  |  |  |  |  |  |  |  |  |  |  |  |  |  |  |  |  |  |  |  |  |  |  |  |  |  |  | x |  |  |  | x |  | 2 |
| White, 2020 |  |  |  |  |  |  |  |  |  |  |  |  |  |  |  |  |  |  |  |  |  |  |  |  |  |  |  |  |  |  | x |  |  |  |  |  | 1 |
| Whitlock, 2006 |  |  |  |  |  |  |  |  |  |  |  |  |  |  |  |  |  |  |  |  |  |  |  |  |  |  |  |  |  |  |  | x |  |  |  |  | 1 |
| Williams, 2018 |  |  |  |  |  |  |  |  |  |  |  |  |  |  |  |  |  |  | x |  |  |  |  |  |  |  |  |  |  |  |  |  |  |  |  |  | 1 |
| Wilson, 1999 |  |  |  |  |  |  |  |  |  |  |  |  |  |  |  |  |  |  |  |  |  |  | X |  |  |  |  |  |  |  |  |  |  |  |  |  | 1 |
| Wrobleski, 2015 |  |  |  |  |  |  |  |  |  |  |  |  |  |  | x |  |  |  |  |  |  |  |  |  |  |  |  |  |  |  |  |  |  |  |  |  | 1 |
| Wusinich, 2020 |  |  |  |  |  |  |  |  |  |  |  |  |  |  |  |  |  |  |  |  |  |  |  |  |  |  |  |  |  |  | x |  |  |  |  |  | 1 |
| Yamaguchi, 2017 |  |  |  |  |  |  |  |  |  |  |  |  |  |  | x |  |  |  | x |  |  |  |  |  |  |  |  |  |  |  |  |  | x |  |  |  | 3 |
| Yanos, 2001 |  |  |  |  |  |  |  |  |  |  |  |  |  |  |  |  |  |  |  |  |  |  | X |  |  |  |  |  |  |  |  |  |  |  |  |  | 1 |
| Young, 2017 |  |  |  |  |  |  |  |  |  |  |  |  |  |  |  |  |  |  | x |  | x |  |  |  |  |  |  |  |  |  |  |  |  |  |  |  | 2 |
| Yuen and Fossey, 2003 | x |  |  |  |  |  |  |  |  |  |  |  |  |  |  |  |  |  |  |  |  |  |  |  |  |  |  | x |  | x |  |  |  |  |  |  | 3 |
| Yuen, 2019 |  |  |  |  |  |  |  |  |  |  |  |  |  |  |  |  |  |  |  |  |  |  |  |  |  |  |  |  |  |  | x |  |  |  |  |  | 1 |
| Zeng, 2020 |  |  |  |  |  |  |  |  |  |  |  |  |  |  |  |  |  |  |  |  |  |  |  |  |  |  |  |  |  |  |  |  |  |  |  | x | 1 |
| Zenga 2018 |  |  |  |  |  |  |  |  |  |  |  |  |  |  |  |  |  | x |  |  |  |  |  |  |  |  |  |  |  |  |  |  |  |  |  |  | 1 |
| Zhang, 2016 |  |  |  | x |  | x |  |  |  |  |  |  |  |  |  |  |  |  |  |  |  |  |  |  |  |  |  |  |  |  |  |  |  |  |  |  | 2 |
| Zhang, 2018 |  |  |  | x |  |  |  |  |  |  |  |  |  |  |  |  |  |  |  |  |  |  |  |  |  |  |  |  |  |  |  |  |  |  |  |  | 1 |

* papers where we only used a portion of included studies

Counts for number of times included: 1 (n=301), 2 (n=67), 3 (n=18), 4 (n=19), 5 (n=4), 6 (n=3), 7 (n=3), 8 (n=2), 9 (n=1)

**Appendix 5:** AMSTAR2 ratings

| **AMSTAR2 CRITERIA** | **1** | **2** | **3** | **4** | **5** | **6** | **7** | **8** | **9 RCT** | **9 NRSI** | **10** | **11 RCT** | **11 NRSI** | **12** | **13** | **14** | **15** | **16** | **Overall score** |
| --- | --- | --- | --- | --- | --- | --- | --- | --- | --- | --- | --- | --- | --- | --- | --- | --- | --- | --- | --- |
| Akerblom 2022 | Yes | Yes | No | No | No | No | No | No | NA | NA | No | No MA | No MA | No MA | No | No | No MA | Yes | Critically low |
| Bailie 2014 | Yes | No | No | Partial yes | No | No | No | Partial yes | Includes only NRSI | Partial yes | No | No MA | No MA | No MA | Yes | No | No MA | No | Critically low |
| Bassuk 2016 | Yes | Partial yes | No | No | Yes | Yes | No | Partial yes | No | Partial yes | No | No MA | No MA | No MA | Yes | Yes | No MA | No | Critically low |
| Bowersox 2021 | Yes | No | Yes | No | No | No | No | No | NA | NA | No | No MA | No MA | No MA | No | No | No MA | No | Critically low |
| Burke 2019 | Yes | No | Yes | No | Yes | No | Yes | Yes | No | Partial yes | No | Yes | Yes | Yes | Yes | Yes | Yes | No | Critically low |
| Chien 2019 | Yes | Yes | No | Yes | Yes | Yes | Partial yes | Yes | Yes | Includes only RCTs | Yes | Yes | Includes only RCT | Yes | Yes | Yes | Yes | Yes | High |
| Chinman 2014 | No | No | No | No | No | No | No | Partial yes | No | No | No | No MA | No MA | No MA | Yes | Yes | No MA | Yes | Critically low |
| du Plessis 2019 | Yes | No | No | No | No | No | No | No | Includes only NRSI | No | No | No MA | No MA | No MA | No | No | No MA | Yes | Critically low |
| Fang 2021 | Yes | No | No | Partial yes | Yes | Yes | No | Yes | Yes | Includes only RCTs | No | Yes | Includes only RCTs | Yes | Yes | Yes | Yes | Yes | Critically low |
| Fortuna 2020 | Yes | Partial yes | No | Partial yes | Yes | Yes | No | Partial yes | No | No | No | No MA | No MA | No MA | No | No | No MA | Yes | Critically low |
| Fuhr 2014 | Yes | Partial yes | No | Partial yes | Yes | Yes | No | No | Yes | Includes only RCTs | No | Yes | Includes only RCTs | Yes | Yes | Yes | Yes | Yes | Low |
| Gaiser 2021 | Yes | No | No | No | Yes | Yes | No | No | No | Partial yes | No | No MA | No MA | No MA | Yes | No | No MA | Yes | Critically low |
| Huang 2020 | Yes | No | No | Partial yes | No | Yes | No | Yes | Yes | Includes only RCTs | No | Yes | Includes only RCT | Yes | No | Yes | Yes | Yes | Critically low |
| Ibrahim 2020 | Yes | Yes | No | Partial yes | Yes | Yes | No | No | Yes | Yes | No | No MA | No MA | No MA | No | **No** | No MA | Yes | Critically low |
| Jones 2013 | Yes | No | No | Partial yes | No | No | No | No | Includes only NRSI | Yes | No | No MA | No MA | No MA | No | No | No MA | Yes | Critically low |
| Lewis 2021 | No | No | No | Partial yes | No | No | No | No | No | No | No | No MA | No MA | No MA | No | No | No MA | No | Critically low |
| Lloyd-Evans 2014 | Yes | No | Yes | Yes | Yes | Yes | Yes | Yes | Yes | Includes only RCTs | No | Yes | Includes only RCT | Yes | Yes | Yes | Yes | Yes | Low |
| Lyons 2021 | Yes | Yes | Yes | Partial yes | Yes | No | Yes | Partial yes | Yes | Includes only RCTs | No | No | NA | No | Yes | Yes | No | Yes | Critically low |
| Miyamoto 2012 | No | No | No | No | No | No | No | No | No | No | No | No MA | No MA | No MA | No | No | No MA | No | Critically low |
| Mutschler 2021 | No | No | No | No | Yes | Yes | No | No | Includes only NRSI | No | No | No MA | No MA | No MA | No | No | No MA | No | Critically low |
| Ong 2022 | Yes | No | Yes | Partial yes | Yes | No | No | Partial yes | NA | NA | No | No MA | No MA | No MA | Yes | No | No MA | Yes | Critically low |
| Peck 2022 | Yes | Yes | No | Partial yes | Yes | Yes | No | Partial yes | Partial yes | Partial yes | No | Yes | No MA | No | No | Yes | Yes | Yes | Critically low |
| Pellizzer 2022 | No | No | No | No | Yes | No | No | Partial yes | Yes | Partial yes | No | No MA | No MA | No MA | Yes | Yes | No MA | Yes | Critically low |
| Pitt 2013 | Yes | Partial yes | No | Partial yes | Yes | Yes | Yes | Yes | Yes | Includes only RCTs | Yes | Yes | Includes only RCT | No | Yes | Yes | No | Yes | Low |
| Reif 2014 | Yes | No | No | No | No | No | No | Partial yes | No | No | No | No MA | No MA | No MA | Yes | Yes | No MA | Yes | Critically low |
| Schlichthorst 2020 | Yes | Yes | No | Yes | No | No | No | Yes | NA | NA | No | No MA | No MA | No MA | No | No | No MA | Yes | Critically low |
| Smit 2022 | Yes | Yes | No | Partial yes | Yes | Yes | No | Partial yes | Yes | Includes only RCTs | No | Yes | NA | Yes | Yes | Yes | Yes | Yes | Low |
| Sun 2022 | No | Partial yes | No | Partial yes | Yes | No | No | Partial yes | Yes | Includes only RCTs | No | Yes | NA | No | No | No | Yes | Yes | Critically low |
| Triece 2022 | Yes | No | No | No | No | No | No | No | Yes | Yes | No | No MA | No MA | No MA | Yes | Yes | No MA | Yes | Critically low |
| Vandewalle 2016 | Yes | No | No | No | No | No | No | No | Includes only NRSI | Partial yes | No | No MA | No MA | No MA | Yes | No | No MA | Yes | Critically low |
| Viking 2022 | No | No | No | No | No | No | Yes | Partial yes | NA | NA | No | No MA | No MA | No MA | No | No | No MA | Yes | Critically low |
| Walker 2013 | Yes | No | Yes | No | No | No | No | No | Includes only NRSI | Yes | No | No MA | No MA | No MA | Yes | No | No MA | No | Critically low |
| Wang 2021 | No | No | No | Partial yes | Yes | Yes | No | No | Yes | Includes only RCTs | No | No | No MA | No | No | Yes | No | Yes | Critically low |
| White 2020 | Yes | Partial yes | No | Partial yes | Yes | No | No | Partial yes | Yes | Includes only RCTs | No | Yes | No MA | No | Yes | Yes | No | Yes | Critically low |
| Zeng 2021 | No | No | No | Partial yes | No | No | No | No | NA | NA | No | No MA | No MA | No MA | No | Yes | No MA | Yes | Critically low |

MA: Meta-Analysis

AMSTAR2 Item descriptions:

1. PICO criteria included

2. Explicit statement that the review methods were established prior to the conduct of the review

3 Selection of study designs to include explained

4. Comprehensive literature search strategy

5.Study selection performed in duplicate

6. Data extraction performed in duplicate

7. List of excluded studies and reasons provided

8. Included studies described in adequate detail

9.Satisfactory assessment of risk of bias (RoB) in individual studies

10. Sources of funding reported

11. Meta-Analysis: appropriate methods for statistical combination of results

12. Meta-Analysis: Assessment of the potential impact of RoB in individual studies

13. Interpretation accounts for RoB

14. Satisfactory explanation for, and discussion of, any heterogeneity observed in the results of the review

15. Meta-Analysis: Adequate investigation of publication bias

16. Potential conflicts of interest reported

Adaptations of AMSTAR2 criteria for scoping reviews and qualitative reviews:

1. For Yes, we removed the requirement of “comparator group” in the research question and inclusion criteria.

2. For Partial yes, we removed the requirement of inclusion of “risk of bias assessment” in the protocol. For Yes, we removed the requirement of inclusion of “plan for investigating causes of heterogeneity” in the protocol (scoping reviews only).

8. For Partial yes, we removed the requirement of “comparators” in the description of studies.

9. We marked this question as NA (scoping reviews only)

**Appendix 6:** Effectiveness of peer support outcomes: results for non-meta-analysis results

**Clinical outcomes - depression**

| **Author** | **Outcome** | **Type of review** | **Main findings** |
| --- | --- | --- | --- |
| Pitt et al | Depression | SR & MA | No effect |
| Gaiser et al | Depression symptoms | SR & NR | Reported decreases |

**Clinical outcomes – mental health symptoms**

| **Author** | **Outcome** | **Type of review** | **Main findings** |
| --- | --- | --- | --- |
| Lyons et al | Global symptoms | SR & MA | Mixed evidence. No effect on global symptoms in studies without follow up. In two trials with follow up, one reported no effect, and one reported reduced global symptoms after peer support. |
| Lyons et al | Psychiatric symptoms (anxiety and psychosis) | SR & MA | Mixed evidence. Improvements in anxiety. No improvements in psychosis. |
| Chien et al | Severity of illness – Brief Symptom Inventory – medium and long term | SR& MA | Medium term: N (studies)=1, N (participants)=458, Mean Difference -0.13, 95% CI -0.25-0.01, Z=2.08 (P=0.04). No effect  Long term: N (studies)=1, N (participants)=440, Mean Difference 0.0, 95% CI -0.11-0.11), I^2^=56.77%, Chi^2^=2.31, df=1 (P=0.13). No effect |
| Chien et al | Severity of illness – Clinical Global Impression scale (medium and long term) | SR&MA | Medium term: N (studies)=1, N (participants)=216, Mean Difference -0.30, 95% CI -0.53-0.07. No effect  Long term: N (studies)=1, N (participants)=216, Mean Difference 0.40, 95% CI 0.15-0.65. improvements |
| Chien et al | Emotional wellbeing (medium term) | SR&MA | N(studies)=1, N(participants)=57, MD=3, 95% CI= -2.76, 8.76, Z=1.02 (p=0.31); I^2^=100%, Tau^2^=0, Chi^2^=0 (df=0, p<.0001). No effect |
| Chien et al | Mental Health Confidence Scale | SR & MA | Medium term: N (studies)=1, N (participants)=221, MD 0.31, 95% CI 0.07-0.55, Z=2.54(P=0.01). Improvements (favouring PS)  Long term: N (studies)=1, N (participants)=106, MD 2.70, 95% CI -2.40-7.80, Z=1.04(P=0.3), No effect |
| Fuhr et al | SMI: Changes in Psychiatric symptoms | SR&MA | No effect of peer-delivered interventions (SMD=0.08, 95%CI=-0.11, 0.26); N(studies)=1, N(participants)=448; Z=0.82 (p=0.41). |
| Lloyd-Evans et al | Symptoms of psychosis (follow up) | SR&MA | Follow up (6 months later):  N=1, 448 participants.  SMD = -0.00 (-0.19, 0.18). No effect. |
| Lloyd-Evans et al | Overall psychiatric symptoms (post treatment and follow up) | SR&MA | Follow up (6 months):  N=1 study, 488 participants.  SMD = 0.08 (-0.26, 0.11). No effect on overall psychiatric symptoms at 6-month follow up |
| Peck et al | Symptom severity | SR&MA | Three pre-post-test studies and one RCT reported significantly reduced symptom severity. One study reported a reduction in symptom severity that did not reach significance. |
| Triece et al | Effectiveness on MH outcomes eg PTSD and depression | SR&MA | All pre-post case series found significant beneficial intervention effects. Six RCTs showed mixed evidence: 3 reported a positive effect, 2 reported both positive and null effects, and one reported no evidence for an effect. |
| Chinman et al | Mental health symptoms | SR&NR | Peers delivering curricula vs treatment as usual:  One RCT reported reductions in anxiety and depression symptoms. Two studies showed no significant differences in symptomology. |
| Fortuna et al | Psychiatric symptomatology | SR & NR | Mixed results, with some studies reporting no effect and others reporting improvements |
| Fortuna et al | Anxiety | SR & NR | No effect |
| Fortuna et al | Mental health quality | SR & NR | No effect |
| Fortuna et al | Psychiatric distress | SR & NR | No effect |
| Pellizzer and Wade | Eating disorder severity outcomes | SR & NR | Mixed evidence. Positive benefits reported in reducing eating disorder psychopathology, improved weight, and BMI. Three further RCTs did not find any differences. |
| Pellizzer and Wade | Mentor eating disorder pathology | SR & NR | Mentors did not experience pathological increases in eating disorder psychopathology. |
| Bowersox et al | Suicidal ideation/ suicide risk | Scoping review | Reduction in suicidal ideation, improvements in self-esteem and stress management, positive support and advice provided on suicide. |
| Schlichthorst et al | Suicidal thoughts | Scoping review | Improvements in understanding suicidal thoughts and decrease in suicidal thoughts. |

**Clinical outcomes – service use**

| **Author** | **Outcome** | **Type of review** | **Main findings** |
| --- | --- | --- | --- |
| Chien et al | Hospital admission | SR & MA | N (studies)=1, N (participants)=19, RR=0.44, 95% CI (0.11-1.75), Z=1.16(P=0.25), I^2^=100%, Tau^2^=0; Chi^2^=0, df=0 (P<0.0001). No clear difference. |
| Chien et al | Use of emergency care | SR & MA | N (studies)=1, N (participants)=57, RR 0.39, 95% CI 0.11-1.32, Z=1.52 (P=0.13). No clear difference |
| Chien et al | >1 primary care visit | SR & MA | N (studies)=1, N (participants)=158, Mean Difference -0.02, 95% CI -3.96-3.95, Z=0.01 (P=0.99). No effect |
| Lloyd-Evans | Hospitalisation | SR& MA | Peer-delivered services:  N=1 study, 114 participants  RR=0.68 (0.45, 1.03)  Little or no evidence that peer support was associated with positive effects on hospitalisation |
| Pitt et al | Hospital admission | SR& MA | Consumer provider vs professional:  N=1 study, 57 participants; RR=0.68, 95% CI=0.45, 1.03. No effect.  Consumer provider as adjunct to usual care:  N=74; MD=-0.64, 95% CI -1.3, 0.02)  No effect. |
| Pitt et al | Length of stay | SR& MA | Consumer provider vs professional  N=1 study; 116 participants.  MD=1.1(-0.72, 2.92).  No effect |
| Chinman et al | Rehospitalisations and hospital days | SR & NR | Peer support added to traditional services:  Mixed evidence. RCT evidence showed fewer rehospitalizations and hospital days. Two RCTs and two quasi-experimental studies found reduced inpatient service use, but this was not found in other RCTs and quasi-experimental trials. One quasi-experimental study found that the presence of a peer in ACT team was associated with an increase in psychiatric hospitalization days. |
| Fortuna et al | Healthcare utilisation | SR & NR | Improvements reported |
| Gaiser et al | Receipt of hospital care | SR & NR | Reduced frequency of use |
| Gaiser et al | Healthcare service use | SR & NR | Seven studies reported significant clinical improvement on measures of healthcare service use and management, including patient activation, medication initiation or distribution, healthcare self-management, and engagement with healthcare services. Five studies reported a decrease in patient use of emergency department services. One study reported an increase in primary care service use. |
| Gaiser et al | Inpatient service use | SR & NR | Five studies reported decreased use of inpatient care, two reported decreased length of inpatient stay, and one reported increased likelihood of inpatient service usage. |
| Gaiser et al | Use of psychotherapy services | SR & NR | Two studies reported an increased likelihood of use, whereas three reported no difference. |
| Reif et al | Readmission to hospital | SR & NR | Mixed evidence. One study (RCT) reported no difference, a second (quasi-experimental) reported One RCT reported longer stays in the community before rehospitalisation. Overall fewer participants in the peer recovery group were hospitalised. |
| Reif et al | Post-discharge participation and retention | SR & NR | Reported increases in rates of post discharge participation and retention and use of recovery coaching. |

**Clinical outcomes – other (eg help-seeking, activation)**

| **Author** | **Outcome** | **Type of review** | **Main findings** |
| --- | --- | --- | --- |
| Chien et al | General health | SR & MA | N (studies)=1, N (participants)=158, Mean Difference -0.02, 95% CI -3.96-3.95, Z=0.01 (P=0.99). No effect |
| Chien et al | Death | SR & MA | N (studies)=1, N (participants)=555, RR 1.52 95% CI 0.43, 5.31, Z=0.65(P=0.52). No effect |
| Chien et al | Approach to healthcare | SR & MA | N (studies)=1, N (participants)=57, MD=2.10, 95% CI= -0.83-5.03, Z=1.4(P=0.16), No effect |
| Chien et al | Self-management – medium term | SR & MA | N (studies)=1, N (participants)=57, MD 0.60, 95% CI -0.10-1.30, Z=1.68 (P=0.09). No effect |
| Chien et al | Internal locus of control for health (medium term) | SR & MA | N (studies)=1, N (participants)=57, MD=3.6, 95% CI=0.99, 6.21, Z=2.7 (p=0.01); I^2^=100%, Tau^2^=0, Chi^2^=0, df=0 (p<.0001).  Improvements favouring PS. |
| Huang et al | Adverse effects | SR & MA | None reported in any study |
| Pitt et al | Professional attitudes that the clients’ needs were met | SR & MA | Consumer providers as an adjunct to usual care: small, significant difference in favour of peer support (MD=1.56 (0.50, 2.62), p=.004). |
| Bassuk et al | Substance use | SR & NR | No effect. |
| Bassuk et al | Post-discharge adherence | SR & NR | Increased post-discharge adherence was reported. |
| Chinman et al | Treatment engagement | SR & NR | ACT team either with or without peers: One RCT reported better engagement six months after entering treatment.  Peer-delivered curricula vs usual care: one RCT reported greater patient activation and primary care visits. |
| Chinman et al | Attendance | SR & NR | ACT team either with or without peers: One RCT reported lower rates of nonattendance at appointments and higher rates of participation in structured social care activities. Higher treatment engagement in peer condition was also reported. |
| Fortuna et al | Psychiatric self-management | SR & NR | Improvements reported |
| Fortuna et al | Perceived control over the illness/ health locus of control | SR & NR | No effect |
| Fortuna et al | Global health quality | SR & NR | No effect |
| Fortuna et al | Health control | SR & NR | No effect |
| Fortuna et al | Mental health control | SR & NR | No effect |
| Fortuna et al | Self-perceived treatment involvement | SR & NR | No effect |
| Fortuna et al | Medication adherence | SR & NR | Mixed results, with some studies reporting no effect and others reporting improvements |
| Fortuna et al | Medication side effects | SR & NR | Improvements reported |
| Gaiser et al | Substance use | SR & NR | Reported decreases |
| Gaiser et al | Medication use | SR & NR | Increases reported |
| Pellizzer & Wafe | Engagement and retention | SR & NR | Higher engagement and retention in sessions reported. |
| Reif et al | Drug use | SR & NR | Improvements reported; lower rates of cocaine and opiate use and higher drug-free rates at six months, very little substance use at six months, reduced relapse and improvements in recognition of substance use as a problem, increased control over alcohol and drug use, decreased substance use, and fewer consequences from substance use. |
| Akerblom et al | Non specified clinical outcomes | Scoping review | Improvements reported |
| Akerblom et al | Treatment attendance | Scoping review | Improvements reported |
| Bowersox et al | Crisis support | Scoping review | Reported reductions in the length of acute suicide risk, and the average length of time that prisoners needed to be kept on emergency watch. Reductions in the length of time that prisoners met emergency, eyes-on suicide risk criteria were found in the overall prisoner population as well as vulnerable subpopulations. |

**Recovery outcomes**

| **Author** | **Outcome** | **Type of review** | **Main findings** |
| --- | --- | --- | --- |
| Lyons et al | Recovery (post-intervention) | SR & MA | No effect |
| Chien et al | Recovery | SR & MA | Long term: N (studies)=1, N (participants)=318, MD 1.58, 95% CI -0.33-3.49, Z=2.72 (P=0.01). No effect |
| Peck et al | Self-perceived recovery | SR & MA | Four pre-post-test studies and one RCT reported improved self-perceived recovery. |
| Wang et al | Wellbeing | SR & MA | N=1 study; individual-led peer support:  SMD = -2.98, 95% CI -3.34, -2.62, p<0.00001.  Improved wellbeing. |
| Bassuk | Recovery outcomes | SR & NS | Most studies reported improvements in recovery outcomes (including rehospitalisation). |
| Chinman et al | Recovery needs | SR & NS | NA |
| Chinman et al | Perceived recovery | SR & NS | Peers delivering curricula vs treatment as usual: One RCT found greater perceived recovery. |
| Fortuna et al | Personal recovery | SR & NS | Significant improvements reported |
| Gaiser et al | Behavioural health and recovery | SR & NS | Ten studies reported significant improvements in behavioral health and recovery, including emotional well-being and mental health improvements; and improved recovery and treatment outcomes. |

**Psychosocial – hope**

| **Author** | **Outcome** | **Type of review** | **Main findings** |
| --- | --- | --- | --- |
| Lyons et al | Hope (follow up) | SR & MA | One study reported a positive effect and the other reported no effect. |
| Chien et al | Hope (Herth Hope Index) | SR & MA | N (studies)=1, N (participants)=217, MD 0.24, 95% CI 0.11-0.37, Z=3.73 (P=0), I^2^=0%, Chi^2^=1.47, df=1 (P=0.69). Improvements favouring PS on this scale (but MA results on different scale show no effect). |
| Peck et al | Hopefulness | SR & MA | Three pre-post-test studies and one RCT reported significantly reduced symptom severity. One study reported a reduction in symptom severity that did not reach significance. |
| Chinman | Hopefulness | SR & NR | Peers delivering curricula vs treatment as usual: two RCTs found greater hope in condition where peers delivered curricula. |
| Fortuna et al | Hope | SR & NR | No effect |
| Akerblom | Hope and empowerment | Scoping review | Hope and self-esteem among peer workers were associated with improvements in hope and empowerment among service users over time |

**Psychosocial – empowerment**

| **Author** | **Outcome** | **Type of review** | **Main findings** |
| --- | --- | --- | --- |
| Lyons et al | Empowerment (post-intervention) | SR & MA | No effect |
| Burke et al | Empowerment | SR & MA | Mixed evidence. A positive effect of intervention on empowerment in six RCTs and two pre-post studies. No effect in four RCTs. |
| Chien et al | Empowerment (Rogers Empowerment Scale and Dutch Empowerment Scale) | SR & MA | RES: N (studies)=1, N (participants)=158, MD -0.95, 95% CI -3.30-1.40, Z=0.79 (P=0.43). No effect  DES: N (studies)=1, N (participants)=220, MD 0.19, 95% CI 0.05-0.33, Z=2.63 (P=0.01). Improvements favouring PS |
| Peck et al | Empowerment | SR & MA | One pre-post-test study and one RCT reported improved empowerment. One study reported a reduction in empowerment. |
| Pitt et al | Empowerment | SR & MA | Consumer provider as adjunct to usual care (12 month follow up)  N=1 study; 1827 participants. Time x group interaction F(1.4059)=2.30, p=0.13.  No effect. |
| Chinman et al | Empowerment | SR & NR | One RCT found increased assertiveness and empowerment. |
| Fortuna et al | Empowerment | SR & NR | No effect |
| Gaiser | Empowerment | SR & NR | Two studies reported improvements in empowerment. |
| Akerblom | Hope & empowerment | Scoping review | Hope and self-esteem among peer workers were associated with improvements in hope and empowerment among service users over time |

**Psychosocial – QoL**

| **Author** | **Outcome** | **Type of review** | **Main findings** |
| --- | --- | --- | --- |
| Lyons et al | QoL | SR & MA | Mixed findings |
| Lloyd-Evans | Quality of life (including follow up) | SR & MA | Follow up (3-6 months)  Peer-delivered services:  N=1 study, 96 participants  No effect but provided no data |
| Pitt et al | QoL | SR & MA | Consumer provider vs professional:  N=1 study, 130 participants  Mean difference=-0.30 (-0.80, 0.20)  No effect.  Consumer provider as adjunct to usual care:  N=1 study, 84 participants  No effect, and insufficient data reported to calculate a summary estimate. |
| Fortuna et al | QoL | SR & NR | No effect |
| Gaiser et al | QoL | SR & NR | Improvement in quality-of-life measures. |
| Pellizzer & Wade | QoL /wellbeing | SR & NR | Mixed evidence. Improved hope, quality of life and mood reported, although in one RCT these differences did not remain at follow up. Three RCTs did not find any effects. |
| Pellizzer & Wade | QoL and mood of mentors | SR & NR | No negative effects on eating disorder pathology or mood reported. |
| Reif | QoL and wellbeing | SR & NR | Improvements reported. |

**Psychosocial – satisfaction with care**

| **Author** | **Outcome** | **Type of review** | **Main findings** |
| --- | --- | --- | --- |
| Huang et al | Satisfaction with intervention | SR & MA | People receiving peer support: Most women were satisfied with the peer support experience.  Peer supporter satisfaction: Peer supporters were satisfied and would volunteer again.  Factors associated with service user satisfaction: The number of peer volunteer contacts, number of conversations, and number of messages left were weakly correlated with maternal overall satisfaction. Emotional support with no practical help may affect the degree of satisfaction. |
| Lloyd-Evans et al | Satisfaction | SR & MA | Peer-delivered services:  N=1 study, 87 participants  SMD=0.48 (0.05, 0.91)  Small negative effect. |
| Chinman et al | Satisfaction (life situation and finances) | SR & NR | One quasi-experimental study reported greater satisfaction with life situation and finances and fewer life problems. A second quasi-experimental study reported improved quality of life. |
| Fortuna et al | Patient satisfaction and experience | SR & NR | Mixed results, with some studies reporting no effect, others reporting improvements and one study reporting low levels of patient satisfaction with an app. |
| Reif et al | Satisfaction | SR & NR | Improvements reported |

**Psychosocial – relational**

| **Author** | **Outcome** | **Type of review** | **Main findings** |
| --- | --- | --- | --- |
| Lyons et al | Social support | SR & MA | No effect |
| Chien et al | Reliance on others | SR & MA | Long term: N (studies)=1, N (participants)=319, MD=0.41, 95% CI=-0.21-1.03, Z=1.3 (p=0.2), I^2^=100%, Tau^2^=0, Chi^2^=0 (df=0, p<.0001. No effect  Medium term: N (studies)=1, N (participants)=343, MD=0.80, 95% CI=0.17-1.43, Z=2.5 (P=0.01). Improvements favouring PS |
| Gaiser et al | Interpersonal relationships, social functioning | SR & NR | Four studies reported significant clinical improvement in interpersonal relationships, including strengthening interpersonal relationships and social functioning as well as the relationship between patient and a primary care provider |
| Bowersox et al | Sense of community | Scoping review | Increased sense of community |

**Psychosocial – self-efficacy**

| **Author** | **Outcome** | **Type of review** | **Main findings** |
| --- | --- | --- | --- |
| Chien et al | Patient self-advocacy | SR & MA | Medium term: N (studies)=1, N (participants)=458, MD 0.08, 95% CI -0.02-0.18, Z=1.63(P=0.1). No effect  Long term: N (studies)=1, N (participants)=447, MD 0.10, 95% CI 0.01-0.19, Z=2.09 (P=0.04). Improvements favouring PS |
| Chien et al | Self-management/ self-efficacy scale | SR & MA | Medium term: N (studies)=1, N (participants)=57, MD 1.20, 95% CI 0.11-2.29, Z=2.16 (P=0.03). Improvements favouring PS |
| Chien et al | General self-efficacy scale | SR & MA | Medium term: N (studies)=1, N (participants)=216, MD 0.90, 95% CI -1.04-2.84, Z=0.91 (P=0.36). No effect  Long term: N (studies)=1, N (participants)=216, MD 2.20, 95% CI 0.35-4.05, Z=2.33(P=0.02). Improvements favouring PS |
| Lyons et al | Self-efficacy | SR & MA | No effect |
| Burke et al | Self-efficacy | SR & MA | Eight RCTs reported significant improvements. No effect was found in 2 RCTs. There were contradictory findings from 2 further RCTs. |
| Akerblom et al | Patient self-efficacy | Scoping review | Higher scores on patients’ level of self-efficacy reported. |
| Schlichthorst et al | Help-seeking | Scoping review | No effect |

**Psychosocial – functioning**

| **Author** | **Outcome** | **Type of review** | **Main findings** |
| --- | --- | --- | --- |
| Chien et la | Global assessment of functioning | SR & MA | N (studies)=1, N (participants)=216, MD -3.90, 95% CI -7.81-0.01, Z=1.96 (P=0.05). No effect |
| Lloyd-Evans | Employment | SR & MA | Peer delivered services:  N= 1 study, 96 participants – no difference reported |
| Pitt et al | Functioning | SR & MA | Consumer provider compared to professional:  N=1 study, 130 participants  Mean Difference=0.0 (-0.07, 0.07) No effect  Consumer provider as adjunct to usual care:  N=1 study, 45 participants  Mean Difference=3.00 (-5.75, 11.75) No effect |
| Chinman et al | Mental and social functioning | SR & NR | One quasi-experimental study found improved social functioning in patients receiving peer support services. A second quasi-experimental study also reported improved social and mental functioning. Two other studies show no significant differences. |
| Fortuna et al | Functioning | SR & NR | No effect |

**Psychosocial - other**

| **Author** | **Outcome** | **Type of review** | **Main findings** |
| --- | --- | --- | --- |
| Burke et al | Internalised stigma | SR & MA | Mixed evidence. Three studies (1 RCT, 2 pre-post) reported an improvement. Two studies (1 RCT, 1 pre-post) reported no improvement. |
| Chien et al | Self-esteem | SR & MA | N (studies)=1, N (participants)=106, MD 0.5, 95% CI -1.22-2.22, Z=0.57 (P=0.57). No effect |
| Chien et al | Assertiveness | SR & MA | Medium term: N (studies)=1, N (participants)=458, MD 0.08, 95% CI -0.06-0.22, Z=1.15(P=0.25), I^2^=100%, Tau^2^=0; Chi^2^=0, df=0 (P<0.0001). No effect  Long term: N (studies)=1, N (participants)=447, MD 0.07, 95% CI -0.06-0.20, Z=1.05 (P=0.29). No effect |
| Lyons et al | Identity | SR & MA | Effects on identity were mixed. |
| Sun et al | Self-stigma | SR & MA | Peer intervention was associated with lower self-stigma, harm in self-stigma, application of stereotype, stigma protocol, and coping resources were significantly changed after the intervention |
| Sun et al | Stigma related stress | SR & MA | Stigma harm in stigma stress was significantly reduced after the intervention. |
| Fortuna et al | Perceived stigma | SR & NR | No effect |
| Schlichthorst et al | Psychological improvements | Scoping review | Improvements reported |

**Cost**

| **Author** | **Outcome** | **Type of review** | **Main findings** |
| --- | --- | --- | --- |
| Chien et al | Direct and indirect costs (Euro): total cost | SR & MA | Medium term: N (studies) = 1, MD 2092.0, 95% CI -72.00-4258.00)  Long term: N (studies) = 1, MD 775.0, 95% CI -1610.00-3160.00) |
| Huang et al | Costs of peer interventions | SR & MA | Evidence indicates that peer support may be a cost-saving intervention strategy. |
| Triece | Cost | SR & MA | One intervention was found to be low cost, and another was found to be cost-saving. |
| Gaiser et al | Cost | SR & NR | One study reported lower total Medicaid expenditures, and another reported peer support claims were associated with higher crisis stabilization service costs, lower psychiatric hospitalization costs, and higher total Medicaid costs. |

**Other**

| **Author** | **Outcome** | **Type of review** | **Main findings** |
| --- | --- | --- | --- |
| Chien et al | Peer outcomes - Improved peer contact | SR & MA | N (studies) = 1, N (participants) = 106, RR 1.85, 95% CI 1.14-3.00, Z=2.48 (P=0.01) |
| Chien et al | Peer outcomes - Negative overall relationship | SR & MA | N (studies)=1, N (participants)=105, MD=-0.19, 95% CI=-0.48, 0.1, Z=1.31(P=0.19), I^2^=100%, Tau^2^=0, Chi^2^=0 (df=0, p<.0001). No effect. |
| Chien et al | Peer outcomes - Positive overall relationship | SR & MA | N (studies)=1, N (participants)=105, MD=-0.43, 95% CI=0.16, 0.70, Z=3.11(P=0) |
| Chien et al | Peer outcomes - Social support for positive interactions | SR & MA | N (studies)=1, N (participants)=106, MD=5.6, 95% CI=-0.51-11.71, Z=1.8 (p=.07), I^2^=100%, Tau^2^=0; Chi2=0 (df=0, p<.0001). No effect. |
| Chien et al | Peer outcomes - Quality of life medium term - EuroQol: Five Dimensions-Visual Analogue Scale (EQ5D-VAS), General Quality of Life Inventory  (GQOLI-74), Manchester Short Assessment of Quality  of Life (MSAQOL), World Health Organisation Quality of Life  (WHOQOL), Quality of Life Brief Version (WHO-QOL-BREF) | SR & MA | EQ5D-VAS: N (studies)=1, N (participants)=216, MD=0.4, 95% CI=-4.52, 5.32, Z=0.16(p=0.87), I^2^=100%, Tau^2^=0, Chi^2^=0 (df=0, p<.0001). No effect  GQOLI-74: N (studies)=1, N (participants)=100, MD 40.34, 95% CI 32.70-47.98, Z=10.35(P<0.0001)  MSAQOL: N (studies)=1, N (participants)=208, MD 0.24, 95% CI -0.04-0.52, Z=1.7(P=0.09)  WHOQOL: N (studies)=1, N (participants)=106, MD 1.0, 95% CI -2.82-4.82, Z=0.51(P=0.61)  WHO-QOL-BREF: N (studies)=1, N (participants)=458, MD 0.20, 95% CI -0.33-0.73, Z=0.74 (P=0.46) |
| Chien et al | Peer outcomes - Quality of life -Long term - EuroQol: Five Dimensions-Visual Analogue Scale (EQ5D-VAS), World Health Organisation Quality of Life  (WHOQOL), Quality of Life Brief Version (WHO-QOL-BREF) | SR & MA | EQ5D-Index: N (studies)=1, N (participants)=216, MD=3.30, 95% CI=-1.83-8.43, Z=1.26(P=0.21)  EQ5D-VAS: N (studies)=1, N (participants)=216, MD=5.0, 95% CI=-0.67-10.67, Z=1.73(P=0.08)  MSAQOL: N (studies)=1, N (participants)=208, MD 0.24, 95% CI -0.04-0.52  WHOQOL: N (studies)=1, N (participants)=431, MD 0.70, 95% CI 0.15-1.25, Z=2.51(P=0.01)  WHO-QOL-BREF: N (studies)=1, N (participants)=106, MD 1.70, 95% CI -2.32-5.72, Z=0.83(P=0.41) |
| Akerblom et al | Service effectiveness and fidelity | Scoping review | Mixed results |

**Appendix 7**

Experiences of peer support (detailed themes)

| **Theme** | **Benefit/ challenge** | **Summary** | **PSW** | **Clients** | **NP Staff** | **Mixed*/**  **Other** | **Ref** |
| --- | --- | --- | --- | --- | --- | --- | --- |
| **What the PSW role can bring** |  |  |  |  |  |  |  |
| **Wellbeing and recovery** | Benefit | PSWs experienced improved wellness and recovery. The role enabled them to reframe and accept their illness and kept them engaged in the recovery process. PSWs also reported increased confidence, self-esteem, self knowledge and personal growth. This was through e.g. using their lived experience to help others, a sense of belonging, self-care, improved interpersonal skills, learning more about their own mental health and being reminded to use this knowledge [for their own recovery], and learning from clients. | ✓ |  |  |  | [57–59] |
|  | Challenge | PSW role could have a negative impact on wellbeing and recovery. Due to ‘sacrifices’ made for the job, the role reminding them of their illness, ‘sick’ label staying with PSWs, and a heavy workload. Clients could be a source of stress including clients who weren’t cooperative or had a greater level of disturbance than the PSWs own experience. | ✓ |  |  |  | [57,59] |
|  | Challenge | Mixed samples reported negative impacts on wellbeing for non-peer staff, PSWs and clients. PSW absenteeism due illness or relapse increased caseload for remaining staff. There is a risk that clients and PSWs could experience distress due to exposure to triggering content. There was fear that PSWs recovery process could negatively impact the support provided. |  |  |  | ✓c | [60–62] |
| **Recovery and role models** | Benefit | The importance of PSWs being role models was related to embodying personal recovery so they could be ‘the evidence’ of recovery. PSW roles aided personal recovery through e.g. providing a route back into employment and social inclusion, gaining skills, structure and stability. PSWs felt mutual benefits from their role, which validated their recovery enabling them to acknowledge their own strengths and achievements. | ✓ |  |  |  | [57,59,61] |
|  | Benefit | PSWs could be role models and inspire recovery for clients. From working with PSWs, clients experienced increased hope, motivation, better social communication skills and social networks, a sense of belonging and improved mental health symptoms. PSWs could show clients that life beyond illness is possible and how negative experiences could be transformed into positive.  Clients valued PSWs sharing their knowledge and skills and felt empowered as they gained knowledge on mental health, e.g. insights from recovery and the shared experience that they had gone through, increasing the relatability of PSWs. Gaining knowledge motivated clients to be optimistic and independent in their recovery. |  | ✓ |  |  | [58,61,63] |
|  | Benefit | Forming relationships with other mothers who had similar experiences was valuable for recovery and essential for maternal self-efficacy. The like-minded support enabled group members to re-evaluate themselves, others and the expectations of motherhood. |  |  |  | ✓e | [64] |
|  | Benefit | From working with PSWs, non-peer staff developed increased empathy and understanding toward people in recovery and gained a belief in recovery. |  |  | ✓ |  | [58] |
|  | Benefit | PSWs are role models, give clients hope of recovery, are valued and provide guidance and support to clients through the process of engaging with mental health services (e.g. how to navigate services). |  |  |  | ✓b | [65][62] |
|  | Challenge | Some papers reported that PSWs are not role models for clients. Reasons included a belief that without formal training and because of their mental health diagnosis PSWs would be ineffective helpers. |  | ✓ |  |  | [58] |
| **Career, social inclusion and identity** | Benefit | PSW roles offered a route back into employment, gaining skills, structure, stability, financial freedom, functioning and increased social networks and also social inclusion e.g. by interacting with non-peer staff, on an equal footing. The role could also be a stepping stone into further employment. The role positively impacted PSWs sense of self, e.g. they reported increased self-acceptance as they no longer had to hide their mental health problems, and enhanced self-esteem. The role also enabled PSWs to redefine themselves and their social value as they moved from service user to provider.  The PSW role enabled them to contribute through work, which helped maintain recovery, e.g. through enhancing the lives of clients struggling with their mental health. | ✓ |  |  |  | [57–59] |
|  | Benefit | PSWs found the role highly rewarding and important. PSW roles enabled service users to find a place in the community beyond ‘patient’ |  |  |  | ✓f | [58,60] |
| **Experiential knowledge, normalisation and reducing stigma** | Benefit | PSW support differed from formal treatment, it normalised and de-medicalised client experiences. This difference felt novel, relaxed and person-centred leading clients to reconnect with ‘real life’ situations, e.g. rebuilding relationships, which improved social functioning. Lack of judgement from PSWs reduced stigma around client experiences of an eating disorder. The sense of a ‘shared experience’ helped clients feel they were ‘getting back to normal’  Clients appreciated PSWs experiential knowledge, perceiving them to be more insightful than non-peer staff as they were viewed as role models in recovery, promoting greater empowerment and hope for clients. Peer-support services were valued by clients and they wanted to see their expansion. PSW services were regarded as a trusted platform that made them feel comfortable and accepted when attending activities, e.g. peer sharing. |  | ✓ |  |  | [61,63] |
|  | Benefit | PSW roles for organisations decreased stigma to mental health problems and set a positive example to other sectors. |  |  |  | ✓h | [58] |
|  | Challenge | Some clients and members of the public found it challenging to view the PSWs as mental health professionals, given concerns on their mental health history and risk of their relapse. Some clients perceived the knowledge shared by PSWs to be of lower value than that by healthcare professionals and should not be fully trusted |  | ✓ |  |  | [63] |
| **Isolation and validation** | Benefit | Having their experiences, e.g. that mothering in illness is difficult, validated by other mothers made life ‘less difficult’. |  |  |  | ✓e | [64] |
|  | Challenge | Meeting other mothers could lead to increased isolation, where their experiences were contrasting, e.g. feeling that others are happy when they are not. |  |  |  | ✓e | [64] |
| **Rapport and empathy with clients** | Benefit | Clients built rapport easier with PSW than non-peer staff due to PSWs having less professional distance and being ‘street smart’, e.g. knowledge about the effect of environment on drug use.  Clients felt that PSWs were more approachable and caring than non-peer staff, enabling them to open up and share their concerns. Clients also perceived greater empathy from PSWs, especially with regard to adverse effects from medications |  | ✓ |  |  | [58,63] |
| **Bridge** | Benefit | PSWs function as a bridge between clients and non-peer staff and within the organisation, by building trust based pathways, supporting the integration of the client across the fragmented care system. |  |  |  | ✓a | [65] |
| **Pioneer and expectations** | Benefit | PSWs were described as pioneers which led to certain expectations. Combined this led to pressure, i.e. no room for failure as this would reduce future PSW opportunities. |  |  |  | ✓a | [65] |
| **Complementary role, expertise and becoming part of the team** | Benefit | Non-peer staff recognised the valuable contribution of PSWs. PSWs fit in with various perspectives, becoming a legitimate team member. For example, PSWs could provide psychosocial support to clients, could replace the work of non-peer staff (e.g. accompanying clients to appointments), had time to do tasks that others may not (e.g. time to just talk to patients), and were valuable sources of experiences and information, e.g. as client advocates. PSWs may gather different knowledge of patients compared to non-peer staff, e.g. client may tell PSW (but not non-peer staff) about drug abuse. The importance of collaborating with PSWs was recognised, e.g. nurses could collaborate with PSWs to improve recovery oriented care. |  |  |  | ✓a | [65] |
|  | Challenge | PSWs may lack a broader perspective on mental health, past their own experience. |  |  |  | ✓a | [65] |
| **Confusion over the PSW role** |  |  |  |  |  |  |  |
| **Role ambiguity** | Benefit | When PSWs were introduced, their role was ambiguous this was positive as it gave flexibility to define the role. |  |  |  | ✓a | [65] |
|  | Challenge | A lack of clarity about the PSW job description meant that PSWs felt confused in their role which affected their confidence, perception of competence, with ramifications for their recovery and uncertainty in their responsibilities to clients. For example, a lack of clarity about the PSW job description led PSWs to feel the role was undervalued and tokenistic, and to feel uncertainty about where to seek support. | ✓ |  |  |  | [57,59] |
|  | Challenge | Some clients perceived a lack of clarity on the PSWs' roles: PSWs were viewed as informal staff who were replaceable, leading to negative perceptions of the PSW services. Some clients perceived peer support to be tokenistic, which led to the content of the PSW intervention ‘feeling irrelevant’. |  | ✓ |  |  | [61,63] |
|  | Challenge | PSWs found their role ambiguous making them anxious to demonstrate their value. Some PSWs felt they had received insufficient training for the role and were expected to develop the role over time. This hampered service delivery and created the perception that employing PSWs was tokenistic. Non-peer staff were unsure of the PSW role, leading to a lack of support from non-peer staff. |  |  |  | ✓b | [33,60] |
| **Boundaries** | Challenge | PSWs found managing the transition from patient to PSW and knowing where to draw the line between friend and service provider, to be challenging, e.g. they are expected to share personal information with clients. Working as a PSW in substance abuse could lead to a disconnect from their own recovery communities due to ethical concerns when sharing information in support groups, putting the PSWs recovery at risk. | ✓ |  |  |  | [57,58] |
|  | Challenge | Boundary issues included whether to relate to service users as friends (seen as unprofessional) or clients. Some PSWs would not share client information with agency staff due to concern about violating friendship with the client. |  |  |  | ✓c | [33,62] |
| **Role conflict and professionalisation** | Challenge | A challenging aspect of the PSW role was the dual identity as a service user and service provider which impacted on relationships and boundaries with non-peer staff and clients and was a source of stress. The dual identity was particularly difficult e.g. where PSWs were working in a team that previously cared for them. There were challenges with maintaining boundaries with clients who experienced similar difficulties to their own, i.e. this could be triggering for PSWs leading to a recurrence of their own mental health issues, they could also more closely connect with people who had similar difficulties but this could have an emotional impact on the PSWs. | ✓ |  |  |  | [59] |
|  | Challenge | The transition from patient to staff is challenging and there was ambiguity about whether PSWs should be seen as patients or staff. Non-peer staff may be concerned about the PSW becoming unwell, especially if they were previously a patient at the facility. This was a barrier to employing PSWs and meant that PSWs felt they were being treated as patients (e.g. repeatedly asked if they are okay). Staff acknowledge that PSWs should not be treated like patients, to challenge the view that consumers cannot recover.  PSWs can be ‘unwilling’ to give up their consumer perspective to adopt ‘professional beliefs and roles’. Managing the tension over professionalisation for PSWs was a challenge for organisations, e.g. training was questioned as leading to professionalisation and interference with the advantage of being a PSW. |  |  |  | ✓d | [33,58,65] |
| **Disclosure of peer status** | Benefit | For some PSWs sharing their story was connected to personal recovery and they progressed from telling an ‘illness story’ to a ‘recovery story’. | ✓ |  |  |  | [59] |
|  | Challenge | Disclosure of personal stories of recovery was intrinsic to the PSW role, however PSWs differed in how comfortable they felt about disclosure. Challenges around disclosure for PSWs included being labelled as ‘mentally unwell’, clients not valuing PSW knowledge due to the PSWs mental health condition, and concern about getting jobs outside of mental health due to their PSW identity.  Some PSWs expressed fears of being socially excluded and labelled as “mentally ill” thus they would avoid sharing their experiences because they believed clients would not trust them. | ✓ |  |  |  | [59,63] |
|  | Challenge | Confusion over when/with whom to disclose lived experience. Disclosure important to educate team on alternative views, e.g. PSW may say ‘I’ve experienced that…’. Disclosure may require discretion when fitting with professional relationships and maintaining boundaries. But ‘professionalism’ may not challenge existing boundaries which could change culture.  Disclosure of peer status affected peer integration, e.g. disclosure could build trust with clients. Some PSWs felt vulnerable and were reluctant to disclose. Staff role titles could identify (e.g. peer specialist)/not identify (e.g. programme aide) the PSW. |  |  |  | ✓a | [33,65] |
| **Organisational challenges and impact** |  |  |  |  |  |  |  |
| **Challenges for healthcare staff/organisations** | Challenge | Non-peer staff felt there were expectations to support, train and supervise PSWs, increasing their workload. Some staff found it challenging to manage having different ‘providers’ [PSWs] in the team. |  |  |  | ✓c | [60] |
|  | Challenge | Confidentiality, disclosure and increased sick time of PSWs compared to nonpeer workers were issues for organizations. |  |  |  | ✓f | [58] |
| **The value of the PSW role and low pay** | Challenge | The value of the PSW role was linked to low pay. Some PSWs struggled to identify benefits of the PSW role to the mental health system and saw themselves as ‘cheap labour’. PSWs had concerns about low pay, few hours and working overtime without compensation. Low pay contributed to role dissatisfaction. Although some PSWs felt they were well compensated and that the role offered empowerment. | ✓ |  |  |  | [57–59] |
|  | Challenge | PSWs received low pay, which was difficult as they wanted jobs that freed them from disability income. Low pay suggested the job was new, not valued or unclear. PSWs felt pay correlated with legitimacy and tokenism (i.e. if paid less than non-peer staff). Reasons for low pay were: hourly pay (instead of salaried), PSW not requiring certification (which could lead to higher pay), stigma from non-peer staff about 'the capacity for people with mental health conditions to work'.  There was variety in PSW pay, which has consequences for the continuity of peer support in teams. |  |  |  | ✓a | [33,65] |
| **Workload** | Challenge | PSW workload could be overwhelming. This could jeopardise other staff relationships, also under pressure from their own workload. Being given so many varying tasks (household tasks, participating in meetings, admin) the role could lose its distinctiveness. This was added to by a lack of understanding of the role of PSWs. |  |  |  | ✓a | [65] |
| **Lack of support and training** | Challenge | PSWs experienced a lack of support and training, potentially related to unclear job descriptions. For example, organisations are failing to provide support for PSWs with a known history of distress. PSWs struggled to develop the skills for their roles, including to work with clients with more complex needs than their own experiences. PSWs reported that their supervision felt superficial and problems in relationships between PSWs and supervisors were identified relating to disagreements and PSWs not feeling they had enough autonomy. | ✓ |  |  |  | [57,59] |
|  | Challenge | It was felt that lived experience wasn’t solely sufficient to work in interprofessional teams. Some PSWs were positive about certification, this could make other staff recognise peer support as a ‘genuine’ occupation. Other PSWs felt that certification could be in conflict with the idea of grassroots, user-led ethos.  Supervision and support were often not offered to PSWs. Risks might arise due to PSWs lack of training and support. Organisations needed to train PSWs and non-peer staff about the value of peer support and develop/implement guidelines. |  |  |  | ✓b | [62,65] |
| **Colleagues and stigma** | Challenge | PSWs could experience negative and rejecting non-peer staff attitudes, e.g. treated as a patient, rather than a colleague, talking inappropriately or joking about people with mental health issues, PSWs not invited to certain work activities, e.g. outside events. PSWs felt excluded, experienced tokenism and stigma, this could lead to isolation and self-stigma. Although some PSWs reported feeling accepted in their teams. | ✓ |  |  |  | [57–59] |
|  | Challenge | There was fear that ‘cheap labour’ provided by PSWs might lead to less non peer staff positions. |  |  | ✓ |  | [58] |
|  | Challenge | The PSW roles could be a threat to other professionals' roles e.g., nurses suspicious they may be replaced.  PSWs felt uncomfortable talking about their role due to stigma, they challenged stigma by taking on more responsibility. Hierarchies in teams undermined PSWs feeling equal in meetings. PSWs needed to find their voice to challenge clinically dominant ways of thinking.  Non-peer staff were uneasy about working alongside people they had previously treated or PSWs being able to see other individual records that may include their friends and other PSWs.  Concerns from healthcare professionals and policymakers over effectiveness and safety of peer-support services led to a lack of support and hostility from non-peer staff. Hence PSWs were accorded less respect and fewer responsibilities, with doubts consequently cast over their credibility. |  |  |  | ✓g | [63,65,66] |
| **Treatment models** | Challenge | PSWs are part of the newer recovery model and had trouble integrating into the traditional treatment model, e.g. where doctors held majority of power and decision making for clients but spent the least time with clients. PSWs expected to contest the traditional treatment model in support of a recovery focus (e.g. by their presence or in some cases by being openly challenging), this led to friction. If organisations are not prepared for PSWs the role doesn’t provide stable employment. | ✓ |  |  |  | [57] |
| **Other** |  |  |  |  |  |  |  |
| **Offering treatment choice** | - | Clients should have opportunities to choose among PSWs as service providers. |  |  |  | ✓c | [62] |

*Note. For ‘mixed’ samples the group or groups that stated the theme is unknown (e.g. PSW or non-peer staff or both).

PSW = Peer Support Worker; NP staff = non-peer staff

a= Mixed: non-peer staff, PSWs, clients, policy makers, peer programme developers; b = Mixed: PSW, non-peer staff, clients, carers, policy makers, peer programme developers; c = Mixed: clients, PSWs, carers, non-peer staff; d = Mixed: PSW, clients, policy makers, peer programme developers, non-peer staff, mental health organisations; e = Other: peer support group members; f = Other: Mental health organisations; Mixed: PSWs, non-peer staff, clients, carers; g = Mixed: PSW, clients, policy makers, peer programme developers, non-peer staff, other = mental health organisations, unspecified (in one study), h = Other: Mental health organisations
